# Trends in HIV testing, the treatment cascade, and HIV incidence among men who have sex with men in Africa: A systematic review and meta-regression analysis

**DOI:** 10.1101/2022.11.14.22282329

**Authors:** James Stannah, Nirali Soni, Jin Keng Stephen Lam, Katia Giguère, Kate M Mitchell, Nadine Kronfli, Joseph Larmarange, Raoul Moh, Marcelin N’zebo Nouaman, Gérard Menan Kouamé, Mathieu Maheu-Giroux, Marie-Claude Boily

## Abstract

**Background:** Gay, bisexual, and other men who have sex with men (MSM) are disproportionately affected by HIV. In Africa, MSM face structural barriers to HIV prevention and treatment including socio-economic disadvantages, stigma, and criminalization that increase their vulnerability to HIV acquisition and transmission and undermine progress towards ending AIDS. This systematic review explores progress towards increases in HIV testing, improving engagement in the HIV treatment cascade, and HIV incidence reductions among MSM in Africa.

**Methods:** We searched Embase, Medline, Global Health, Scopus, and Web of Science from January 1980-March 2022 for cross-sectional and longitudinal studies reporting HIV testing, knowledge of status, care, antiretroviral therapy (ART) use, viral suppression, and/or HIV incidence among MSM in Africa. We pooled surveys using Bayesian generalized linear mixed-effects models, used meta-regression to assess time trends, and compared HIV incidence estimates among MSM with those of all men.

**Findings:** Of 8,992 articles identified, we included 148 unique studies published from 2005-2022. HIV testing increased over time in Central/Western and Eastern Africa and in 2020, we estimate that 88% (95% credible interval (CrI) 57-97%) of MSM had tested in the past 12 months, but 66% (19-94%) of MSM living with HIV knew their HIV status, although this is probably underestimated given non-disclosure. Current ART use increased over time in Central/Western (OR_year_=1.4, 95%CrI 1.1-2.0, N=8) and Eastern/Southern Africa (OR_year_=1.4, 1.0-1.8, N=17) and in 2020 we estimate that 75% (18-98%) of MSM living with HIV in Africa were currently on ART. Nevertheless, we did not find strong evidence viral suppression increased, and in 2020 we estimate that only 62% (12-95%) of MSM living with HIV were virally suppressed. HIV incidence among MSM did not decrease over time (IRR_year_=1.0, 0.7-1.3, N=38) and remained high in 2020 (5.4 per 100 person-years, 0.9-33.9) and substantially higher (27-150 times higher) than among all men.

**Interpretation:** No decreases in HIV incidence have been observed among MSM in Africa over time, despite some increases in HIV testing and ART use. Achieving the UNAIDS 95-95-95 targets for diagnosis, treatment, and viral suppression equitably for all requires renewed focus on this key population. Combination interventions for MSM are urgently required to reduce disparities in HIV incidence and tackle the social, structural, and behavioural factors that make MSM vulnerable to HIV acquisition.

**Funding:** US National Institutes of Health, UK Medical Research Council, Canadian Institutes of Health Research, Fonds de Recherche du Québec – Santé.

## INTRODUCTION

Globally, gay, bisexual, and other men who have sex with men (MSM) and other key populations experience a disproportionate burden of HIV^1^. Key populations are individuals who are vulnerable to HIV acquisition and transmission and who experience unmet HIV prevention needs. In 2021, members of key populations and their sexual partners accounted for 70% of new annual HIV infections globally and 21% occurred among MSM^1^. Globally, MSM may be up to 28 times more likely to acquire HIV compared to heterosexual men ^1^. Vulnerabilities to HIV can be partly explained by sexual behaviours, but the sociocultural and political contexts in which MSM live are important drivers of these vulnerabilities. Today, MSM face criminalization in 70 countries, including 31 in Africa^2^. In many settings, they are marginalized for their sexual identities and behaviours and face violence, stigma, and discrimination^1-4^. These punitive and discriminatory norms and legislations often impede access to primary HIV prevention and the treatment and care cascade, exacerbating susceptibilities to HIV acquisition and transmission. To “End AIDS”, the Global AIDS Strategy 2021-2026 calls for equitable and equal access to HIV services, as well as breaking down legal and societal barriers to achieving HIV outcomes^5^.

In 2021, an estimated 18% of new annual HIV acquisitions in Central and Western Africa occurred among MSM, compared to 3% in Eastern and Southern Africa, where epidemics are more generalized^1^. Despite this, HIV prevalence is much higher among MSM than the general population in all regions of Africa, highlighting the need for contextualized approaches to HIV prevention, and MSM-focused interventions across epidemic typologies^1^.

As with other key populations, MSM can be hard to reach, and nationally representative data on HIV service utilization and incidence are not available. This poses challenges to evaluating progress towards ending AIDS. The UNAIDS 95-95-95 targets for 2025 calls for 95% knowledge of status among those living with HIV, 95% treatment coverage among those diagnosed, and 95% viral suppression among those on treatment. Increasingly, dedicated surveys are being carried out to collect such indicators among MSM in Africa and to identify barriers and improve uptake of services to reduce new infections.

A previous systematic review and meta-analysis of HIV testing and the HIV treatment cascade from 2004-2017 among MSM in Africa reported that levels of diagnosis, treatment, and viral suppression were too low to achieve the previous UNAIDS 90-90-90 targets for 2020^6^. Here, we update and substantially expand on this previous review to improve our understanding of temporal trends in HIV testing, knowledge of status, care, treatment coverage, and viral suppression, and HIV incidence among MSM, and to evaluate progress towards achieving the new UNAIDS 95-95-95 targets for 2025 and ending AIDS among MSM in Africa.

## METHODS

### Search strategy and data extraction

We searched Web of Science, Scopus, and Ovid Embase, MEDLINE, and Global Health online databases for articles reporting HIV testing, knowledge of status, engagement in care, antiretroviral therapy (ART) use, viral suppression, or HIV incidence among MSM in Africa, published from January 1^st^, 1980, up to March 24^th^, 2022, using search terms for HIV, MSM, and Africa (Table S1).

We first screened articles by title and abstract, and then screened full texts for eligible studies. We included peer-reviewed cross-sectional or longitudinal studies that were conducted in any African country. We excluded conference abstracts, posters, and presentations, review articles, mathematical modelling studies, qualitative studies, and policy analyses. We did not exclude studies based on language. We perused the bibliographies of reviews and full texts for further relevant articles.

From the included studies, we extracted or calculated the following outcomes:

1. proportions of MSM who self-reported ever testing for HIV;
2. proportions of MSM who self-reported testing for HIV in the past 3, 6, and 12 months;
3. proportions of MSM living with HIV (confirmed with a biomarker) who knew they were living with HIV (from self-reports only or complemented with biomarkers, hereafter referred to as HIV aware MSM);
4. proportions of MSM living with HIV who self-reported engagement in care (as defined by study authors)
5. proportions of MSM living with HIV or HIV aware MSM, who were currently on ART (from self-reports or biomarkers)
6. proportions of MSM living with HIV, HIV aware MSM, or MSM currently on ART who were virally suppressed (confirmed with viral load testing and based on viral thresholds defined by study authors);
7. HIV incidence rates among MSM.

We also extracted information on participants (e.g., study population, age,), study characteristics (e.g., study design, region of Africa, country, study years), and indicators of study quality (e.g., sampling methods, definitions of MSM employed by studies, and interview methods).

When multiple articles reported observations of the same outcome from the same study, we extracted the observation derived from the largest sample size. For studies that included transgender women (TGW), where possible, we included observations among MSM only, otherwise we used the aggregate observation reported. For studies conducted in multiple countries, we extracted observations for each country separately, if reported, otherwise we used the aggregate observation but did not assign it a specific country. For studies conducted in multiple sub-national regions of a single country, we extracted only the aggregate observation. In studies of HIV incidence that reported multiple observations over consecutive non-overlapping follow-up periods, these were, otherwise we considered only the incidence rate covering the total follow-up period. In studies that reported them, weighted observations that accounted for sampling method (e.g., respondent-driven sampling, cluster, or time-location sampling) were preferred over to crude observations (Text S1).

Screening and data extraction were conducted by three independent reviewers (JS, NS, and JKSL). Discrepancies were resolved by KG. This systematic review and meta-regression analysis were completed according to PRISMA guidelines^7^.

### Data analyses

To pool observations and obtain region- and country-level estimates of HIV testing, stages in the HIV treatment cascade, and HIV incidence over time, we performed meta-regression analyses using Bayesian generalized linear mixed-effects models. Outcomes needed a minimum of 10 survey observations to be pooled. A Bayesian multilevel framework was chosen because MSM survey estimates are heterogenous, data is sparse geographically, and few countries have several surveys^6^. In our models, we included study-level random intercepts, nested within country and region, allowing us to improve the accuracy and precision of estimates in settings with fewer observations^8^. To assess time trends, we used the mean-centered calendar year (or year and month for HIV incidence) as a continuous variable (using the midpoint year of each study), with random slopes by country, nested within regions. Regions were classified as Central/Western, Eastern/Southern, and Northern Africa based on UNAIDS’ classifications^1^. If both Central and Western Africa or Eastern and Southern Africa had >10 survey observations, we included those regions separately in our analyses. We modelled proportions of ever and recent HIV testing, knowledge of status, ART use, and viral suppression using a binomial likelihood. For HIV incidence rates, we used a Poisson likelihood with log(person-time) as an offset. We did not weight countries by their estimated numbers of MSM given wide variations and uncertainties in these population size estimates^9-11^. We used non-informative prior distributions on the model parameters and elicited weakly informative prior distributions on the group-level variance parameters of the random effects (Text S2). Posterior distributions were obtained using Hamiltonian Monte Carlo, implemented in Stan^12^, and summarized using medians and 95% credible intervals (CrI). We reported time trends for countries with observations from at least three different time points. Finally, we compared our estimates of knowledge of status, current ART use, and viral suppression among MSM living with HIV, and HIV incidence with year-matched UNAIDS estimates for all men^13^.

We assessed the risk of bias in included studies by appraising studies according to five criteria covering the appropriateness of the sampling method, statistical adjustment for complex survey design, the representativeness of the study population of MSM, the inclusion of transgender women as MSM in surveys, and the risk of misclassification in ascertaining study outcomes (further details in Text S3). Studies received a score ranging from 0-5 for each outcome reported, representing higher (score 0-1), moderate (score 2-3), and lower (score 4-5) risk of bias in reported study outcomes. We assessed publication bias using funnel plots.

Analyses were conducted in R, version 4.0.2, using the “brms” and “rstan” packages^14,15^.

## RESULTS

### Search results

We initially identified 19,419 publications and, after removing 10,427 duplicates, and 7,921 publications at the title and abstract screening stage, we assessed the eligibility of 1,071 full texts (Figure S1). Four additional articles were identified from bibliographies of relevant articles. Overall, we included 231 articles from 147 unique studies (Table 1), nearly doubling the number of studies identified in a previous systematic review (Figure S2)^6^.

**Table 1.**
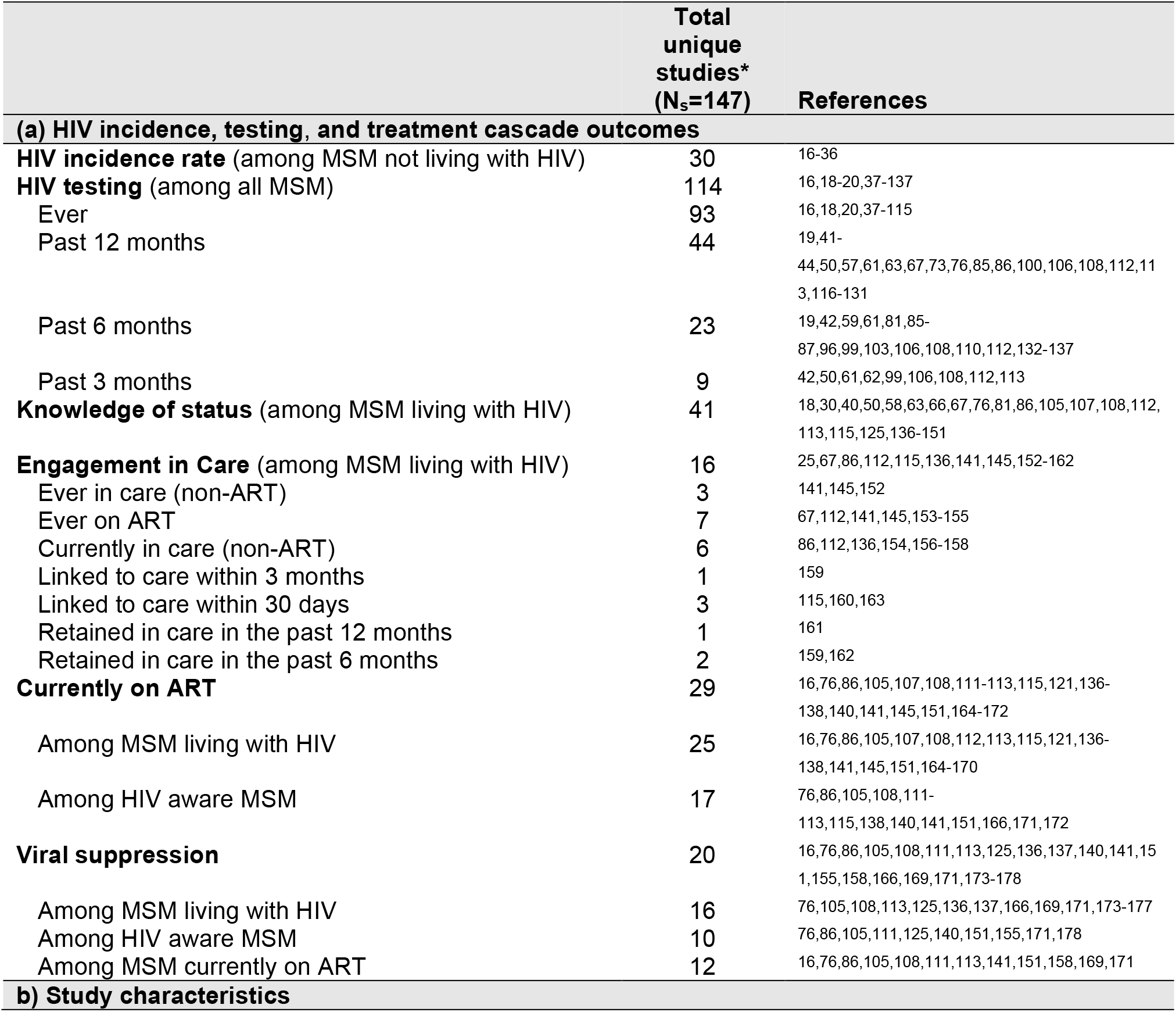
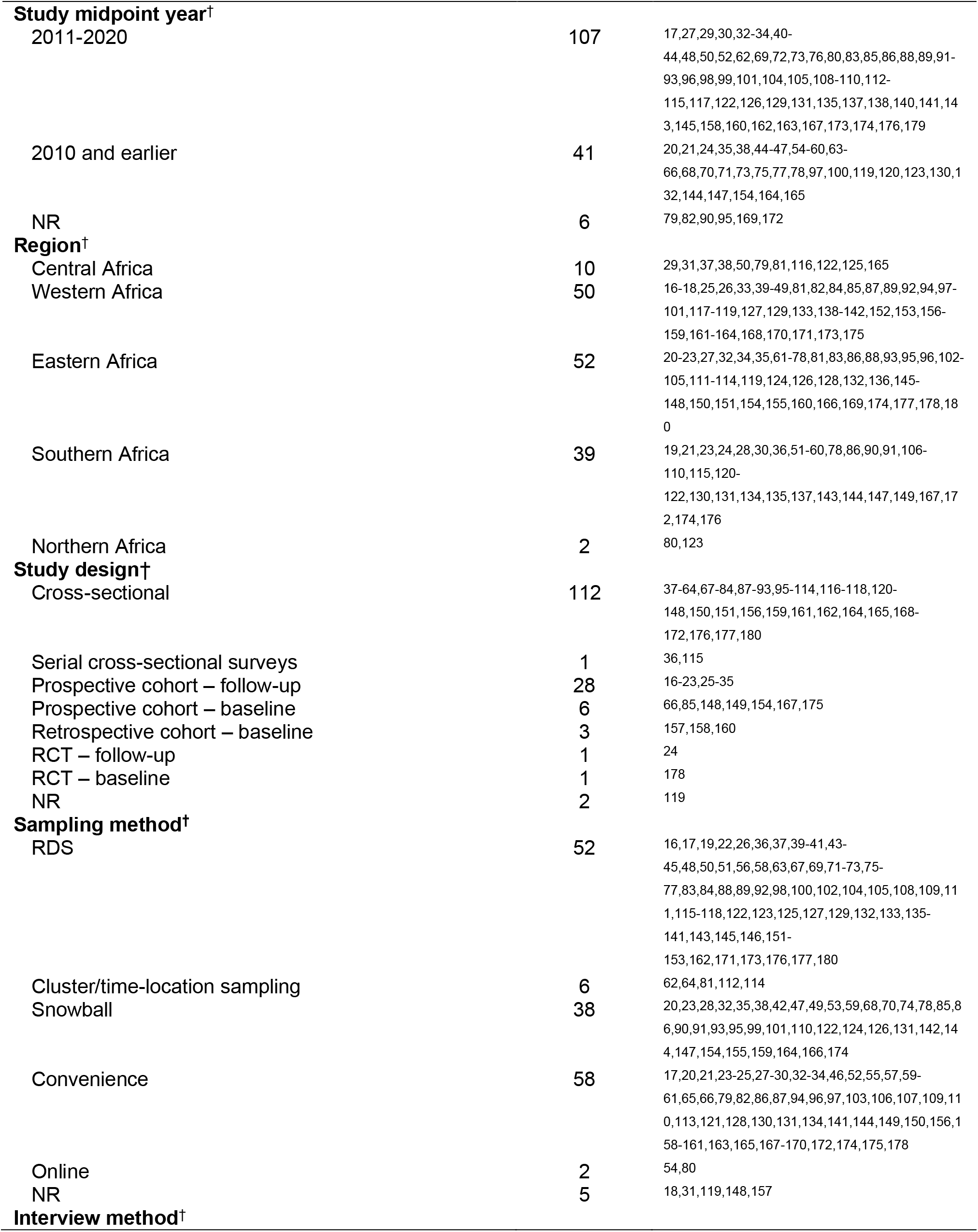
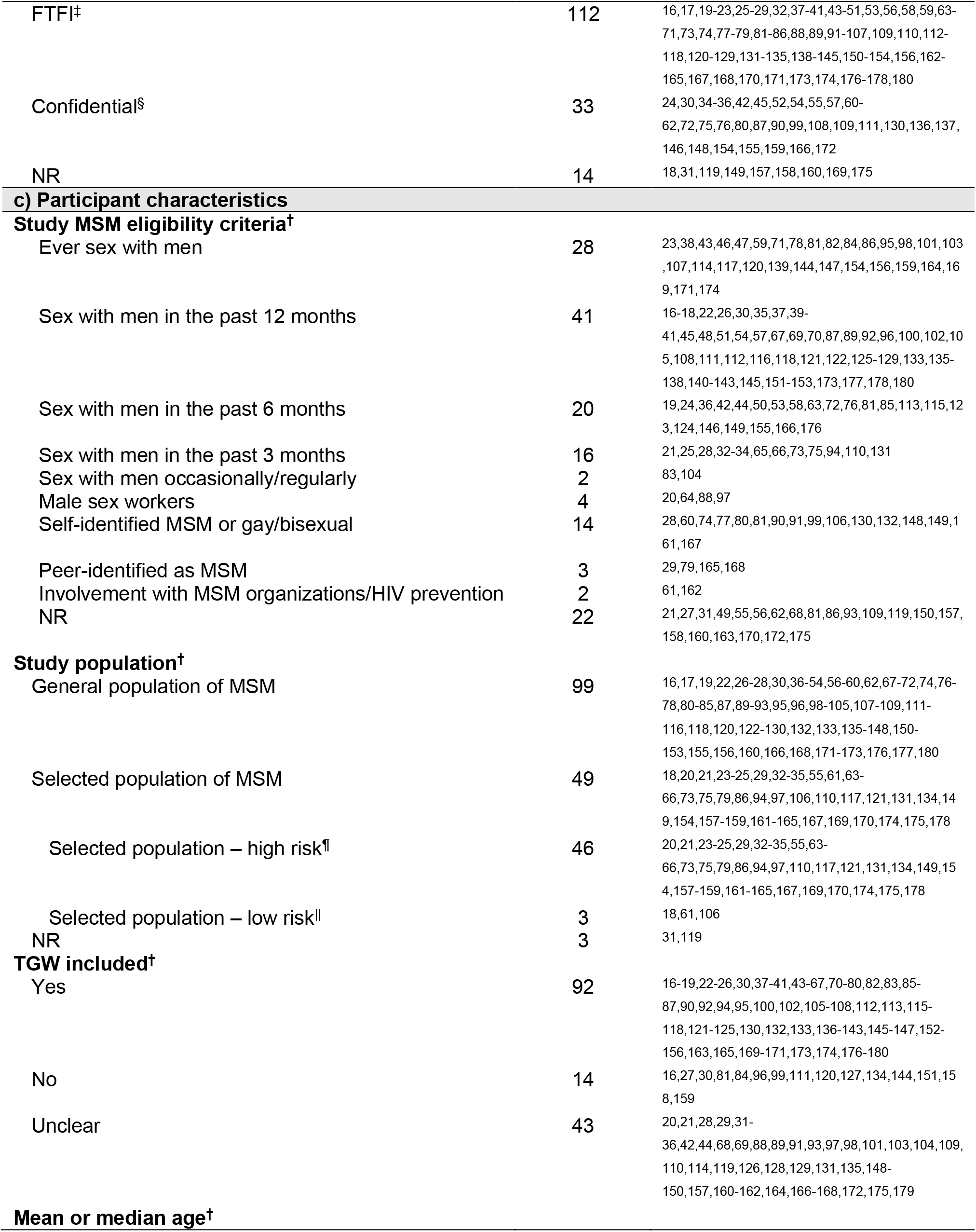
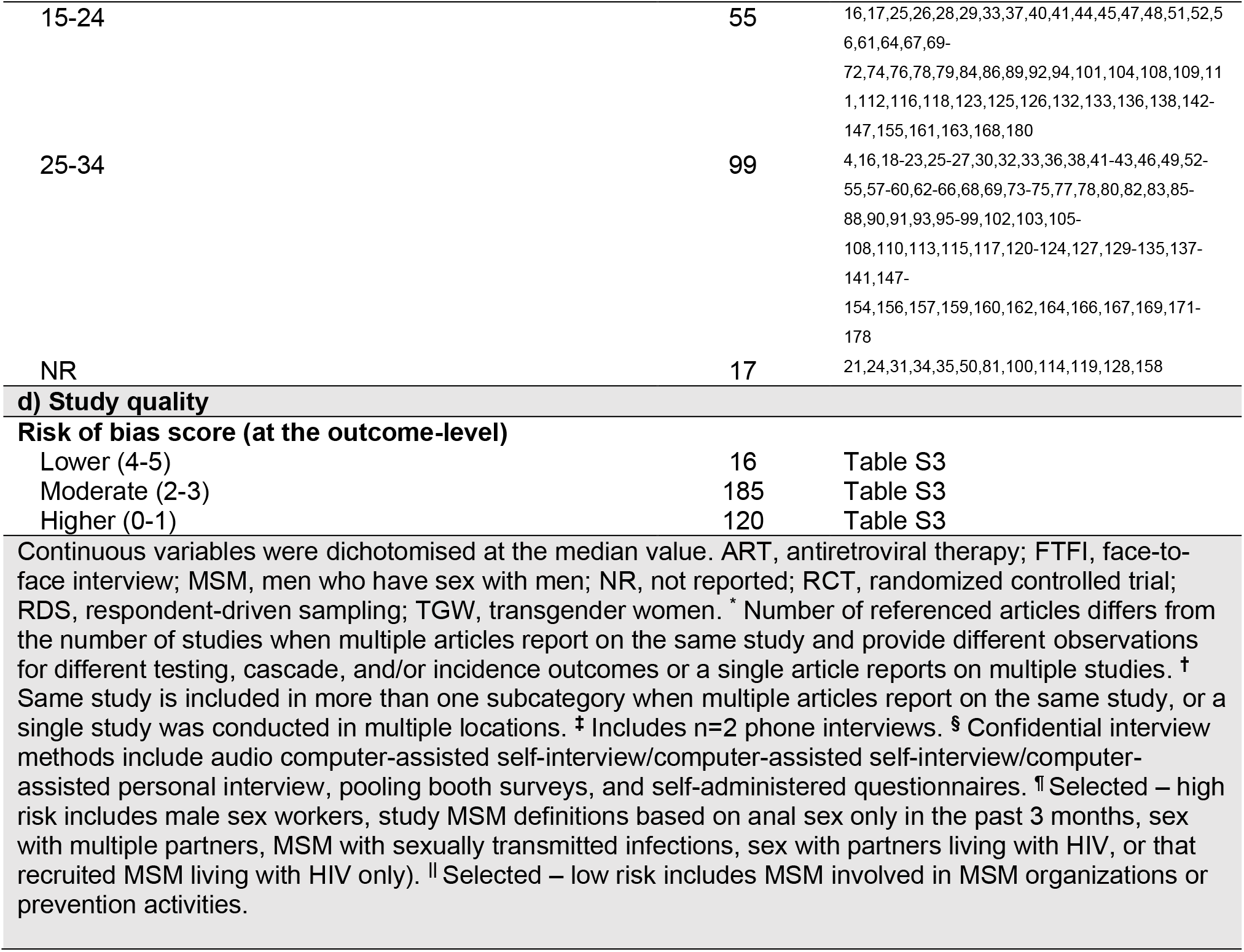
Number of unique studies included in our review that reported (a) HIV incidence, testing, and treatment cascade outcomes among men who have sex with men (MSM) in Africa, and summary of (b) study characteristics, (c) participant characteristics, and (d) study quality, of included studies, and references of individual publications that provided observations that were included in our analyses.

Included studies predominantly reported ever HIV testing (number of studies [N_s_]=93, number of independent observations [N_o_]=93, number of MSM [N_MSM_]=43,628), testing in the past 12 months (N_s_=44, N_o_=44, N_MSM_=21,865), and knowledge of status (N_s_=41, N_o_=41, N_MSM_=6,403; Table 1a). Fewer studies reported testing over shorter recall periods (N_s_=27, N_o_=33, N_MSM_=9,298), MSM currently on ART (N_s_=29, N_o_=44, N_MSM_=4,301), MSM virally suppressed (defined based on viral loads ranging from ≤20 to <1000 copies per mL; N_s_=20, N_o_=38, N_MSM_=2,981), or HIV incidence (N_s_=30, N_o_=38, N_MSM_=5,044; Table 1a). Few observations of engagement in care other than current ART use were available.

Most studies were conducted between 2011-2020 (N_s_=102) and in Western (N_s_=49), Eastern (N_s_=49), and Southern (N_s_=39) Africa (Table 1b). Few studies were from Central (N_s_=9) or Northern (N_s_=2) Africa. Observations were available from 30 countries, including 26 countries with HIV testing data, 24 countries with HIV treatment cascade data, and 12 countries with HIV incidence data. In 96 studies, conducted in 23 countries, sexual partnerships between men were criminalized at the time the study was conducted (Table 1b).

Studies reporting HIV testing and treatment cascade outcomes were primarily cross-sectional (N_s_=121), whilst incidence studies were mostly prospective cohort (N_s_=28). Most studies used convenience sampling (N_s_=58) or respondent-driven sampling (RDS, N_s_=52; Table 1b). When recruiting participants, most studies defined MSM using eligibility criteria based on self-reported sexual behaviours (e.g., anal, anal/oral, and anal/oral/masturbatory sex) with men in the past 12 months (N_s_=41), and participants were mainly recruited from the general population of MSM (N_s_=89; Table 1c). However, in over 90% of studies, MSM definitions either included transgender women, or it was unclear whether they did (Table 1c). Overall, study sample sizes ranged from 23 to 5,796 MSM. Enrolled MSM were largely young, with a mean or median age ranging between 25-34 years (N_o_=85; Table 1c). Face-to-face interviews were primarily used to collect self-reported information (N_s_=112; Table 1b).

### HIV testing, the treatment cascade, and HIV incidence among MSM in Africa: estimates for 2020 and time trends

In 2020, we estimated from self-reports that 87% (95%CrI 31-97%) of MSM had ever tested for HIV (Table 2). We estimated that ever HIV testing increased from 64% (95%CrI 54-74%) in 2010 to 88% (73-95%) in 2020 in Central/Western Africa (Odds Ratio per year [OR_year_]=1.15, 95% CrI 1.02-1.28, N_o_=33), and from 61% (50-71%) in 2010 to 93% (81-97%) in 2020 in Eastern Africa (OR_year_=1.24, 1.09-1.40, N_o_=33), increasing particularly in Kenya and Tanzania (Figure 1, Table 2, Figure S3, S16). Most observations were available from South Africa and Kenya.

**Table 2.** Estimated time trends in HIV testing, treatment cascade, and HIV incidence among men who have sex with men (MSM) in Africa and estimated outcomes in 2010 and 2020, overall and by region of Africa.

| Outcome | Region of Africa | N<sub>o</sub> | Estimate of time trend | 95% CrI | Estimate in 2010 | 95% CrI | Estimate in 2020 | 95% CrI |
| --- | --- | --- | --- | --- | --- | --- | --- | --- |
| **Ever HIV testing (%)** |  | 89 |  |  |  |  |  |  |
| Among all MSM* | Overall | 89 | OR=1.11 | 0.79–1.41 | 66% | 46–85% | 87% | 31–97% |
|  | Central/Western Africa | 33 | **OR=1.15** | **1.02–1.28** | **64%** | **54–74%** | **88%** | **73–95%** |
|  | Eastern Africa | 33 | **OR=1.24** | **1.09–1.40** | **61%** | **50–71%** | **93%** | **81–97%** |
|  | Southern Africa | 22 | OR=1.10 | 0.92–1.27 | 69% | 56–80% | 86% | 58–96% |
| **Past 12 months HIV testing (%)** |  | 44 |  |  |  |  |  |  |
| Among all MSM† | Overall | 44 | OR=1.23 | 0.98–1.53 | 48% | 30–66% | 88% | 57–97% |
|  | Central/Western Africa | 17 | **OR=1.23** | **1.06–1.45** | **48%** | **34–63%** | **88%** | **71–96%** |
|  | Eastern Africa | 14 | **OR=1.26** | **1.09–1.48** | **45%** | **29–60%** | **89%** | **72–96%** |
|  | Southern Africa | 12 | OR=1.21 | 0.99–1.44 | 50% | 34–67% | 87% | 60–96% |
| **Knowledge of status (%)** |  | 41 |  |  |  |  |  |  |
| Among MSM living with HIV | Overall | 41 | OR=1.23 | 0.98–1.53 | 15% | 4–41% | 66% | 19–94% |
|  | Central/Western Africa | 9 | **OR=1.23** | **1.06–1.45** | **13%** | **4–33%** | **65%** | **20–95%** |
|  | Eastern Africa | 17 | **OR=1.26** | **1.09–1.48** | **12%** | **5–26%** | **65%** | **34–88%** |
|  | Southern Africa | 15 | OR=1.24 | 0.89–1.71 | 22% | 10–44% | 66% | 22–89% |
| **Currently on ART (%)** |  | 42 |  |  |  |  |  |  |
| Among MSM living with HIV | Overall | 25 | OR=1.37 | 0.82–2.25 | 11% | 1–67% | 75% | 18–98% |
|  | Central/Western Africa | 8 | **OR=1.42** | **1.08–1.95** | **10%** | **2–34%** | **78%** | **41–96%** |
|  | Eastern/Southern Africa | 17 | **OR=1.36** | **1.04–1.82** | **11%** | **2–39%** | **73%** | **39–92%** |
| Among HIV aware MSM | Overall | 17 | OR=1.41 | 0.73–2.58 | 25% | 1–93% | 92% | 31–100% |
|  | Central/Western Africa | 5 | OR=1.43 | 0.79–2.05 | 24% | 2–84% | 92% | 28–100% |
|  | Eastern/Southern Africa | 12 | OR=1.45 | 0.98–2.17 | 25% | 3–74% | 93% | 63–99% |
| **Viral suppression (%)** |  | 35 |  |  |  |  |  |  |
| Among MSM living with HIV | Overall | 16 | OR=1.28 | 0.69–2.30 | 12% | 0–86% | 62% | 12–95% |
|  | Central/Western Africa | 4 | OR=1.30 | 0.81–2.20 | 11% | 1–69% | 64% | 18–96% |
|  | Eastern/Southern Africa | 12 | OR=1.29 | 0.95–1.79 | 11% | 1–48% | 61% | 32–84% |
| Among HIV aware MSM | Overall | 10 | OR=1.16 | 0.52–2.50 | 43% | 1–99% | 78% | 9–99% |
|  | Central/Western Africa | 3 | OR=1.13 | 0.46–2.62 | 51% | 1–99% | 79% | 5–99% |
|  | Eastern/Southern Africa | 7 | OR=1.20 | 0.68–2.08 | 36% | 2–95% | 78% | 23–97% |
| Among MSM currently on ART | Overall | 11 | OR=1.17 | 0.49–2.62 | 66% | 1–100% | 91% | 24–100% |
|  | Central/Western Africa | 4 | OR=1.23 | 0.51–3.11 | 67% | 1–100% | 93% | 28–100% |
|  | Eastern/Southern Africa | 7 | OR=1.16 | 0.56–2.35 | 65% | 2–100% | 89% | 39–99% |
| **HIV incidence rate (py<sup>-100</sup>)** |  | 38 |  |  |  |  |  |  |
| Among MSM not living with HIV | Overall | 38 | IRR=0.97 | 0.73–1.32 | 7.4py<sup>-100</sup> | 1.9–27.3 | 5.4py<sup>-100</sup> | 0.9–33.9 |
|  | Central/Western Africa | 17 | IRR=0.97 | 0.82–1.16 | 8.1py<sup>-100</sup> | 3.1–20.6 | 5.9py<sup>-100</sup> | 2.3–14.8 |
|  | Eastern/Southern Africa | 21 | IRR=0.97 | 0.82–1.16 | 6.8py<sup>-100</sup> | 3.0–15.2 | 5.0py<sup>-100</sup> | 1.6–14.7 |

ART, antiretroviral therapy; CrI, credible interval; IRR, incidence rate ratio; MSM, men who have sex with men; N<sub>o</sub>, number of observations; OR, odds ratio; py<sup>-100</sup>, per 100 person-years.
\* n=1 observation from Northern Africa included in analysis but not shown
† n=1 observation from Northern Africa included in analysis but not shown

**Figure 1.**
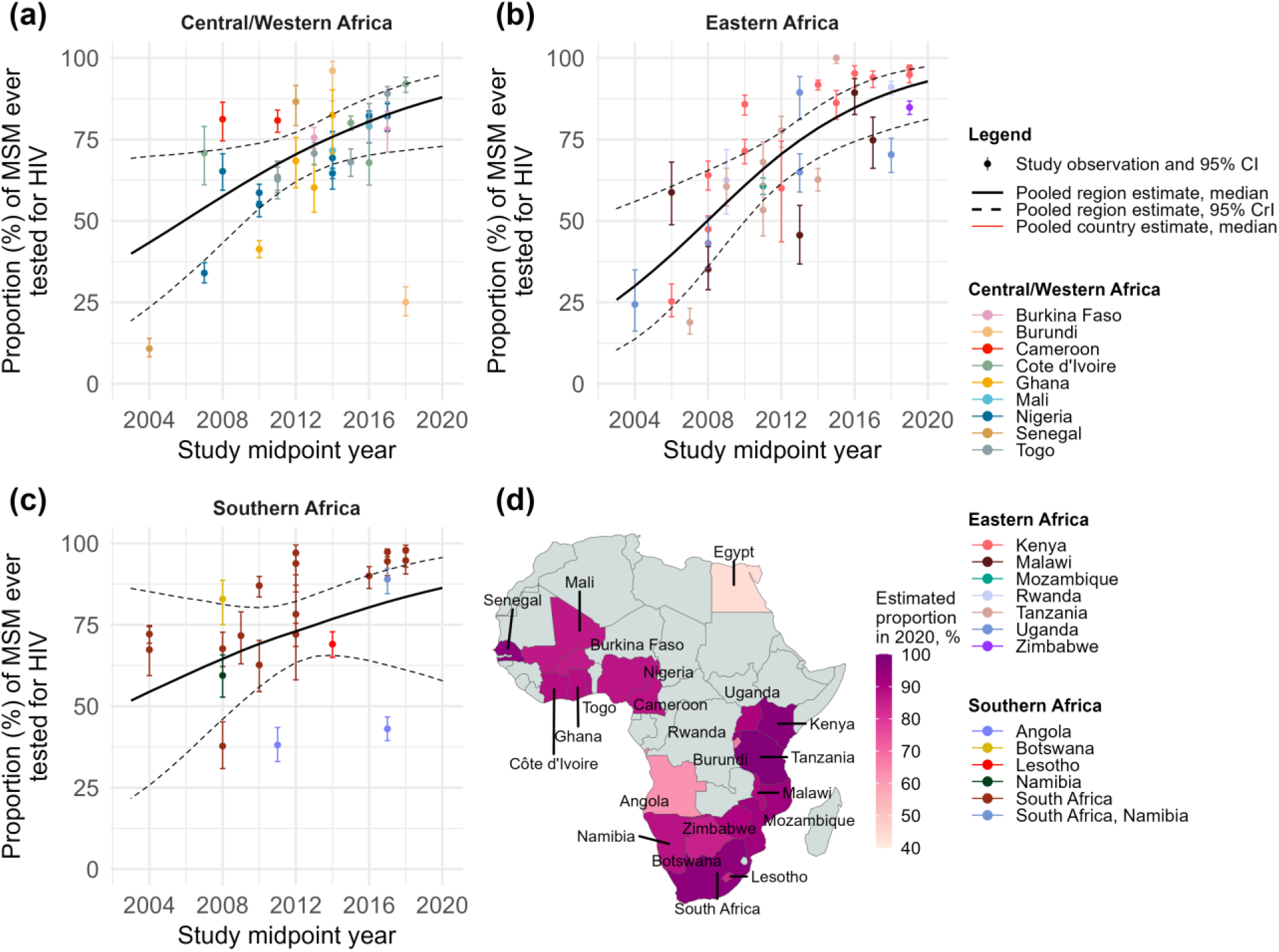
Ever HIV testing among men who have sex with men (MSM) over time, by region and country of Africa. Ever HIV testing among MSM in (a) Central/Western Africa, (b) Eastern Africa, and (c) Southern Africa, and (d) the estimated proportion of MSM ever tested for HIV in 2020, by country, estimated using a Bayesian logistic generalized linear mixed-effects model, with study-, country-, and region-level random effects. Points represent available study observations and their 95% confidence intervals, coloured by country in which the study was conducted. The black solid and dotted lines represent the estimated region-level proportions and 95% credible intervals (CrI), respectively. Coloured solid lines represent estimated country-level proportions for countries with at least 3 estimates from 3 different time points (see Figure S16 for individual country trends and 95% CrI).

In 2020 we estimated that 88% (57-97%) of MSM in Africa had been tested for HIV in the past 12 months (Table 2). Testing in the past 12 months increased from 48% (34-63%) in 2010 to 88% (71-96%) in 2020 in Central/Western Africa (OR_year_=1.23, 1.06-1.45, N_o_=17), and from 45% (29-60%) in 2010 to 89% (72-96%) in 2020 in Eastern Africa (OR_year_=1.26, 1.09-1.48, N_o_=14), although only one observation was available for most countries (Figure 2, Table 2, Figure S4, S17). In the Southern Africa region, ever and past 12 months testing both seemed to increase, but trends were inconclusive, although clear increases occurred in South Africa (Figure 1c, 2c, Table 2, Figure S16-17). There were not enough observations from Northern Africa, and for HIV testing in the past 3 months, to assess time trends (Figure S6). Time trends in past 6 months HIV testing were inconclusive (Table S2, Figure S5).

**Figure 2.**
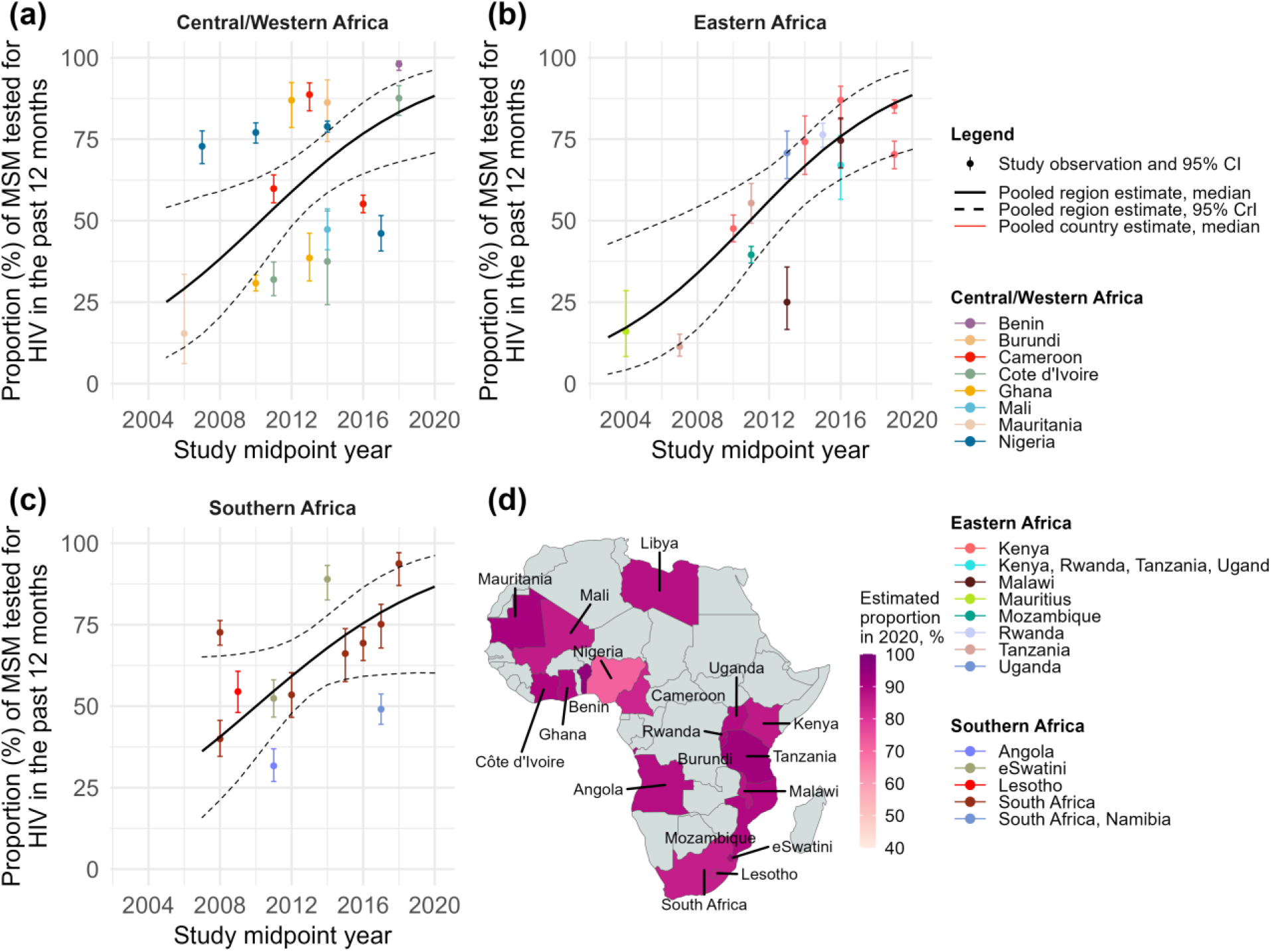
HIV testing in the past 12 months among men who have sex with men (MSM) over time, by region and country of Africa. Past 12 months HIV testing in (a) Central/Western Africa, (b) Eastern Africa, (c) Southern Africa, and (d) the estimated proportion of MSM tested for HIV in the past 12 months in 2020, by country. Points represent available study observations and their 95% confidence intervals, coloured by country in which the study was conducted. The black solid and dotted lines represent the estimated region-level proportions and 95% credible intervals (CrI), respectively. Coloured solid lines represent estimated country-level proportions for countries with at least 3 estimates from 3 different time points (see Figure S17 for individual country-level time trends and 95% credible intervals). N=1 observation from Mauritius not shown on the map.

Among MSM living with HIV, we estimated that knowledge of status in 2020 was 66% (19-94%). Knowledge of status increased substantially over time from 13% (4-33%) in 2010 to 65% (20-95%) in 2020 in Central/Western Africa (OR_year_=1.23, 1.06-1.45, N_o_=9) and from 12% (5-26%) in 2010 to 65% (34-88%) in 2020 in Eastern Africa (OR_year_=1.26, 1.09-1.48, N_o_=17) (Figure 3, Table 2, Figure S7, S18). Similar increases in Southern Africa were observed, although trends were inconclusive (Figure 3c, Table 2). In all regions, observations of knowledge of status were heterogenous, and overall only six countries had multiple observations.

**Figure 3.**
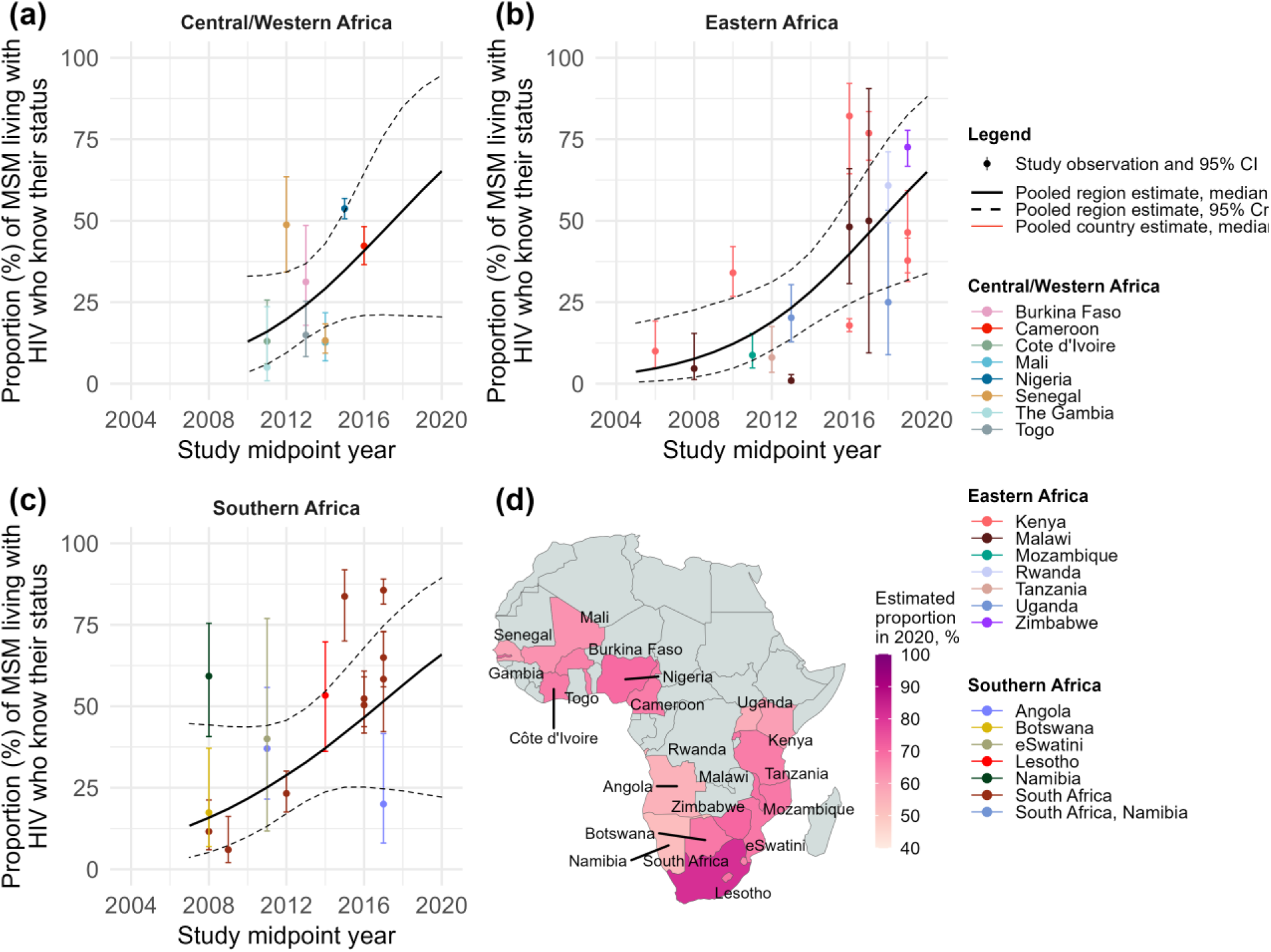
Knowledge of status among men who have sex with men (MSM) living with HIV over time, by region and country of Africa. Knowledge of status in (a) Central/Western Africa, (b) Eastern Africa, (c) Southern Africa, and (d) the estimated proportion of MSM living with HIV who know their status in 2020, by country. Points represent available study observations and their 95% confidence intervals, coloured by country in which the study was conducted. The black solid and dotted lines represent the estimated region-level proportions and 95% credible intervals (CrI), respectively. Coloured solid lines represent estimated country-level proportions for countries with at least 3 estimates from 3 different time points (see Figure S18 for individual country-level time trends and 95% CrI).

With the exception of current ART use, there were too few observations of the remaining engagement in care outcomes (e.g., ever or currently receiving non-ART care, retention in care in the past 12 months, ever ART use) to investigate time trends (Text S4, Figure S8-9).

Among MSM living with HIV, we estimated that 75% (18-98%) of them would be on ART in 2020. Current ART use among MSM living with HIV increased from 10% (2-34%) in 2010 to 78% (41-96%) in 2020 in Central/Western Africa (OR_year_=1.42, 1.08-1.95, N_o_=8), and from 11% (2-39%) in 2010 to 73% (39-92%) in 2020 in Eastern/Southern Africa (OR_year_=1.36, 1.04-1.82, N_o_=17) (Figure 4, Table 2, Figure S10, S19). Time trends in current ART use among those aware were similar, albeit inconclusive, and in 2020 current ART use among diagnosed MSM was 92% (31-100%; Table 2, Figure S11, S20-21).

**Figure 4.**
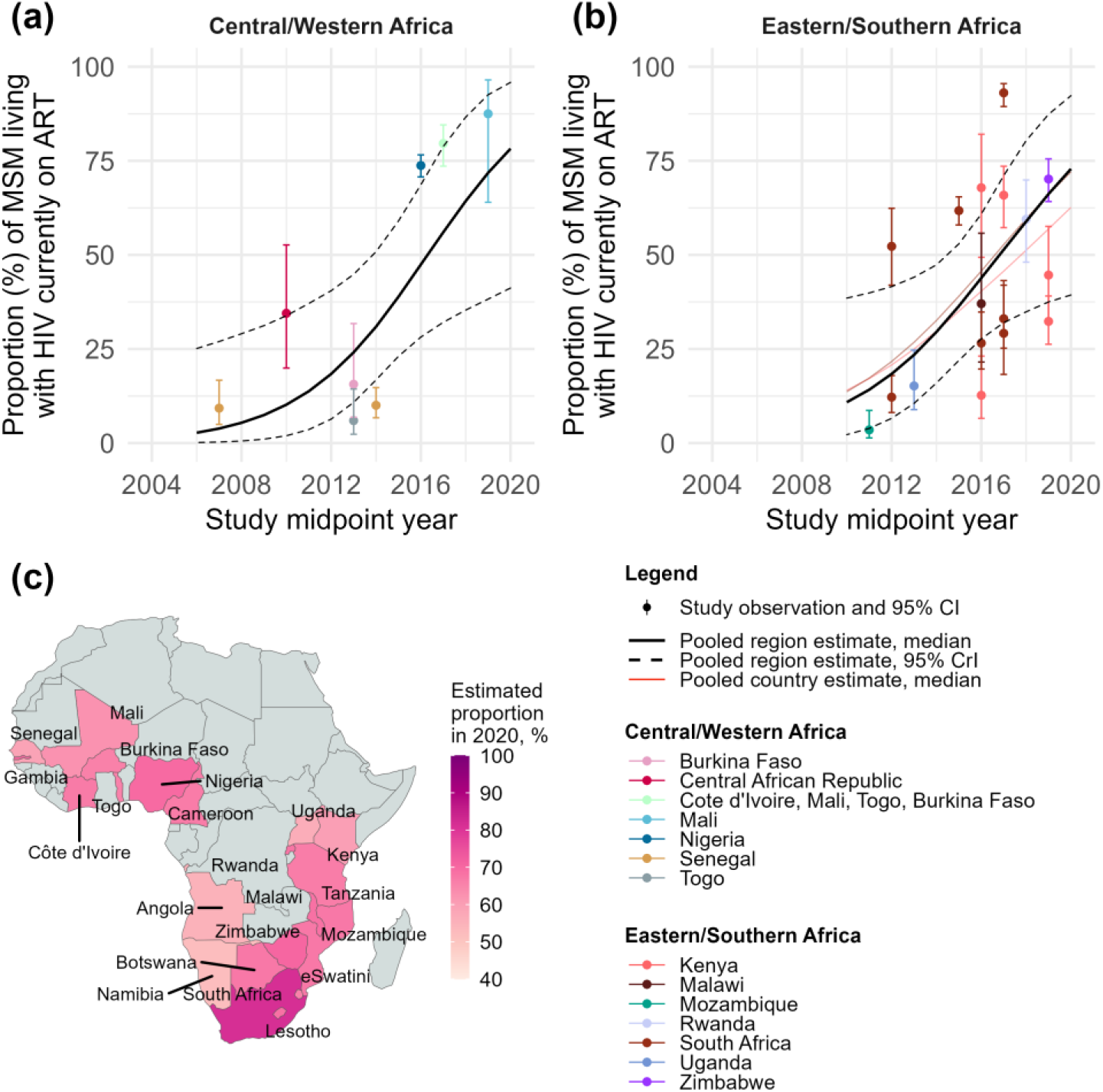
Current antiretroviral therapy (ART) use among men who have sex with men (MSM) living with HIV over time, by region and country of Africa. Current ART use among MSM living with HIV in (a) Central/Western Africa, (b) Eastern/Southern Africa, and (c) the estimated proportion of MSM living with HIV currently on ART in 2020, by country. Points represent available study observations and their 95% confidence intervals, coloured by country in which the study was conducted. The black solid and dotted lines represent the estimated region-level proportions and 95% credible intervals (CrI), respectively. Coloured solid lines represent estimated country-level proportions for countries with at least 3 estimates from 3 different time points (see Figure S19 for individual country-level time trends and 95% CrI).

In 2020, we found that viral suppression was achieved among 62% (95%CrI 12-95%) of MSM living with HIV (Table 2). Time trends in viral suppression among MSM living with HIV suggested potential increases over time, from 11% (1-48%) in 2010 to 61% (32-84%) in 2020 in Eastern/Southern Africa (OR_year_=1.29, 0.95-1.79, N_o_=12), although all credible intervals crossed the null (Figure 5, Table 2, Figure S12, S22). Our 2020 viral suppression estimates were 78% (9-99%) among HIV aware MSM and 91% (24-100%) among MSM currently on ART (Table 2, Figure S13-14, S19, S23-25).

**Figure 5.**
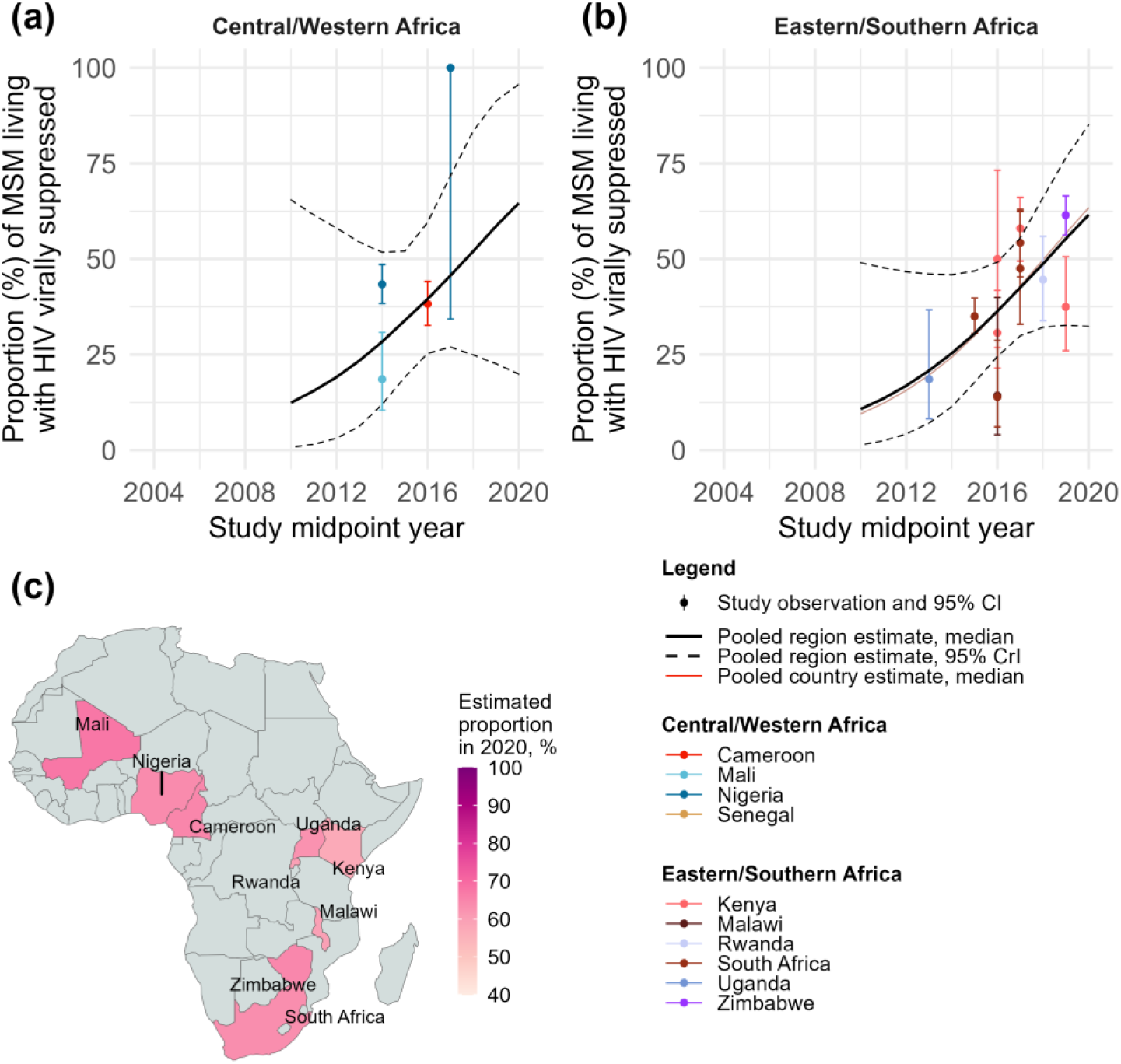
Viral suppression among men who have sex with men (MSM) living with HIV over time, by region and country of Africa. Viral suppression among MSM living with HIV in (a) Central/Western Africa, (b) Eastern/Southern Africa, and (c) the estimated proportion of MSM living with HIV virally suppressed in 2020, by country. Points represent available study observations and their 95% confidence intervals, coloured by country in which the study was conducted. The black solid and dotted lines represent the estimated region-level proportions and 95% credible intervals (CrI), respectively. Coloured solid lines represent estimated country-level proportions for countries with at least 3 estimates from 3 different time points (see Figure S22 for individual country-level time trends and 95% CrI).

In 2020, we estimated that HIV incidence among African MSM was 5.4 per 100 person years (95%CrI 0.9-33.9) and there was no conclusive evidence of a decline in HIV incidence among MSM in Africa over time since 2010 (IRR_year_=0.97, 0.73-1.31, N_o_=15), or in any region (Central/Western Africa: IRR_year_=0.97, 0.82-1.16, N_o_=17; Eastern/Southern Africa: IRR_year_=0.97, 0.82-1.16, N_o_=21; Figure 6, Table 2, Figure S15, S26).

**Figure 6.**
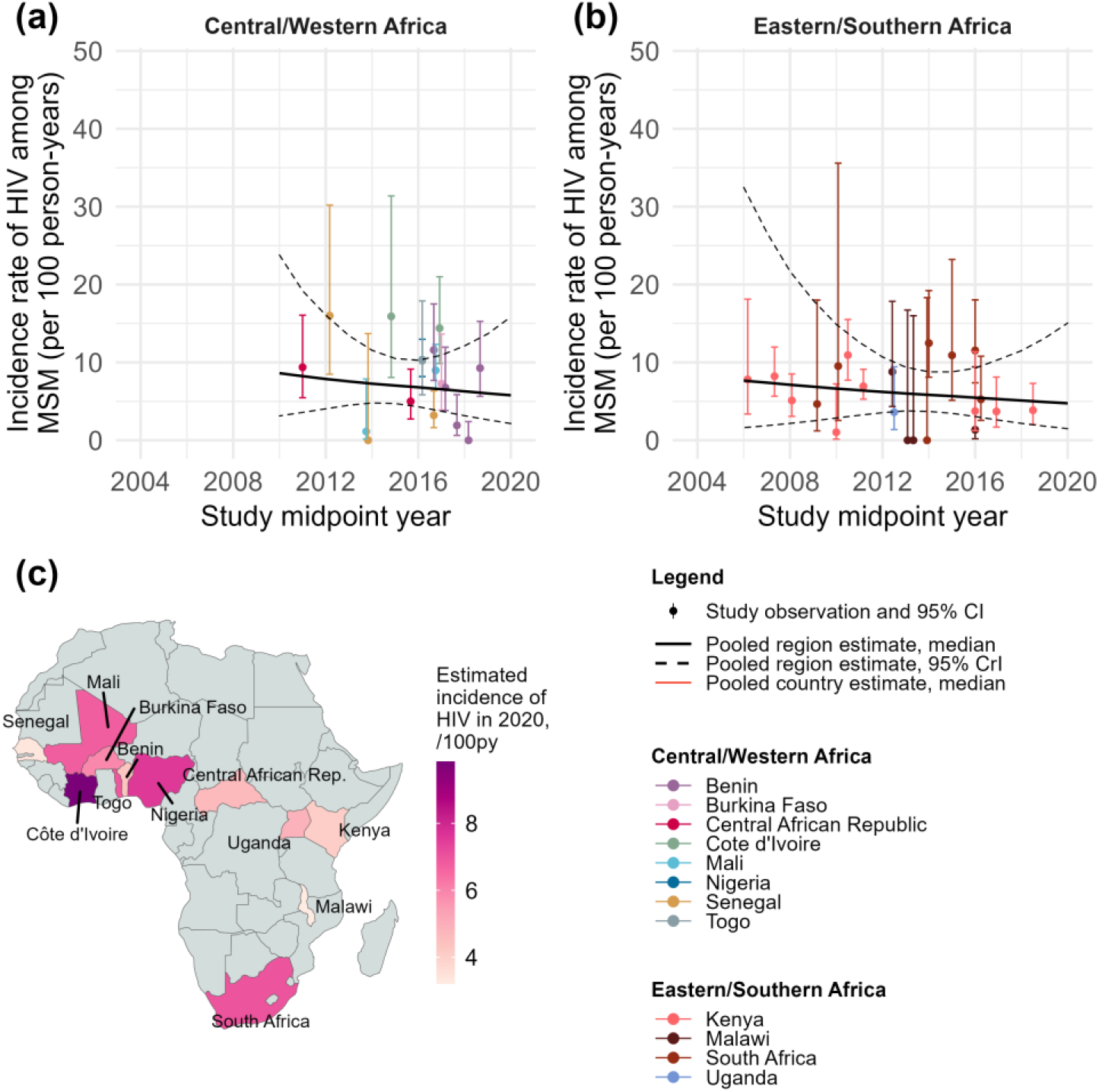
HIV incidence among men who have sex with men (MSM) over time, by region and country of Africa. (a) HIV incidence over time among MSM in Central/Western Africa, (b) Eastern/Southern Africa, and (c) the estimated incidence of HIV among MSM in 2020, by country, estimated using a Bayesian Poisson generalized linear mixed-effects model, with study-, country-, and region-level random effects. Points represent available study observations and their 95% confidence intervals, coloured by country in which the study was conducted. The black solid and dotted lines represent the estimated region-level proportions and 95% credible intervals (CrI), respectively. Coloured solid lines represent estimated country-level proportions for countries with at least 3 estimates from 3 different time points Points represent available study observations and their 95% confidence intervals, coloured by country in which the study was conducted. The black solid and dotted lines represent the estimated region-level HIV incidence rate and 95% credible intervals (CrI), respectively. Coloured solid lines represent estimated country-level HIV incidence rates for countries with at least 3 estimates from 3 different time points (see Figure S26 for individual country-level time trends and 95% CrI).

### The HIV treatment cascade and HIV incidence among MSM compared with all men

Knowledge of status among MSM living with HIV between 2015 and 2020 were consistently lower than year-matched UNAIDS estimates among all men living with HIV aged 15+ in Eastern/Southern Africa, yielding a prevalence ratio (PR) of 0.74 (0.39-0.97) in 2020, but were similar in Central/Western Africa (Figure S27a-b). Point estimates of the PR for current ART use and viral suppression varied in direction across region, but the uncertainty intervals were wide and crossed the null (Figure S27c-f).

Our estimates of HIV incidence among MSM were substantially higher than corresponding UNAIDS estimates among all men aged 15-49 (Figure 7). In 2020, UNAIDS reported an HIV incidence among men of 0.04% in Eastern/Southern Africa and 0.20% in Central/Western Africa. This entails that HIV incidence among MSM could be 27 times higher (95%CrI 8-86 times) than among all men in Eastern/Southern Africa, and 150 times higher (95%CrI 57-419) in Central/Western Africa.

**Figure 7.**
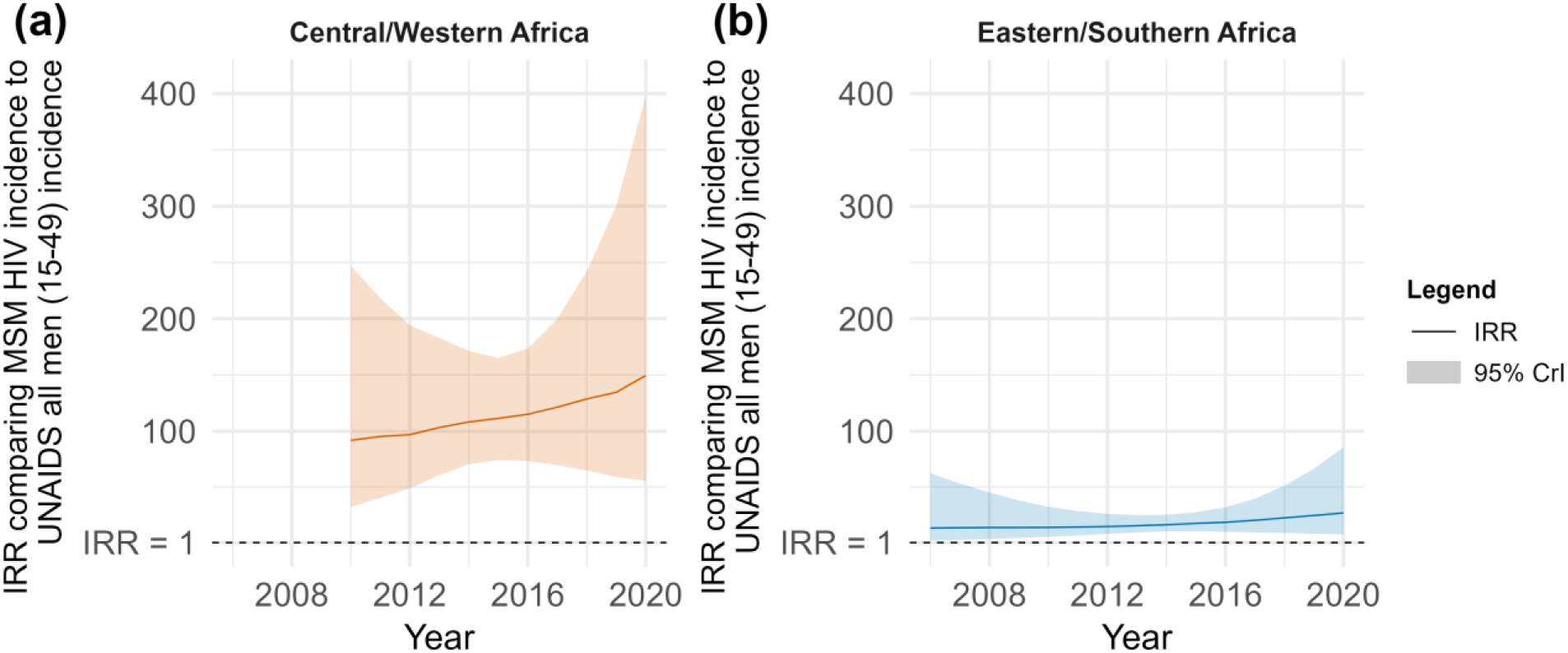
Incidence rate ratios (IRR) and 95% credible intervals (CrI) over time comparing our estimates of HIV incidence among men who have sex with men (MSM) with UNAIDS estimates among all men (aged 15-49), by region of Africa. (a) Central/Western Africa, and (b) Eastern/Southern Africa. Solid lines correspond to medians and shaded areas to the 95% CrI.

### Study quality and risk of bias

Across all studies, risk of bias in reported outcomes was moderate (N_outcomes_=185; Table S3). Study outcomes with a higher risk of bias (N_outcomes_=120) were largely limited by non-representative sampling designs, selected study populations of MSM, and non-confidential interview methods. Funnel plots did not provide strong evidence of publication bias in study observations of HIV incidence, testing, and treatment cascade outcomes, and there was little difference between directly reported study observations and those calculated from available data (Figure S28).

## DISCUSSION

In this comprehensive systematic review and meta-regression study, we highlighted improvements in HIV testing and ART coverage over time among MSM in Africa. Nevertheless, 1 in 3 MSM living with HIV do not have a suppressed viral load. HIV incidence among MSM in Africa was close to 5 per 100 person-years in 2020, and there was no indication of a temporal decline in new HIV acquisition between 2006 and 2020. Such HIV incidence rates among MSM are 27-150 times larger than corresponding rates among all men, highlighting the extreme disparities and exacerbated vulnerabilities to HIV acquisition and transmission among MSM in Africa.

HIV incidence among the overall population has steadily declined over the past decade, by 44% in Eastern/Southern Africa, and 43% in Central/Western Africa. This decline has mainly been attributed to ART scale-up and the resulting population-level viral suppression^181^. As this study suggests, HIV incidence declines among the overall population may not reflect HIV incidence trends among MSM. If fewer resources are allocated to prevention in response to decreasing incidence trends in the overall population, progress among key populations could be compromised^182^. This is especially salient in Central/Western Africa where our 2020 HIV incidence estimate among MSM was 150 times higher than among all men. Even in a hyperendemic context in Eastern/Southern Africa where MSM are estimated to have accounted for only 6% of new HIV acquisitions in 2020, incidence was 27 times higher^1^. In all regions, these disparities are worsening over time as incidence decreases among the general population, despite recent advances in biomedical prevention, including oral and injectable pre-exposure prophylaxis (PrEP), for which access currently remains very limited^1,183,184^. Studies suggest high willingness to use HIV prevention, including PrEP, among MSM in Africa and has potential population-level benefits, through network effects of preventing onward transmissions^185-189^. Yet, services are not available in many countries, or are too far away, too inconvenient, or not adapted to the needs of MSM^190^. These are compounded by economic barriers such as poverty that further limit access^190,191^. Resources to provide services are often limited, efficacious interventions may not be scalable, and services dedicated to MSM may not be appealing if men fear being identified as MSM^190,192^. This study highlights the need for combination HIV prevention, with elements of structural, behavioural, and biomedical interventions. Such an approach is considered the most desirable strategy for attracting and retaining MSM in care and prevention services to achieve reductions in HIV incidence^190^.

Understanding where losses to follow-up occur along the HIV treatment cascade is critical to developing appropriate interventions to reduce HIV transmission and incidence among MSM. We estimated that, in 2020, most MSM had ever tested for HIV (87%) and tested in the past year (88%), and that testing has generally increased over time, particularly in Central/Western Africa, and Eastern Africa, mirroring population-level increases in HIV testing^193^. Nevertheless, only 66% of MSM living with HIV in 2020 were aware of their status. Knowledge of status also remains lower among MSM than among all men living with HIV in Africa. However, knowledge of status may be underestimated since the majority of studies relied on self-reports, which are susceptible to underreporting^194,195^. This is particularly apparent when comparing our knowledge of status estimates with those from current ART coverage, which are roughly 10%-point higher. Going forward, biomarkers could be used to adjust self-reports, but this is only useful in settings where ART coverage is high. More generally, enabling environments are needed that encourage uptake of HIV testing, linkage to care, and disclosure of HIV status. Expanding community-led services, including involving peer-navigators to support MSM to access and remain in care, and increasing the use of alternative, decentralized HIV testing modalities such as HIV self-tests and virtual services could improve knowledge of status and linkage into care for MSM in Africa^196^.

Current ART use has increased over time to reach 78% and 73% of all MSM living with HIV in 2020 in Central/Western Africa and Eastern/Southern Africa, respectively. These coverage estimates are on par with those reported for all men, but slightly higher than those from a recent synthesis of MSM surveys that reported estimates of 52% and 69% in Central/Western Africa and Eastern/Southern Africa in 2021^11^, but the uncertainty intervals in both studies overlap.

Nonetheless, viral suppression among all MSM living with HIV in Africa was lower, at 62%. Our estimates of ART use and viral suppression are lower than what is needed to achieve the 95-95-95 targets, which require that at least 90% of all MSM living with HIV are on ART, and 86% are virally suppressed. Failure to close these gaps leaves MSM vulnerable to ongoing transmission and continued HIV-related morbidity and mortality, undermining the strategy the end AIDS. Innovative drug delivery models, including peer-navigation and provision of ART outside of clinics, could help increase equitable access to first-line ART regimens and increase viral suppression among MSM^197^. Long-acting ART formulations, once availability increases, could also be important for overcoming some barriers to ART adherence.

Our results should be interpreted considering several limitations. First, although we did not exclude studies based on language, we used English and French search terms, which may have missed studies published in other languages. Second, most included studies used non-representative sampling designs, largely relying on convenience sampling, particularly in cohort studies that measured incidence, whilst RDS was common in cross-sectional studies. RDS could oversample young, urban, socially connected MSM, and miss older, non-gay identifying MSM, but can theoretically yield more representative estimates when adjusted for sampling design^198^. However, few of the included studies that used complex sampling designs (including RDS, cluster, and time-location sampling) provided adjusted estimates. Third, variable MSM definitions were applied to recruit participants, and most studies included some transgender women. Fourth, self-reported outcomes were often assessed in face-to-face interviews, which may be impacted by social desirability and recall biases. Increased use of confidential interview methods including audio computer-assisted self-interviews (ACASI) could improve accuracy^199^. Finally, there was high uncertainty in our MSM estimates over time, partly due to the limited number of available studies and heterogeneous observations. There were particularly few observations of engagement in care, ART use, and viral suppression among MSM with minimal increases since previous reviews, and few observations of any outcome from Central or Northern Africa^6^.

Strengths of this study include the substantial increase in the number of included studies compared to previous reviews, using data from 148 studies in 30 countries encompassing over 40,000 MSM, conducted from 2003-2020^6,200,201^. Importantly, we provide novel analyses and results of pooled HIV incidence among MSM over time in Africa. We pooled observations using mixed-effects meta-regression models within a Bayesian framework, which allowed us to borrow information across observations to produce estimates in settings with sparse data. We also calculated additional study estimates, minimizing publication bias.

## CONCLUSIONS

Despite continued increases in HIV testing among and engagement in the HIV treatment cascade among MSM living with HIV across settings, HIV incidence remains high among MSM across Africa and does not appear to be decreasing. Better combination interventions tailored to the primary HIV prevention needs of MSM that address the social, structural, and behavioural factors that exacerbate their vulnerabilities to HIV will likely be important to increase access to ART and viral suppression and, ultimately, reduce disparities in HIV incidence.

## Data Availability

All data produced in the present study are available upon reasonable request to the authors.

## Contributions

JS, MC-B, MM-G, and JL conceptualized this review and planned the analysis. JS, NS, and LJ did the search and independently did all stages of screening. JS, NS, and LJ independently extracted data, and JS conducted all analyses. NS and KG double checked the data extraction. MM-G and MC-B checked the data analysis. JS, MM-G, and MC-B interpreted the results and conceptualized the first draft of the review. KM, NK, RM, MNN, and GMK made substantial contributions to the interpretation of the results and edited the manuscript. All authors read and approved the final version of the manuscript.

## REFERENCES

1. UNAIDS. IN DANGER: UNAIDS Global AIDS Update 2022, 2022.

2. ILGA World:, Lucas Ramon Mendos, Kellyn Botha, et al. State-Sponsored Homophobia 2020: Global Legislation Overview Update. Geneva, Switzerland: ILGA, 2020.

3. Wiginton JM, Murray SM, Poku O, et al. Disclosure of same-sex practices and experiences of healthcare stigma among cisgender men who have sex with men in five sub-Saharan African countries. BMC Public Health 2021; 21(1): 2206.

4. Zahn R, Grosso A, Scheibe A, et al. Human Rights Violations among Men Who Have Sex with Men in Southern Africa: Comparisons between Legal Contexts. PLoS One 2016; 11(1): e0147156.

5. UNAIDS. Global AIDS Strategy 2021-2026 — End Inequalities. End AIDS. Geneva, Switzerland, 2021.

6. Stannah J, Dale E, Elmes J, et al. HIV testing and engagement with the HIV treatment cascade among men who have sex with men in Africa: a systematic review and meta-analysis. Lancet HIV 2019; 6(11): e769–e87.

7. Moher D, Altman DG, Liberati A, Tetzlaff J. PRISMA statement. Epidemiology 2011; 22(1): 128.

8. Maheu-Giroux M, Sardinha L, Stöckl H, et al. A framework to model global, regional, and national estimates of intimate partner violence. BMC Med Res Methodol 2022; 22(1): 159.

9. WHO, UNAIDS. Key populations strategic information: Recommended population size estimates of men who have sex with men. Geneva, 2020.

10. Baral S, Turner RM, Lyons CE, et al. Population Size Estimation of Gay and Bisexual Men and Other Men Who Have Sex With Men Using Social Media-Based Platforms. JMIR Public Health Surveill 2018; 4(1): e15.

11. Stevens O, Sabin K, Garcia SA, et al. Key population size, HIV prevalence, and ART coverage in sub-Saharan Africa: systematic collation and synthesis of survey data. medRxiv 2022: 2022.07.27.22278071.

12. Stan Development Team. Stan Modeling Language Users Guide and Reference Manual, Version 2.30. 2022.

13. AIDSinfo. UNAIDS 2020 Estimates. 2020.

14. Bürkner P-C. brms: An R Package for Bayesian Multilevel Models Using Stan. Journal of Statistical Software 2017; 80(1): 1 – 28.

15. Team SD. RStan: the R interface to Stan. R package version 2016; 2(1): 522.

16. Ramadhani HO, Crowell TA, Nowak RG, et al. Association of age with healthcare needs and engagement among Nigerian men who have sex with men and transgender women: cross-sectional and longitudinal analyses from an observational cohort. Journal of the International AIDS Society 2020; 23: e25599.

17. Lyons CE, Olawore O, Turpin G, et al. Intersectional stigmas and HIV-related outcomes among a cohort of key populations enrolled in stigma mitigation interventions in Senegal. AIDS (London, England) 2020; 34(Suppl 1): S63.

18. Dramé FM, Crawford EE, Diouf D, Beyrer C, Baral SD. A pilot cohort study to assess the feasibility of HIV prevention science research among men who have sex with men in Dakar, Senegal. Journal of the International AIDS Society 2013; 16: 18753.

19. Lippman SA, Lane T, Rabede O, et al. High acceptability and increased HIV testing frequency following introduction of HIV self-testing and network distribution among South African MSM. Journal of acquired immune deficiency syndromes (1999) 2018; 77(3): 279.

20. McKinnon LR, Gakii G, Juno JA, et al. High HIV risk in a cohort of male sex workers from Nairobi, Kenya. Sexually transmitted infections 2014; 90(3): 237–42.

21. Kamali A, Price MA, Lakhi S, et al. Creating an African HIV clinical research and prevention trials network: HIV prevalence, incidence and transmission. PloS one 2015; 10(1): e0116100.

22. Wirtz AL, Trapence G, Jumbe V, et al. Feasibility of a combination HIV prevention program for men who have sex with men in Blantyre, Malawi. Journal of acquired immune deficiency syndromes (1999) 2015; 70(2): 155.

23. Sandfort TG, Mbilizi Y, Sanders EJ, et al. HIV incidence in a multinational cohort of men and transgender women who have sex with men in sub-Saharan Africa: Findings from HPTN 075. PloS one 2021; 16(2): e0247195.

24. Buchbinder SP, Glidden DV, Liu AY, et al. Who should be offered HIV pre-exposure prophylaxis (PrEP)?: A secondary analysis of a Phase 3 PrEP efficacy trial in men who have sex with men and transgender women. The Lancet infectious diseases 2014; 14(6): 468.

25. Dah TTE, Yaya I, Sagaon-Teyssier L, et al. Adherence to quarterly HIV prevention services and its impact on HIV incidence in men who have sex with men in West Africa (CohMSM ANRS 12324–Expertise France). BMC Public Health 2021; 21(1): 1–13.

26. Hessou SP, Glele-Ahanhanzo Y, Adekpedjou R, et al. HIV incidence and risk contributing factors among men who have sex with men in Benin: A prospective cohort study. PloS one 2020; 15(6): e0233624.

27. Kimani M, van der Elst EM, Chiro O, et al. Pr EP interest and HIV-1 incidence among MSM and transgender women in coastal Kenya. Journal of the International AIDS Society 2019; 22(6): e25323.

28. Maenetje P, Lindan C, Makkan H, et al. HIV incidence and predictors of inconsistent condom use among adult men enrolled into an HIV vaccine preparedness study, Rustenburg, South Africa. PloS one 2019; 14(4): e0214786.

29. Mbeko Simaleko M, Longo JDD, Magloire C-PS, et al. Persistent high-risk behavior and escalating HIV, syphilis and hepatitis B incidences among men who have sex with men living in Bangui, Central African Republic. Pan African Medical Journal 2018; 29(1): 1–11.

30. Sullivan PS, Phaswana-Mafuya N, Baral SD, et al. HIV prevalence and incidence in a cohort of South African men and transgender women who have sex with men: the Sibanye Methods for Prevention Packages Programme (MP3) project. Journal of the International AIDS Society 2020; 23: e25591.

31. Mbeko Simaleko M, Camengo Police SM, Longo JDD, Piette D, Humblet CP, Gresenguet G. Efficacité de la Combinaison d’Interventions de Prévention chez les Hommes Ayant des Rapports Sexuels avec des Hommes à Bangui (République Centrafricaine). Health sciences and disease 2020; 21(7): 94–9.

32. Wahome EW, Graham SM, Thiong’o AN, et al. PrEP uptake and adherence in relation to HIV-1 incidence among Kenyan men who have sex with men. EClinicalMedicine 2020; 26: 100541.

33. Couderc C, Keita BD, Anoma C, et al. Is PrEP needed for MSM in West Africa? HIV incidence in a prospective multicountry cohort. JAIDS Journal of Acquired Immune Deficiency Syndromes 2017; 75(3): e80–e2.

34. Robb ML, Eller LA, Kibuuka H, et al. Prospective study of acute HIV-1 infection in adults in East Africa and Thailand. New England Journal of Medicine 2016; 374(22): 2120–30.

35. Mdodo R, Gust D, Otieno FO, et al. Investigation of HIV incidence rates in a high-risk, high-prevalence Kenyan population: potential lessons for intervention trials and programmatic strategies. Journal of the International Association of Providers of AIDS Care (JIAPAC) 2016; 15(1): 42–50.

36. Lane T, Osmand T, Marr A, Struthers H, McIntyre JA, Shade SB. Brief Report: High HIV Incidence in a South African Community of Men Who Have Sex With Men: Results From the Mpumalanga Men’s Study, 2012–2015. JAIDS Journal of Acquired Immune Deficiency Syndromes 2016; 73(5): 609–11.

37. Holland CE, Papworth E, Billong SC, et al. Access to HIV services at non-governmental and community-based organizations among men who have sex with men (MSM) in Cameroon: An integrated biological and behavioral surveillance analysis. PLoS One 2015; 10(4): e0122881.

38. Lorente N, Henry E, Fugon L, et al. Proximity to HIV is associated with a high rate of HIV testing among men who have sex with men living in Douala, Cameroon. AIDS care 2012; 24(8): 1020–7.

39. Goodman SH, Grosso AL, Ketende SC, et al. Examining the correlates of sexually transmitted infection testing among men who have sex with men in Ouagadougou and Bobo-Dioulasso, Burkina Faso. Sexually transmitted diseases 2016; 43(5): 302–9.

40. Hakim AJ, Aho J, Semde G, et al. The epidemiology of HIV and prevention needs of men who have sex with men in Abidjan, Cote d’Ivoire. PloS one 2015; 10(4): e0125218.

41. Girault P, Green K, Clement NF, Rahman YAA, Adams B, Wambugu S. Piloting a social networks strategy to increase HIV testing and counseling among men who have sex with men in Greater Accra and Ashanti Region, Ghana. AIDS and Behavior 2015; 19(11): 1990–2000.

42. Nelson LE, Wilton L, Agyarko-Poku T, et al. The association of HIV stigma and HIV/STD knowledge with sexual risk behaviors among adolescent and adult men who have sex with men in Ghana, West Africa. Research in nursing & health 2015; 38(3): 194–206.

43. Lahuerta M, Patnaik P, Ballo T, et al. HIV prevalence and related risk factors in men who have sex with men in Bamako, Mali: findings from a bio-behavioral survey using respondent-driven sampling. AIDS and Behavior 2018; 22(7): 2079–88.

44. Eluwa GI, Adebajo SB, Eluwa T, Ogbanufe O, Ilesanmi O, Nzelu C. Rising HIV prevalence among men who have sex with men in Nigeria: a trend analysis. BMC Public Health 2019; 19(1): 1–10.

45. Adebajo S, Obianwu O, Eluwa G, et al. Comparison of audio computer assisted self-interview and face-to-face interview methods in eliciting HIV-related risks among men who have sex with men and men who inject drugs in Nigeria. PloS one 2014; 9(1): e81981.

46. Strömdahl S, Onigbanjo Williams A, Eziefule B, et al. An assessment of stigma and human right violations among men who have sex with men in Abuja, Nigeria. BMC International Health and Human Rights 2019; 19(1): 1–7.

47. Wade AS, Kane CT, Diallo PAN, et al. HIV infection and sexually transmitted infections among men who have sex with men in Senegal. Aids 2005; 19(18): 2133–40.

48. Ruiseñor-Escudero H, Lyons C, Ketende S, et al. Prevalence and factors associated to disclosure of same-sex practices to family members and health care workers among men who have sex with men in Togo. AIDS care 2019.

49. Bakai TA, Ekouevi DK, Tchounga BK, et al. Condom use and associated factors among men who have sex with men in Togo, West Africa. Pan African Medical Journal 2016; 23(1).

50. Kendall C, Kerr LRFS, Mota RMS, et al. Population size, HIV, and behavior among MSM in Luanda, Angola: challenges and findings in the first ever HIV and syphilis biological and behavioral survey. Journal of acquired immune deficiency syndromes (1999) 2014; 66(5): 544.

51. Stahlman S, Grosso A, Ketende S, et al. Characteristics of men who have sex with men in southern Africa who seek sex online: a cross-sectional study. Journal of medical Internet research 2015; 17(5): e4230.

52. Batist E, Brown B, Scheibe A, Baral SD, Bekker LG. Outcomes of a community-based HIV-prevention pilot programme for township men who have sex with men in Cape Town, South Africa. Journal of the International AIDS Society 2013; 16: 18754.

53. Siegler AJ, Sullivan PS, De Voux A, et al. Exploring repeat HIV testing among men who have sex with men in Cape Town and Port Elizabeth, South Africa. AIDS care 2015; 27(2): 229–34.

54. Wagenaar BH, Sullivan PS, Stephenson R. HIV knowledge and associated factors among internet-using men who have sex with men (MSM) in South Africa and the United States. PloS one 2012; 7(3): e32915.

55. Eaton LA, Pitpitan EV, Kalichman SC, et al. Men who report recent male and female sex partners in Cape Town, South Africa: an understudied and underserved population. Archives of Sexual Behavior 2013; 42(7): 1299–308.

56. Tun W, Kellerman S, Maimane S, et al. HIV-related conspiracy beliefs and its relationships with HIV testing and unprotected sex among men who have sex with men in Tshwane (Pretoria), South Africa. AIDS care 2012; 24(4): 459–67.

57. Knox J, Sandfort T, Yi H, Reddy V, Maimane S. Social vulnerability and HIV testing among South African men who have sex with men. International journal of STD & AIDS 2011; 22(12): 709–13.

58. Lane T, Raymond HF, Dladla S, et al. High HIV prevalence among men who have sex with men in Soweto, South Africa: results from the Soweto Men’s Study. AIDS and Behavior 2011; 15(3): 626–34.

59. Lane T, Shade SB, McIntyre J, Morin SF. Alcohol and sexual risk behavior among men who have sex with men in South African township communities. AIDS and Behavior 2008; 12(1): 78–85.

60. Nel JA, Yi H, Sandfort TG, Rich E. HIV-untested men who have sex with men in South Africa: the perception of not being at risk and fear of being tested. AIDS and Behavior 2013; 17(1): 51–9.

61. Coulaud P-j, Mujimbere G, Nitunga A, et al. An assessment of health interventions required to prevent the transmission of HIV infection among men having sex with men in Bujumbura, Burundi. Journal of community health 2016; 41(5): 1033–43.

62. Bhattacharjee P, McClarty LM, Musyoki H, et al. Monitoring HIV prevention programme outcomes among key populations in Kenya: findings from a national survey. PLoS One 2015; 10(8): e0137007.

63. Muraguri N, Tun W, Okal J, et al. HIV and STI prevalence and risk factors among male sex workers and other men who have sex with men in Nairobi, Kenya. Journal of acquired immune deficiency syndromes (1999) 2015; 68(1): 91.

64. Luchters S, Geibel S, Syengo M, et al. Use of AUDIT, and measures of drinking frequency and patterns to detect associations between alcohol and sexual behaviour in male sex workers in Kenya. BMC public health 2011; 11(1): 1–8.

65. Möller LM, Stolte IG, Geskus RB, et al. Changes in sexual risk behavior among MSM participating in a research cohort in coastal Kenya. AIDS (London, England) 2015; 29(0 3): S211.

66. Sanders EJ, Graham SM, Okuku HS, et al. HIV-1 infection in high risk men who have sex with men in Mombasa, Kenya. Aids 2007; 21(18): 2513–20.

67. Wirtz AL, Trapence G, Kamba D, et al. Geographical disparities in HIV prevalence and care among men who have sex with men in Malawi: results from a multisite cross-sectional survey. The Lancet HIV 2017; 4(6): e260–e9.

68. Ntata PRT, Muula AS, Siziya S. Socio-demographic characteristics and sexual health related attitudes and practices of men having sex with men in central and southern Malawi. Tanzania Journal of Health Research 2008; 10(3): 124–30.

69. Boothe MAS, Semá Baltazar C, Sathane I, et al. Young key populations left behind: The necessity for a targeted response in Mozambique. PLoS One 2021; 16(12): e0261943.

70. Chapman J, Koleros A, Delmont Y, Pegurri E, Gahire R, Binagwaho A. High HIV risk behavior among men who have sex with men in Kigali, Rwanda: making the case for supportive prevention policy. AIDS Care 2011; 23(4): 449–55.

71. Nyoni JE, Ross MW. Condom use and HIV-related behaviors in urban Tanzanian men who have sex with men: a study of beliefs, HIV knowledge sources, partner interactions and risk behaviors. AIDS Care 2013; 25(2): 223–9.

72. Romijnders KA, Nyoni JE, Ross MW, et al. Lubricant use and condom use during anal sex in men who have sex with men in Tanzania. Int J STD AIDS 2016; 27(14): 1289–302.

73. Khatib A, Haji S, Khamis M, et al. Reproducibility of Respondent-Driven Sampling (RDS) in Repeat Surveys of Men Who have Sex with Men, Unguja, Zanzibar. AIDS Behav 2017; 21(7): 2180–7.

74. Wanyenze RK, Musinguzi G, Matovu JK, et al. “If You Tell People That You Had Sex with a Fellow Man, It Is Hard to Be Helped and Treated”: Barriers and Opportunities for Increasing Access to HIV Services among Men Who Have Sex with Men in Uganda. PLoS One 2016; 11(1): e0147714.

75. Hladik W, Barker J, Ssenkusu JM, et al. HIV infection among men who have sex with men in Kampala, Uganda--a respondent driven sampling survey. PLoS One 2012; 7(5): e38143.

76. Hladik W, Sande E, Berry M, et al. Men Who Have Sex with Men in Kampala, Uganda: Results from a Bio-Behavioral Respondent Driven Sampling Survey. AIDS Behav 2017; 21(5): 1478–90.

77. Kajubi P, Kamya MR, Raymond HF, et al. Gay and bisexual men in Kampala, Uganda. AIDS Behav 2008; 12(3): 492–504.

78. Fay H, Baral SD, Trapence G, et al. Stigma, health care access, and HIV knowledge among men who have sex with men in Malawi, Namibia, and Botswana. AIDS Behav 2011; 15(6): 1088–97.

79. Longo JDD, Diemer HSC, Tonen-Wolyec S, et al. HIV self-testing in Central Africa: stakes and challenges. Health sciences and disease 2018; 19(2).

80. Elmahy AG. Reaching Egyptian Gays Using Social Media: A Comprehensive Health Study and a Framework for Future Research. J Homosex 2018; 65(13): 1867–76.

81. Herce ME, Miller WM, Bula A, et al. Achieving the first 90 for key populations in sub-Saharan Africa through venue-based outreach: challenges and opportunities for HIV prevention based on PLACE study findings from Malawi and Angola. J Int AIDS Soc 2018; 21 Suppl 5(Suppl Suppl 5): e25132.

82. Lieber M, Reynolds CW, Lieb W, McGill S, Beddoe AM. Human Papillomavirus Knowledge, Attitudes, Practices, and Prevalence Among Men Who Have Sex With Men in Monrovia, Liberia. J Low Genit Tract Dis 2018; 22(4): 326–32.

83. Mmbaga EJ, Moen K, Leyna GH, Mpembeni R, Leshabari MT. HIV Prevalence and Associated Risk Factors Among Men Who Have Sex With Men in Dar es Salaam, Tanzania. J Acquir Immune Defic Syndr 2018; 77(3): 243–9.

84. Teclessou JN, Akakpo SA, Ekouevi KD, Koumagnanou G, Singo-Tokofai A, Pitche PV. Evolution of HIV prevalence and behavioral factors among MSM in Togo between 2011 and 2015. Pan Afr Med J 2017; 28: 191.

85. Tun W, Vu L, Dirisu O, et al. Uptake of HIV self-testing and linkage to treatment among men who have sex with men (MSM) in Nigeria: A pilot programme using key opinion leaders to reach MSM. J Int AIDS Soc 2018; 21 Suppl 5(Suppl Suppl 5): e25124.

86. Sandfort TGM, Dominguez K, Kayange N, et al. HIV testing and the HIV care continuum among sub-Saharan African men who have sex with men and transgender women screened for participation in HPTN 075. PLoS One 2019; 14(5): e0217501.

87. Tobin-West C, Nwajagu S, Maduka O, Oranu E, Onyekwere V, Tamuno I. Exploring the HIV-risk practices of men who have sex with men in Port Harcourt city, Nigeria. Annals of Tropical Medicine and Public Health 2017; 10(3).

88. Nyblade L, Reddy A, Mbote D, et al. The relationship between health worker stigma and uptake of HIV counseling and testing and utilization of non-HIV health services: the experience of male and female sex workers in Kenya. AIDS Care 2017; 29(11): 1364–72.

89. Sadio AJ, Gbeasor-Komlanvi FA, Konu YR, et al. Prevalence of HIV infection and hepatitis B and factors associated with them among men who had sex with men in Togo in 2017. Med Sante Trop 2019; 29(3): 294–301.

90. Jobson G, Tucker A, de Swardt G, et al. Gender identity and HIV risk among men who have sex with men in Cape Town, South Africa. AIDS Care 2018; 30(11): 1421–5.

91. Maleke K, Makhakhe N, Peters RP, et al. HIV risk and prevention among men who have sex with men in rural South Africa. Afr J AIDS Res 2017; 16(1): 31–8.

92. Ulanja MB, Lyons C, Ketende S, et al. The relationship between depression and sexual health service utilization among men who have sex with men (MSM) in Côte d’Ivoire, West Africa. BMC Int Health Hum Rights 2019; 19(1): 11.

93. Okoboi S, Lazarus O, Castelnuovo B, et al. Peer distribution of HIV self-test kits to men who have sex with men to identify undiagnosed HIV infection in Uganda: A pilot study. PLoS One 2020; 15(1): e0227741.

94. Yaya I, Diallo F, Kouamé MJ, et al. Decrease in incidence of sexually transmitted infections symptoms in men who have sex with men enrolled in a quarterly HIV prevention and care programme in West Africa (CohMSM ANRS 12324-Expertise France). Sex Transm Infect 2022; 98(2): 85–94.

95. Magesa DJ, Mtui LJ, Abdul M, et al. Barriers to men who have sex with men attending HIV related health services in Dar es Salaam, Tanzania. Tanzan J Health Res 2014; 16(2): 118–26.

96. Lillie T, Boyee D, Kamariza G, Nkunzimana A, Gashobotse D, Persaud N. Increasing Testing Options for Key Populations in Burundi Through Peer-Assisted HIV Self-Testing: Descriptive Analysis of Routine Programmatic Data. JMIR Public Health Surveill 2021; 7(9): e24272.

97. Vuylsteke B, Semde G, Sika L, et al. High prevalence of HIV and sexually transmitted infections among male sex workers in Abidjan, Cote d’Ivoire: need for services tailored to their needs. Sex Transm Infect 2012; 88(4): 288–93.

98. Inghels M, Kouassi AK, Niangoran S, et al. Telephone peer recruitment and interviewing during a respondent-driven sampling (RDS) survey: feasibility and field experience from the first phone-based RDS survey among men who have sex with men in Côte d’Ivoire. BMC Med Res Methodol 2021; 21(1): 25.

99. Abubakari GM, Nelson LE, Ogunbajo A, et al. Implementation and evaluation of a culturally grounded group-based HIV prevention programme for men who have sex with men in Ghana. Glob Public Health 2021; 16(7): 1028–45.

100. Gyamerah AO, Taylor KD, Atuahene K, et al. Stigma, discrimination, violence, and HIV testing among men who have sex with men in four major cities in Ghana. AIDS Care 2020; 32(8): 1036–44.

101. Ekouevi DK, Dagnra CY, Goilibe KB, et al. HIV seroprevalence and associated factors among men who have sex with men in Togo. Rev Epidemiol Sante Publique 2014; 62(2): 127–34.

102. Parmley LE, Harris TG, Chingombe I, et al. Engagement in the pre-exposure prophylaxis (PrEP) cascade among a respondent-driven sample of sexually active men who have sex with men and transgender women during early PrEP implementation in Zimbabwe. J Int AIDS Soc 2022; 25(2): e25873.

103. Ross MW, Kashiha J, Nyoni J, Larsson M, Agardh A. Electronic Media Access and Use for Sexuality and Sexual Health Education Among Men Who Have Sex With Men in Four Cities in Tanzania. International Journal of Sexual Health 2018; 30(3): 264–70.

104. Mmbaga EJ, Dodo MJ, Leyna GH, Moen K, Leshabari MT. Sexual practices and perceived susceptibility to HIV infection among men who have sex with men in Dar Es Salaam, mainland Tanzania. Journal of AIDS and Clinical Research 2012; 2012: 1–6.

105. Twahirwa Rwema JO, Lyons CE, Herbst S, et al. HIV infection and engagement in HIV care cascade among men who have sex with men and transgender women in Kigali, Rwanda: a cross-sectional study. J Int AIDS Soc 2020; 23 Suppl 6(Suppl 6): e25604.

106. Pillay D, Stankevitz K, Lanham M, et al. Factors influencing uptake, continuation, and discontinuation of oral PrEP among clients at sex worker and MSM facilities in South Africa. PLoS One 2020; 15(4): e0228620.

107. Scheibe A, Young K, Versfeld A, et al. Hepatitis B, hepatitis C and HIV prevalence and related sexual and substance use risk practices among key populations who access HIV prevention, treatment and related services in South Africa: findings from a seven-city cross-sectional survey (2017). BMC Infect Dis 2020; 20(1): 655.

108. Fearon E, Tenza S, Mokoena C, et al. HIV testing, care and viral suppression among men who have sex with men and transgender individuals in Johannesburg, South Africa. PLoS One 2020; 15(6): e0234384.

109. Montgomery ET, Browne EN, Atujuna M, et al. Long-Acting Injection and Implant Preferences and Trade-Offs for HIV Prevention Among South African Male Youth. J Acquir Immune Defic Syndr 2021; 87(3): 928–36.

110. Stephenson R, Darbes LA, Chavanduka T, Essack Z, van Rooyen H. HIV Testing, Knowledge and Willingness to Use PrEP Among Partnered Men Who Have Sex With Men in South Africa and Namibia. AIDS Behav 2021; 25(7): 1993–2004.

111. Smith AD, Kimani J, Kabuti R, Weatherburn P, Fearon E, Bourne A. HIV burden and correlates of infection among transfeminine people and cisgender men who have sex with men in Nairobi, Kenya: an observational study. Lancet HIV 2021; 8(5): e274–e83.

112. Bhattacharjee P, Isac S, Musyoki H, et al. HIV prevalence, testing and treatment among men who have sex with men through engagement in virtual sexual networks in Kenya: a cross-sectional bio-behavioural study. J Int AIDS Soc 2020; 23 Suppl 2(Suppl 2): e25516.

113. Dijkstra M, Mohamed K, Kigoro A, et al. Peer Mobilization and Human Immunodeficiency Virus (HIV) Partner Notification Services Among Gay, Bisexual, and Other Men Who Have Sex With Men and Transgender Women in Coastal Kenya Identified a High Number of Undiagnosed HIV Infections. Open Forum Infect Dis 2021; 8(6): ofab219.

114. Githuka G, Hladik W, Mwalili S, et al. Populations at increased risk for HIV infection in Kenya: results from a national population-based household survey, 2012. J Acquir Immune Defic Syndr 2014; 66 Suppl 1(Suppl 1): S46–56.

115. Lane T, Osmand T, Marr A, et al. The Mpumalanga Men’s Study (MPMS): results of a baseline biological and behavioral HIV surveillance survey in two MSM communities in South Africa. PLoS One 2014; 9(11): e111063.

116. Park JN, Papworth E, Billong SC, et al. Correlates of prior HIV testing among men who have sex with men in Cameroon: a cross-sectional analysis. BMC Public Health 2014; 14: 1220.

117. Bouscaillou J, Evanno J, Prouté M, et al. Prevalence and risk factors associated with HIV and tuberculosis in people who use drugs in Abidjan, Ivory Coast. Int J Drug Policy 2016; 30: 116–23.

118. Aho J, Hakim A, Vuylsteke B, et al. Exploring risk behaviors and vulnerability for HIV among men who have sex with men in Abidjan, Cote d’Ivoire: poor knowledge, homophobia and sexual violence. PLoS One 2014; 9(6): e99591.

119. Adam PC, de Wit JB, Toskin I, et al. Estimating levels of HIV testing, HIV prevention coverage, HIV knowledge, and condom use among men who have sex with men (MSM) in low-income and middle-income countries. J Acquir Immune Defic Syndr 2009; 52 Suppl 2: S143–51.

120. Baral S, Adams D, Lebona J, et al. A cross-sectional assessment of population demographics, HIV risks and human rights contexts among men who have sex with men in Lesotho. J Int AIDS Soc 2011; 14: 36.

121. Rebe K, Lewis D, Myer L, et al. A Cross Sectional Analysis of Gonococcal and Chlamydial Infections among Men-Who-Have-Sex-with-Men in Cape Town, South Africa. PLoS One 2015; 10(9): e0138315.

122. Rao A, Stahlman S, Hargreaves J, et al. Sampling Key Populations for HIV Surveillance: Results From Eight Cross-Sectional Studies Using Respondent-Driven Sampling and Venue-Based Snowball Sampling. JMIR Public Health Surveill 2017; 3(4): e72.

123. Valadez JJ, Berendes S, Jeffery C, et al. Filling the Knowledge Gap: Measuring HIV Prevalence and Risk Factors among Men Who Have Sex with Men and Female Sex Workers in Tripoli, Libya. PLoS One 2013; 8(6): e66701.

124. Shangani S, Naanyu V, Mwangi A, et al. Factors associated with HIV testing among men who have sex with men in Western Kenya: a cross-sectional study. Int J STD AIDS 2017; 28(2): 179–87.

125. Bowring AL, Ketende S, Rao A, et al. Characterising unmet HIV prevention and treatment needs among young female sex workers and young men who have sex with men in Cameroon: a cross-sectional analysis. Lancet Child Adolesc Health 2019; 3(7): 482–91.

126. Ntale RS, Rutayisire G, Mujyarugamba P, et al. HIV seroprevalence, self-reported STIs and associated risk factors among men who have sex with men: a cross-sectional study in Rwanda, 2015. Sex Transm Infect 2019; 95(1): 71–4.

127. Ahouada C, Diabaté S, Mondor M, et al. Acceptability of pre-exposure prophylaxis for HIV prevention: facilitators, barriers and impact on sexual risk behaviors among men who have sex with men in Benin. BMC Public Health 2020; 20(1): 1267.

128. Virkud AV, Arimi P, Ssengooba F, et al. Access to HIV prevention services in East African cross-border areas: a 2016-2017 cross-sectional bio-behavioural study. J Int AIDS Soc 2020; 23 Suppl 3(Suppl 3): e25523.

129. Diabaté S, Kra O, Biékoua YJ, et al. Pre-exposure prophylaxis among men who have sex with men in Côte d’Ivoire: a quantitative study of acceptability. AIDS Care 2021; 33(9): 1228–36.

130. Burrell E, Mark D, Grant R, Wood R, Bekker LG. Sexual risk behaviours and HIV-1 prevalence among urban men who have sex with men in Cape Town, South Africa. Sex Health 2010; 7(2): 149–53.

131. Stephenson R, Darbes LA, Chavanduka T, Essack Z, van Rooyen H. Intimate Partner Violence among Male Couples in South Africa and Namibia. Journal of Family Violence 2022; 37(3): 395–405.

132. Raymond HF, Kajubi P, Kamya MR, Rutherford GW, Mandel JS, McFarland W. Correlates of unprotected receptive anal intercourse among gay and bisexual men: Kampala, Uganda. AIDS Behav 2009; 13(4): 677–81.

133. Moran A, Scheim A, Lyons C, et al. Characterizing social cohesion and gender identity as risk determinants of HIV among cisgender men who have sex with men and transgender women in Côte d’Ivoire. Ann Epidemiol 2020; 42: 25–32.

134. Russell C, Tahlil K, Davis M, et al. Barriers to condom use among key populations in Namibia. Int J STD AIDS 2019; 30(14): 1417–24.

135. Knox J, Reddy V, Lane T, Lovasi GS, Hasin D, Sandfort T. Safer sex intentions modify the relationship between substance use and sexual risk behavior among black South African men who have sex with men. Int J STD AIDS 2019; 30(8): 786–94.

136. Smith AD, Fearon E, Kabuti R, et al. Disparities in HIV/STI burden and care coverage among men and transgender persons who have sex with men in Nairobi, Kenya: a cross-sectional study. BMJ Open 2021; 11(12): e055783.

137. Fearon E, Bourne A, Tenza S, et al. Online socializing among men who have sex with men and transgender people in Nairobi and Johannesburg and implications for public health-related research and health promotion: an analysis of qualitative and respondent-driven sampling survey data. J Int AIDS Soc 2020; 23 Suppl 6(Suppl 6): e25603.

138. Holland CE, Kouanda S, Lougué M, et al. Using Population-Size Estimation and Cross-sectional Survey Methods to Evaluate HIV Service Coverage Among Key Populations in Burkina Faso and Togo. Public Health Rep 2016; 131(6): 773–82.

139. Hakim A, Patnaik P, Telly N, et al. High Prevalence of Concurrent Male-Male Partnerships in the Context of Low Human Immunodeficiency Virus Testing Among Men Who Have Sex With Men in Bamako, Mali. Sex Transm Dis 2017; 44(9): 565–70.

140. Tiamiyu AB, Lawlor J, Hu F, et al. HIV status disclosure by Nigerian men who have sex with men and transgender women living with HIV: a cross-sectional analysis at enrollment into an observational cohort. BMC Public Health 2020; 20(1): 1282.

141. Lyons CE, Ketende S, Diouf D, et al. Potential Impact of Integrated Stigma Mitigation Interventions in Improving HIV/AIDS Service Delivery and Uptake for Key Populations in Senegal. J Acquir Immune Defic Syndr 2017; 74 Suppl 1(Suppl 1): S52–s9.

142. Mason K, Ketende S, Peitzmeier S, et al. A cross-sectional analysis of population demographics, HIV knowledge and risk behaviors, and prevalence and associations of HIV among men who have sex with men in the Gambia. AIDS Res Hum Retroviruses 2013; 29(12): 1547–52.

143. Stahlman S, Johnston LG, Yah C, et al. Respondent-driven sampling as a recruitment method for men who have sex with men in southern sub-Saharan Africa: a cross-sectional analysis by wave. Sex Transm Infect 2016; 92(4): 292–8.

144. Baral S, Burrell E, Scheibe A, Brown B, Beyrer C, Bekker LG. HIV risk and associations of HIV infection among men who have sex with men in peri-urban Cape Town, South Africa. BMC Public Health 2011; 11: 766.

145. Boothe MAS, Sathane I, Baltazar CS, et al. Low engagement in HIV services and progress through the treatment cascade among key populations living with HIV in Mozambique: alarming gaps in knowledge of status. BMC Public Health 2021; 21(1): 146.

146. Ross MW, Nyoni J, Ahaneku HO, Mbwambo J, McClelland RS, McCurdy SA. High HIV seroprevalence, rectal STIs and risky sexual behaviour in men who have sex with men in Dar es Salaam and Tanga, Tanzania. BMJ Open 2014; 4(8): e006175.

147. Baral S, Trapence G, Motimedi F, et al. HIV prevalence, risks for HIV infection, and human rights among men who have sex with men (MSM) in Malawi, Namibia, and Botswana. PLoS One 2009; 4(3): e4997.

148. Korhonen C, Kimani M, Wahome E, et al. Depressive symptoms and problematic alcohol and other substance use in 1476 gay, bisexual, and other MSM at three research sites in Kenya. Aids 2018; 32(11): 1507–15.

149. van Liere G, Kock MM, Radebe O, et al. High Rate of Repeat Sexually Transmitted Diseases Among Men Who Have Sex With Men in South Africa: A Prospective Cohort Study. Sex Transm Dis 2019; 46(11): e105–e7.

150. Okoboi S, Castelnuovo B, Van Geertruyden JP, et al. Cost-Effectiveness of Peer-Delivered HIV Self-Tests for MSM in Uganda. Front Public Health 2021; 9: 651325.

151. Harris TG, Wu Y, Parmley LE, et al. HIV care cascade and associated factors among men who have sex with men, transgender women, and genderqueer individuals in Zimbabwe: findings from a biobehavioural survey using respondent-driven sampling. The Lancet HIV 2022; 9(3): e182–e201.

152. Baral SD, Ketende S, Schwartz S, et al. Evaluating respondent-driven sampling as an implementation tool for universal coverage of antiretroviral studies among men who have sex with men living with HIV. J Acquir Immune Defic Syndr 2015; 68 Suppl 2(0 2): S107–13.

153. Li Y, Liu H, Ramadhani HO, et al. Genetic clustering analysis for HIV infection among MSM in Nigeria: implications for intervention. Aids 2020; 34(2): 227–36.

154. Graham SM, Mugo P, Gichuru E, et al. Adherence to antiretroviral therapy and clinical outcomes among young adults reporting high-risk sexual behavior, including men who have sex with men, in coastal Kenya. AIDS Behav 2013; 17(4): 1255–65.

155. Kunzweiler CP, Bailey RC, Okall DO, Graham SM, Mehta SD, Otieno FO. Factors Associated With Prevalent HIV Infection Among Kenyan MSM: The Anza Mapema Study. J Acquir Immune Defic Syndr 2017; 76(3): 241–9.

156. Ogunbajo A, Kershaw T, Kushwaha S, Boakye F, Wallace-Atiapah ND, Nelson LE. Barriers, Motivators, and Facilitators to Engagement in HIV Care Among HIV-Infected Ghanaian Men Who have Sex with Men (MSM). AIDS Behav 2018; 22(3): 829–39.

157. Ibiloye O, Akande P, Plang J, et al. Community health worker-led ART delivery improved scheduled antiretroviral drug refill among men who have sex with men in Lagos State, Nigeria. Int Health 2021; 13(2): 196–8.

158. Ibiloye O, Jwanle P, Masquillier C, et al. Long-term retention and predictors of attrition for key populations receiving antiretroviral treatment through community-based ART in Benue State Nigeria: A retrospective cohort study. PLoS One 2021; 16(11): e0260557.

159. Gu LY, Zhang N, Mayer KH, et al. Autonomy-Supportive Healthcare Climate and HIV-Related Stigma Predict Linkage to HIV Care in Men Who Have Sex With Men in Ghana, West Africa. J Int Assoc Provid AIDS Care 2021; 20: 2325958220978113.

160. Rucinski K, Masankha Banda L, Olawore O, et al. HIV Testing Approaches to Optimize Prevention and Treatment for Key and Priority Populations in Malawi. Open Forum Infect Dis 2022; 9(4): ofac038.

161. Offie DC, Obeagu EI, Akueshi C, et al. Facilitators and Barriers to Retention in HIV Care among HIV Infected MSM Attending Community Health Center Yaba, Lagos Nigeria. Journal of Pharmaceutical Research International 2021.

162. Afolaranmi TO, Hassan ZI, Ugwu OJ, et al. Retention in HIV care and its predictors among HIV-infected men who have sex with men in Plateau state, North Central Nigeria. J Family Med Prim Care 2021; 10(4): 1596–601.

163. Dah TTE, Yaya I, Mensah E, et al. Rapid antiretroviral therapy initiation and its effect on treatment response in MSM in West Africa. Aids 2021; 35(13): 2201–10.

164. Ndiaye HD, Tchiakpe E, Vidal N, et al. HIV type 1 subtype C remains the predominant subtype in men having sex with men in Senegal. AIDS Res Hum Retroviruses 2013; 29(9): 1265–72.

165. Mboumba Bouassa RS, Mbeko Simaleko M, Camengo SP, et al. Unusual and unique distribution of anal high-risk human papillomavirus (HR-HPV) among men who have sex with men living in the Central African Republic. PLoS One 2018; 13(5): e0197845.

166. Kunzweiler CP, Bailey RC, Mehta SD, et al. Factors associated with viral suppression among HIV-positive Kenyan gay and bisexual men who have sex with men. AIDS Care 2018; 30(Sup5): S76–s88.

167. Rees K, Radebe O, Arendse C, et al. Utilization of Sexually Transmitted Infection Services at 2 Health Facilities Targeting Men Who Have Sex With Men in South Africa: A Retrospective Analysis of Operational Data. Sex Transm Dis 2017; 44(12): 768–73.

168. Koyalta D, Mboumba Bouassa RS, Maiga AI, et al. High Prevalence of Anal Oncogenic Human Papillomavirus Infection in Young Men Who Have Sex with Men Living in Bamako, Mali. Infect Agent Cancer 2021; 16(1): 51.

169. Gebrebrhan H, Kambaran C, Sivro A, et al. Rectal microbiota diversity in Kenyan MSM is inversely associated with frequency of receptive anal sex, independent of HIV status. Aids 2021; 35(7): 1091–101.

170. Yaya I, Boyer V, Ehlan PA, et al. Heterogeneity in the Prevalence of High-Risk Human Papillomavirus Infection in Human Immunodeficiency Virus-Negative and Human Immunodeficiency Virus-Positive Men Who Have Sex With Men in West Africa. Clin Infect Dis 2021; 73(12): 2184–92.

171. Hakim AJ, Coy K, Patnaik P, et al. An urgent need for HIV testing among men who have sex with men and transgender women in Bamako, Mali: Low awareness of HIV infection and viral suppression among those living with HIV. PLoS One 2018; 13(11): e0207363.

172. Cloete A, Simbayi LC, Kalichman SC, Strebel A, Henda N. Stigma and discrimination experiences of HIV-positive men who have sex with men in Cape Town, South Africa. AIDS Care 2008; 20(9): 1105–10.

173. Billings E, Kijak GH, Sanders-Buell E, et al. New Subtype B Containing HIV-1 Circulating Recombinant of sub-Saharan Africa Origin in Nigerian Men Who Have Sex With Men. J Acquir Immune Defic Syndr 2019; 81(5): 578–84.

174. Palumbo PJ, Zhang Y, Clarke W, et al. Uptake of antiretroviral treatment and viral suppression among men who have sex with men and transgender women in sub-Saharan Africa in an observational cohort study: HPTN 075. Int J Infect Dis 2021; 104: 465–70.

175. Ibiloye O, Decroo T, Eyona N, Eze P, Agada P. Characteristics and early clinical outcomes of key populations attending comprehensive community-based HIV care: Experiences from Nasarawa State, Nigeria. PLoS One 2018; 13(12): e0209477.

176. Kufa T, Lane T, Manyuchi A, et al. The accuracy of HIV rapid testing in integrated bio-behavioral surveys of men who have sex with men across 5 Provinces in South Africa. Medicine (Baltimore) 2017; 96(28): e7391.

177. Parmley LE, Chingombe I, Wu Y, et al. High Burden of Active Syphilis and Human Immunodeficiency Virus/Syphilis Coinfection Among Men Who Have Sex With Men, Transwomen, and Genderqueer Individuals in Zimbabwe. Sex Transm Dis 2022; 49(2): 111–6.

178. Graham SM, Micheni M, Chirro O, et al. A Randomized Controlled Trial of the Shikamana Intervention to Promote Antiretroviral Therapy Adherence Among Gay, Bisexual, and Other Men Who Have Sex with Men in Kenya: Feasibility, Acceptability, Safety and Initial Effect Size. AIDS Behav 2020; 24(7): 2206–19.

179. !!! INVALID CITATION !!! (29, 35, 37-42, 44, 45, 47, 50, 55, 56, 58, 68, 70, 72, 80, 86, 93, 100, 103, 106, 113, 121, 122, 125, 126, 130, 135, 137, 140, 143, 144, 146, 147, 152, 153, 155, 158, 161, 165, 167-171, 173-175, 177, 179, 183, 185, 187, 188, 192, 194-196).

180. Horth RZ, Cummings B, Young PW, et al. Correlates of HIV Testing Among Men Who have Sex with Men in Three Urban Areas of Mozambique: Missed Opportunities for Prevention. AIDS Behav 2015; 19(11): 1978–89.

181. UNAIDS. UNAIDS Data 2021. 2021.

182. Garnett GP. Reductions in HIV incidence are likely to increase the importance of key population programmes for HIV control in sub-Saharan Africa. Journal of the International AIDS Society 2021; 24(S3): e25727.

183. UNAIDS. Global AIDS Update. Seizing the moment: Tackling entrenched inequalities to end epidemics. 2020.

184. Irungu EM, Baeten JM. PrEP rollout in Africa: status and opportunity. Nature Medicine 2020; 26(5): 655–64.

185. Sun Z, Gu Q, Dai Y, et al. Increasing awareness of HIV pre-exposure prophylaxis (PrEP) and willingness to use HIV PrEP among men who have sex with men: a systematic review and meta-analysis of global data. Journal of the International AIDS Society 2022; 25(3): e25883.

186. Karuga RN, Njenga SN, Mulwa R, et al. “How I Wish This Thing Was Initiated 100 Years Ago!” Willingness to Take Daily Oral Pre-Exposure Prophylaxis among Men Who Have Sex with Men in Kenya. PLOS ONE 2016; 11(4): e0151716.

187. Ogunbajo A, Tsai AC, Kanki PJ, Mayer KH. Acceptability of and Preferences for Long-Acting Injectable HIV PrEP and Other PrEP Modalities among Sexual Minority Men in Nigeria, Africa. AIDS and Behavior 2022; 26(7): 2363–75.

188. McGillen JB, Anderson S-J, Dybul MR, Hallett TB. Optimum resource allocation to reduce HIV incidence across sub-Saharan Africa: a mathematical modelling study. The Lancet HIV 2016; 3(9): e441–e8.

189. Anderson S-J, Cherutich P, Kilonzo N, et al. Maximising the effect of combination HIV prevention through prioritisation of the people and places in greatest need: a modelling study. The Lancet 2014; 384(9939): 249–56.

190. Sullivan PS, Carballo-Diéguez A, Coates T, et al. Successes and challenges of HIV prevention in men who have sex with men. The Lancet 2012; 380(9839): 388–99.

191. Scheibe A, Kanyemba B, Syvertsen J, Adebajo S, Baral S. Money, power and HIV: economic influences and HIV among men who have sex with men in sub-Saharan Africa. African journal of reproductive health 2014; 18(1): 84–92.

192. Inghels M, Kouassi AK, Niangoran S, et al. Preferences and access to community-based HIV testing sites among men who have sex with men (MSM) in Côte d’Ivoire. BMJ open 2022; 12(6): e052536.

193. Giguère K, Eaton JW, Marsh K, et al. Trends in knowledge of HIV status and efficiency of HIV testing services in sub-Saharan Africa, 2000-20: a modelling study using survey and HIV testing programme data. Lancet HIV 2021; 8(5): e284–e93.

194. Xia Y, Milwid RM, Godin A, et al. Accuracy of self-reported HIV-testing history and awareness of HIV-positive status in four sub-Saharan African countries. AIDS 2021; 35(3): 503–10.

195. Soni N, Giguère K, Boily M-C, et al. Under-reporting of known HIV-positive status among people living with HIV: a systematic review and meta-analysis. AIDS and Behavior 2021; 25(12): 3858–70.

196. Organization WH. Consolidated guidelines on HIV, viral hepatitis and STI prevention, diagnosis, treatment and care for key populations. Consolidated guidelines on HIV, viral hepatitis and STI prevention, diagnosis, treatment and care for key populations; 2022.

197. Ehrenkranz P, Grimsrud A, Rabkin M. Differentiated service delivery: navigating the path to scale. Current Opinion in HIV and AIDS 2019; 14(1): 60–5.

198. Johnston LG, Sabin K. Sampling Hard-to-Reach Populations with Respondent Driven Sampling. Methodological Innovations Online 2010; 5(2): 38–48.

199. Adebajo S, Obianwu O, Eluwa G, et al. Comparison of Audio Computer Assisted Self-Interview and Face-To-Face Interview Methods in Eliciting HIV-Related Risks among Men Who Have Sex with Men and Men Who Inject Drugs in Nigeria. PLOS ONE 2014; 9(1): e81981.

200. Risher K, Mayer KH, Beyrer C. HIV treatment cascade in MSM, people who inject drugs, and sex workers. Curr Opin HIV AIDS 2015; 10(6): 420–9.

201. Joshi K, Lessler J, Olawore O, et al. Declining HIV incidence in sub-Saharan Africa: a systematic review and meta-analysis of empiric data. Journal of the International AIDS Society 2021; 24(10): e25818.

