## Supplementary Materials for "Trends in HIV testing, the treatment cascade, and HIV incidence among men who have sex with men in Africa: A systematic review and meta-regression analysis"

**Table S1:** Search terms for HIV testing, treatment cascade, and incidence studies, by database and search domain.

| |
|---|
| **a) Embase search strategy**<br>Search conducted March 24th 2022 – 4622 articles retrieved |
| ***HIV domain*** (exp Human immunodeficiency virus/ OR exp acquired immune deficiency syndrome/ OR exp Human immunodeficiency virus infection/ OR exp Human immunodeficiency virus antibody/ OR exp Human immunodeficiency virus prevalence/ OR exp HIV test/ OR "hiv*".ab,kw,ti. OR human immun#deficiency virus.ab,kw,ti. OR human immun# deficiency virus.ab,kw,ti. OR acquired immun#deficiency syndrome.ab,kw,ti. OR acquired immun# deficiency syndrome.ab,kw,ti. OR "AIDS*".ab,kw,ti. OR SIDA.ab,kw,ti. OR syndrome d'immunodeficience acquise.ab,kw,ti. OR VIH.ab,kw,ti. OR virus de l'immunodeficience humaine.ab,kw,ti.) |
| ***AND MSM domain*** (exp male homosexuality/ OR exp bisexuality/ OR gay.ab,kw,ti. OR MSM.ab,kw,ti. OR men who have sex with men.ab,kw,ti. OR men that have sex with men.ab,kw,ti. OR HRSH.ab,kw,ti. OR hommes qui ont des relations sexuelles avec des hommes.ab,kw,ti. OR same-sex.ab,kw,ti. OR same sex.ab,kw,ti. OR queer.ab,kw,ti. OR "bisex*".ab,kw,ti. OR "homosex*".ab,kw,ti. OR same-gender.ab,kw,ti. OR same gender.ab,kw,ti. OR "meme sex*".ab,kw,ti. OR "meme genre*".ab,kw,ti. OR (male adj2 sex worker*).ab,kw,ti. OR "male sex work*".ab,kw,ti. OR exp men who have sex with men/ OR exp "sexual and gender minority"/ OR exp "men who have sex with men and women"/ OR sexual minority men.ab,kw,ti. OR (sexual and gender minority men).ab,kw,ti.) |
| ***AND Africa domain*** (exp africa/ OR exp "africa south of the sahara"/ OR exp north africa/ OR exp South Africa/ OR exp North Africa/ OR exp Central Africa/ OR exp African/ OR "Africa*".ab,kw,ti. OR "Afriq*".ab,kw,ti. OR "Algeri*".ab,kw,ti. OR "Angola*".ab,kw,ti. OR "Benin*".ab,kw,ti. OR (Botswana* OR Matswana* OR Batswana*).ab,kw,ti. OR (Burkina* OR Burundi*).ab,kw,ti. OR (Cabo Verde* OR Cape Verde* OR Cap#Vert).ab,kw,ti. OR (Camero* OR Central African Republic* OR republique centrafricaine OR Chad* OR Tchad* OR Comor* OR Cote d'Ivoire OR Ivory Coast OR Ivorian*).ab,kw,ti. OR (Djibouti OR Democratic Republic of the Congo OR Democratic Republic of the Congo OR Congo*).ab,kw,ti. OR (Egypt* OR Equatorial Guinea* OR Guinee Equatoriale OR Equatoguinean* OR Eritrea* OR Erythree* OR eSwatini* OR Ethiop*).ab,kw,ti. OR (Gabon* OR Gambi* OR Ghana* OR Guine*).ab,kw,ti. OR "Kenya*".ab,kw,ti. OR (Lesotho* OR Bathoso* OR Liberia* OR Liby*).ab,kw,ti. OR (Madagas* OR Malawi* OR Mali* OR Maurit* OR Moroc* OR Maroc* OR Mozambi*).ab,kw,ti. OR (Namibi* OR Niger*).ab,kw,ti. OR (Rwanda* OR Rouanda* OR Ruanda*).ab,kw,ti. OR (Sao* OR Senegal* OR Seychel* OR Sierra Leon* OR Somali* OR South Africa* OR Afrique du Sud OR South Sudan* OR Soudan du sud OR Sudan* OR Swazi*).ab,kw,ti. OR (Tanzani* OR Togo* OR Republique togolaise or tunisi*).ab,kw,ti. OR (Uganda* OR Ouganda*).ab,kw,ti. OR (Zambi* OR Zimbabwe*).ab,kw,ti. OR exp Algeria/ OR exp Angola/ OR exp Benin/ OR exp Botswana/ OR exp Burkina Faso/ OR exp Burundi/ OR exp Cape Verde/ OR exp Cameroon/ OR exp Central African Republic/ OR exp Chad/ OR exp Comoros/ OR exp Cote d'Ivoire/ OR exp Djibouti/ OR exp Congo/ OR exp Democratic Republic Congo/ OR exp Egypt/ OR exp Guinea-Bissau/ OR exp Guinea/ OR exp Equatorial Guinea/ OR exp Eritrea/ OR exp Ethiopia/ OR exp Gabon/ OR exp Gambia/ OR exp Ghana/ OR exp Kenya/ OR exp Lesotho/ OR exp Liberia/ OR exp Libyan Arab Jamahiriya/ OR exp Madagascar/ OR exp Malawi/ OR exp Mali/ OR exp Mauritania/ OR exp Mauritius/ OR exp Morocco/ OR exp Mozambique/ OR exp Namibia/ OR exp Niger/ OR exp Nigeria/ OR exp Rwanda/ OR exp "Sao Tome and Principe"/ OR exp Senegal/ OR exp Seychelles/ OR exp Sierra Leone/ OR exp Somalia/ OR exp South Africa/ OR exp South Sudan/ OR exp Sudan/ OR exp Eswatini/ OR exp Tanzania/ OR exp Togo/ OR exp Tunisia/ OR exp Uganda/ OR exp Zambia/ OR exp Zambia/ OR exp Zimbabwe/) |
| ***AND limit to yr=”1980-Current”*** |
| **b) Medline search strategy**<br>Search conducted March 24th 2022 – 3163 articles retrieved |
| ***HIV domain*** (exp HIV/ OR exp hiv infections/ OR exp acquired immunodeficiency syndrome/ OR exp HIV testing/ OR exp HIV seropositivity/ OR (HIV* OR human immun#deficiency virus OR human immun# deficiency virus OR acquired immun#deficiency syndrome OR acquired immun# deficiency syndrome OR AIDS* OR SIDA OR syndrome d'immunodeficience acquise OR VIH OR virus de l'immunodeficience humaine).ab,kw,ti.) |
| ***AND MSM domain*** (exp Homosexuality, Male/ OR exp Bisexuality/ OR exp "Sexual and Gender Minorities"/ OR "homosex*".ab,kw,ti. OR sexual minority men.ab,kw,ti. OR (sexual and gender minority men).ab,kw,ti. OR (gay OR MSM OR men who have sex with men OR men that have sex with men).ab,kw,ti. OR (HRSH OR hommes qui ont des relations sexuelles avec des hommes).ab,kw,ti. OR (same-sex OR same sex OR same-gender OR same gender OR queer OR bisex*).ab,kw,ti. OR (male adj2 sex worker*).ab,kw,ti. OR "male sex work*".ab,kw,ti. OR (meme sex* OR meme genre*).ab,kw,ti.) |

| |
|---|
| ***AND Africa domain*** (exp Africa, Central/ OR exp "Africa South of the Sahara"/ OR exp Africa, Southern/ OR exp Africa, Northern/ OR exp Africa, Western/ OR exp Africa, Eastern/ OR exp Africa/ OR exp South Africa/ OR (Africa* OR Afriq*).ab,kw,ti. OR exp Algeria/ OR exp Angola/ OR exp Benin/ OR exp Botswana/ OR exp Burkina Faso/ OR exp Burundi/ OR exp Cabo Verde/ OR exp Cameroon/ OR exp Central African Republic/ OR exp Chad/ OR exp Comoros/ OR exp Cote d'Ivoire/ OR exp Djibouti/ OR exp "Democratic Republic of the Congo"/ OR exp Congo/ OR exp Egypt/ OR exp Guinea/ OR exp Equatorial Guinea/ OR exp Guinea-Bissau/ OR exp Eritrea/ OR exp Ethiopia/ OR exp Gabon/ OR exp Gambia/ OR exp Ghana/ OR exp Kenya/ OR exp Lesotho/ OR exp Liberia/ OR exp Libya/ OR exp Madagascar/ OR exp Malawi/ OR exp Mali/ OR exp Mauritania/ OR exp Mauritius/ OR exp Morocco/ OR exp Mozambique/ OR exp Namibia/ OR exp Niger/ OR exp Nigeria/ OR exp Rwanda/ OR exp "Sao Tome and Principe"/ OR exp Senegal/ OR exp Seychelles/ OR exp Sierra Leone/ OR exp Somalia/ OR exp Sudan/ OR exp South Sudan/ OR exp Eswatini/ OR exp Tanzania/ OR exp Togo/ OR exp Tunisia/ OR exp Uganda/ OR exp Zambia/ OR exp Zimbabwe/ OR (Algeri* OR Angola*).ab,kw,ti. OR (Benin* OR Botswana* OR Motswana* OR Batswana* OR Burkina* OR Burundi*).ab,kw,ti. OR (Cabo Verde* OR Cape Verde* OR Cap-Vert).ab,kw,ti. OR (Camero* OR Central African Republic* OR republique centrafricaine OR Chad* OR Tchad* OR Comor* OR Cote d'Ivoire OR Ivory Coast OR Ivorian*).ab,kw,ti. OR (Djibouti OR Democratic Republic of the Congo OR Democratic Republic of the Congo OR Congo*).ab,kw,ti. OR (Egypt* OR Equatorial Guinea* OR Guinee Equatoriale OR Equatoguinean* OR Eritrea* OR Erythree* OR eSwatini* OR Ethiop*).ab,kw,ti. OR (Gabon* OR Gambi* OR Ghana* OR Guine*).ab,kw,ti. OR "Kenya*".ab,kw,ti. OR (Lesotho* OR Bathoso* OR Liberia* OR Liby*).ab,kw,ti. OR (Madagas* OR Malawi* OR Mali* OR Maurit* OR Moroc* OR Maroc* OR Mozambi*).ab,kw,ti. OR (Namibi* OR Niger*).ab,kw,ti. OR (Rwanda* OR Rouanda* OR Ruanda*).ab,kw,ti. OR (Sao* OR Senegal* OR Seychel* OR Sierra Leon* OR Somali* OR South Africa* OR Afrique du Sud OR South Sudan* OR Soudan du sud OR Sudan* OR Swazi*).ab,kw,ti. OR (Tanzani* OR Togo* OR Republique togolaise OR tunisi*).ab,kw,ti. OR (Uganda* OR Ouganda*).ab,kw,ti. OR (Zambi* OR Zimbabwe*).ab,kw,ti.) |
| ***AND limit to yr="1980-Current"*** |
| **c) Global Health search strategy**<br>Search conducted March 24th 2022 – 1951 articles retrieved |
| ***HIV domain*** (exp human immunodeficiency viruses/ OR exp human immunodeficiency virus 1/ OR exp human immunodeficiency virus 2/ OR exp acquired immune deficiency syndrome/ OR exp aids related complex/ OR exp hiv infections/ OR exp hiv-1 infections/ OR exp hiv-2 infections/ OR (HIV* OR human immun#deficiency virus OR human immun# deficiency virus OR acquired immun#deficiency syndrome OR acquired immun# deficiency syndrome OR AIDS* OR SIDA OR syndrome d'immunodeficience acquise OR VIH OR virus de l'immunodeficience humaine).ab,ti.) |
| ***AND MSM domain*** (exp homosexuality/ OR exp homosexual transmission/ OR exp men who have sex with men/ OR exp bisexuality/ OR exp homosexual men/ OR (gay OR MSM OR men who have sex with men OR men that have sex with men).ab,ti. OR (HRSH OR hommes qui ont des relations sexuelles avec des hommes).ab,ti. OR (same-sex OR same sex OR same-gender OR same gender OR queer OR bisex*).ab,ti. OR "male sex work*".ab,ti. OR (male adj2 sex work*).ab,ti. OR (meme sex* OR meme genre*).ab,ti. OR "homosex*".ab,ti. OR sexual minority men.ab,ti. OR (sexual and gender minority men).ab,ti.) |
| ***AND Africa domain*** (exp "Africa South of Sahara"/ OR exp East Africa/ OR exp Africa/ OR exp Central Africa/ OR exp North Africa/ OR exp Southern Africa/ OR exp West Africa/ OR exp Algeria/ OR exp Angola/ OR exp Benin/ OR exp Botswana/ OR exp Burkina Faso/ OR exp Burundi/ OR exp Cape Verde/ OR exp Cameroon/ OR exp Central African Republic/ OR exp Chad/ OR exp Comoros/ OR exp Cote d'Ivoire/ OR exp Djibouti/ OR exp Congo/ OR exp Congo Democratic Republic/ OR exp Egypt/ OR exp Equatorial Guinea/ OR exp Guinea-Bissau/ OR exp Guinea/ OR exp Eritrea/ OR exp Ethiopia/ OR exp Gabon/ OR exp Gambia/ OR exp Ghana/ OR exp Kenya/ OR exp Lesotho/ OR exp Liberia/ OR exp Libya/ OR exp Madagascar/ OR exp Malawi/ OR exp Mali/ OR exp Mauritania/ OR exp Mauritius/ OR exp Morocco/ OR exp Mozambique/ OR exp Namibia/ OR exp Niger/ OR exp Nigeria/ OR exp Rwanda/ OR exp "sao tome and principe"/ OR exp Senegal/ OR exp Seychelles/ OR exp Sierra Leone/ OR exp Somalia/ OR exp South Africa/ OR exp South Sudan/ OR exp Sudan/ OR exp swaziland/ OR exp Tanzania/ OR exp Togo/ OR exp Tunisia/ OR exp Uganda/ OR exp Zambia/ OR exp Zimbabwe/ OR (Africa* OR Afriq*).ab,ti. OR (Algeri* OR Angola*).ab,ti. OR (Benin* OR Botswana* OR Motswana* OR Batswana* OR Burkina* OR Burundi*).ab,ti. OR (Cabo Verde* OR Cape Verde* OR Cap-Vert).ab,ti. OR (Camero* OR Central African Republic* OR republique centrafricaine OR Chad* OR Tchad* OR Comor* OR Cote d'Ivoire OR Ivory Coast or Ivorian*).ab,ti. OR (Djibouti OR Democratic Republic of the Congo OR Democratic Republic of the Congo OR Congo*).ab,ti. OR (Egypt* OR Equatorial Guinea* OR Guinee Equatoriale OR Equatoguinean* OR Eritrea* OR Erythree* OR eSwatini* OR Ethiop*).ab,ti. OR (Gabon* OR Gambi* OR Ghana* OR Guine*).ab,ti. OR "Kenya*".ab,ti. OR (Lesotho* OR Bathoso* OR Liberia* OR Liby*).ab,ti. OR (Madagas* OR Malawi* OR Mali* OR Maurit* OR Moroc* OR Maroc* OR Mozambi*).ab,ti. OR (Namibi* OR Niger*).ab,ti. OR (Rwanda* OR Rouanda* OR Ruanda*).ab,ti. OR (Sao* OR Senegal* OR Seychel* OR Sierra Leon* OR Somali* OR South Africa* OR Afrique du Sud OR South Sudan* OR Soudan du sud OR Sudan* OR Swazi*).ab,ti. OR (Tanzani* OR Togo* OR Republique togolaise or tunisi*).ab,ti. OR (Uganda* OR Ouganda*).ab,ti. OR (Zambi* OR Zimbabwe*).ab,ti.) |
| ***AND limit to yr="1980-Current"*** |
| **d) Scopus search strategy**<br>Search conducted March 24th 2022 – 5451 articles retrieved |
| ***HIV domain*** (TITLE-ABS-KEY(aids*) OR TITLE-ABS-KEY("acquired immune deficiency syndrome") OR TITLE-ABS-KEY("acquired immun?deficiency syndrome") OR TITLE-ABS-KEY("acquired immun? deficiency syndrome") OR TITLE- |

| |
|---|
| ABS-KEY(HIV*) OR TITLE-ABS-KEY("human immun?deficiency virus") OR TITLE-ABS-KEY("human immun? deficiency virus") OR TITLE-ABS-KEY(SIDA) OR TITLE-ABS-KEY("syndrome d'immunodeficience acquise") OR TITLE-ABS-KEY(VIH) OR TITLE-ABS-KEY("virus de l'immunodeficience humaines")) |
| ***AND MSM domain*** (TITLE-ABS-KEY(homosex*) OR TITLE-ABS-KEY(bisex*) OR TITLE-ABS-KEY("men who have sex with men") OR TITLE-ABS-KEY("men that have sex with men") OR TITLE-ABS-KEY("same sex") OR TITLE-ABS-KEY("same-sex") OR TITLE-ABS-KEY(gay) OR TITLE-ABS-KEY(MSM) OR TITLE-ABS-KEY(queer) OR TITLE-ABS-KEY("male sex work*") OR TITLE-ABS-KEY("male W/2 sex work*") OR TITLE-ABS-KEY("same gender") OR TITLE-ABS-KEY("same-gender") OR TITLE-ABS-KEY("meme sex*") OR TITLE-ABS-KEY("meme genre*") OR TITLE-ABS-KEY(HRSH) OR TITLE-ABS-KEY("hommes qui ont des relations sexuelles avec des hommes") OR TITLE-ABS-KEY("sexual minority men") OR TITLE-ABS-KEY("sexual and gender minority men")) |
| ***AND Africa domain*** (TITLE-ABS-KEY(africa*) OR TITLE-ABS-KEY(afriq*) OR TITLE-ABS-KEY(algeri*) OR TITLE-ABS-KEY(angola*) OR TITLE-ABS-KEY(benin*) OR TITLE-ABS-KEY(botswana*) OR TITLE-ABS-KEY(motswana*) OR TITLE-ABS-KEY(batswana*) OR TITLE-ABS-KEY(burkina*) OR TITLE-ABS-KEY(burundi*) OR TITLE-ABS-KEY("Cabo Verde*") OR TITLE-ABS-KEY("Cape Verde*") OR TITLE-ABS-KEY("Cap-Vert") OR TITLE-ABS-KEY(Camero*) OR TITLE-ABS-KEY("Central African Republic*") OR TITLE-ABS-KEY("republique centrafricaine") OR TITLE-ABS-KEY(chad*) OR TITLE-ABS-KEY(tchad*) OR TITLE-ABS-KEY(comor*) OR TITLE-ABS-KEY("cote d'ivoire") OR TITLE-ABS-KEY("ivory coast") OR TITLE-ABS-KEY(ivorian*) OR TITLE-ABS-KEY(djibouti*) OR TITLE-ABS-KEY("democratic republic of the congo") OR TITLE-ABS-KEY(congo*) OR TITLE-ABS-KEY(egypt*) OR TITLE-ABS-KEY("equatorial guinea*") OR TITLE-ABS-KEY("guinee equatoriale") OR TITLE-ABS-KEY(equatoguinean*) OR TITLE-ABS-KEY(eritrea*) OR TITLE-ABS-KEY(erythree*) OR TITLE-ABS-KEY(ethiop*) OR TITLE-ABS-KEY(gabon*) OR TITLE-ABS-KEY(gambi*) OR TITLE-ABS-KEY(ghana*) OR TITLE-ABS-KEY(guine*) OR TITLE-ABS-KEY(kenya*) OR TITLE-ABS-KEY(lesotho*) OR TITLE-ABS-KEY(bathoso*) OR TITLE-ABS-KEY(liberia*) OR TITLE-ABS-KEY(liby*) OR TITLE-ABS-KEY(madagas*) OR TITLE-ABS-KEY(malawi*) OR TITLE-ABS-KEY(mali*) OR TITLE-ABS-KEY(maurit*) OR TITLE-ABS-KEY(moroc*) OR TITLE-ABS-KEY(maroc*) OR TITLE-ABS-KEY(mozambi*) OR TITLE-ABS-KEY(namibi*) OR TITLE-ABS-KEY(niger*) OR TITLE-ABS-KEY(rwanda*) OR TITLE-ABS-KEY(rouanda*) OR TITLE-ABS-KEY(ruanda*) OR TITLE-ABS-KEY(sao*) OR TITLE-ABS-KEY(senegal*) OR TITLE-ABS-KEY(seychel*) OR TITLE-ABS-KEY("sierra leone*") OR TITLE-ABS-KEY(somali*) OR TITLE-ABS-KEY("south africa*") OR TITLE-ABS-KEY("afrique du sud") OR TITLE-ABS-KEY("south sudan*") OR TITLE-ABS-KEY("soudan du sud") OR TITLE-ABS-KEY(sudan*) OR TITLE-ABS-KEY(soudan*) OR TITLE-ABS-KEY(swazi*) OR TITLE-ABS-KEY(eswatini) OR TITLE-ABS-KEY(tanzani*) OR TITLE-ABS-KEY(togo*) OR TITLE-ABS-KEY(tunisi*) OR TITLE-ABS-KEY("republique togolaise") OR TITLE-ABS-KEY(uganda*) OR TITLE-ABS-KEY(ouganda*) OR TITLE-ABS-KEY(zambi*) OR TITLE-ABS-KEY(zimbabwe*)) |
| ***AND PUBYEAR > 1979*** |
| **e) Web of Science search strategy**<br>Search conducted March 24th 2022 – 4232 articles retrieved |
| ***HIV domain*** (TS = (AIDS* or "acquired immune deficiency syndrome" or "acquired immun?deficiency syndrome" or "acquired immun? deficiency syndrome" or HIV* or "human immun?deficiency virus" or "human immun? deficiency virus" or SIDA or "syndrome d'immunodeficience acquise" or VIH or "virus de l'immunodeficience humaine")) |
| ***AND MSM domain*** (TS = (homosex* or bisex* or "men who have sex with men" or "men that have sex with men" or "same sex" or "same-sex" or "same gender" or "same-gender" or gay or MSM or queer or "male sex work*" or male near/2 "sex work*" or "meme sex*" or "meme genre*" or "harsh" or "hommes qui ont des relations sexuelles avec des hommes" or "sexual minority men" or "sexual and gender minority men")) |
| ***AND Africa domain*** (TS = (Africa* or afriq* or algeri* or angola* or benin* or botswana* or motswana* or batswana* or burkina* or burundi* or "cabo verde*" or "cap-vert*" or "cape verde*" or camero* or "central african republic*" or "republique centrafricaine" or chad* or tchad* or comor* or "cote d'ivoire" or "ivory coast" or ivorian* or djibouti* or "democratic republic of the congo" or congo* or egypt* or "equatorial guinea*" or "guinee equatoriale" or equatoguinean* or eritrea* or erythree* or ethiop* or gabon* or gambi* or ghana* or guine* or kenya* or lesotho* or bathoso* or liberia* or liby* or madagas* or malai* or mali* or maurit* or moroc* or maroc* or mozambi* or namibi* or niger* or rwanda* or rouanda* or ruanda* or sao* or senegal* or seychel* or "sierra leone*" or somali* or "south africa*" or "afrique du sud" or "south sudan*" or "soudan du sud" or sudan* or soudan* or swazi* or eswatini* or tanzani* or togo* or tunisi* or uganda* or ouganda* or zambi* or zimbabwe*)) |
| ***AND Timespan=1980-2022*** |

**Text S1. Including respondent driven sampling-adjusted HIV testing and treatment cascade proportions which accounted for sampling design**

To derive the estimated proportion of HIV testing and treatment cascade outcomes in R requires specifying the numerator (n) and denominator (N) of observations before pooling. As pooling does not account for design effect, we conducted extra steps to be able to include observations from respondent-driven sampling (RDS) and time-location or cluster sampling studies that reported weighted proportions adjusted for sampling design, which typically have a wider confidence interval than the corresponding crude proportion (n/N), due to the design effect). In practice, this only applied to RDS studies.

To include RDS-adjusted observations that accounted for sampling design in our meta-regression analyses, we extracted the RDS-adjusted proportion ($p_{rds}$) and the RDS-adjusted 95% confidence interval ($95\%CI_{rds}$) from studies that reported them. We then used these to obtain an estimate of the design effect ($DE_{rds}$), which we calculated from the ratio of variances of the RDS-adjusted proportion and the simple random sample (SRS) proportion for each adjusted observation reported. We then used the design effect to derive the effective sample size, including estimates of the numerator ($n_{rds}$) and denominator ($N_{rds}$), which were included in our meta-regression analyses.

To estimate the effective numerator and denominator of adjusted observations and their 95%CI reported in RDS studies, we used the information on n, N, $p_{rds}$, and $95\%CI_{rds}$ and performed the following steps:

1) Derive the variance of the RDS-adjusted proportion from the $95\%CI_{rds}$:

$$\text{var}_{\text{rds}} = \left(\frac{p_{\text{rds\_uci}} - p_{\text{rds\_lci}}}{3.92}\right)^2$$

where $\text{var}_{rds}$ is the variance of the RDS-adjusted proportion accounting for sampling design and $p_{rds\_uci}$ and $p_{rds\_lci}$ are the upper and lower confidence limits of the $95\%CI_{rds}$.

2) Derive the variance of the SRS proportion, using the RDS-adjusted proportion and the crude sample size, N:

$$\text{var}_{\text{srs}} = \frac{p_{\text{rds}} \times (1 - p_{\text{rds}})}{N}$$

where $\text{var}_{srs}$ is the variance of the RDS proportion not accounting for sampling design (as in a simple random sample).

3) Derive the design effect (the ratio of the variances of the RDS-adjusted and SRS proportions):

$$\text{DE}_{\text{rds}} = \frac{\text{var}_{\text{rds}}}{\text{var}_{\text{srs}}}$$

where $DE_{rds}$ is the design effect.

4) Derive the effective sample size from the crude sample size and the design effect:

$$N_{\text{rds}} = \frac{N}{\text{DE}_{\text{rds}}}$$

where $N_{rds}$ is the effective sample size/denominator.

5) Finally, derive the effective numerator for the RDS-adjusted observations:

$$n_{\mathrm{rds}} = N_{\mathrm{rds}} \times p_{\mathrm{rds}}$$

6) Use $n_{rds}$ and $N_{rds}$ in the meta-regression analyses

**Text S2. Details of model specifications of generalized linear mixed effects models for meta-regression by study year**

Depending on the outcomes, either Bayesian logistic or Poisson generalized mixed effects model (GLMM) are used. These are detailed below.

*Binomial regression model for proportions*

For HIV testing, knowledge of status, current ART use, and viral suppression outcomes among men who have sex with men (MSM) in Africa, we used a binomial regression model. It takes the following form:

$$y_i \sim \text{Binomial}(n_i, \theta_i)$$

$$\text{logit}(\theta_i) = a_i + b_i$$

Where $y_i$ is the number of MSM with the outcome (e.g., ever or recently testing for HIV, who know their status, currently on ART, or virally suppressed) in study *i*. These are assumed to follow a binomial distribution. The logit-transformed proportion $\theta_i$ is modeled as the sum of study-specific intercepts $a_i$, and the time trend $b_i$.

$$a_i = \alpha_g + \alpha_{r[i]} + \alpha_{c[i]} + \alpha_{s[i]}$$

The random intercept $a_i$ for study *i*, corresponds to the sum of the global intercept $\alpha_g$, the region-level intercept $\alpha_{r[i]}$ for region *r*, the country-level intercept $\alpha_{c[i]}$ for country *c*, and the survey-specific intercept $\alpha_{s[i]}$ for survey *s*.

$$b_i = (\beta_g + \beta_{r[i]} + \beta_{c[i]})X_i$$

The time trend $b_i$ for study *i* is modeled as a random slope. It corresponds to the sum of the global time trend $\beta_g$, the region-level time trend $\beta_{r[i]}$, and the country-level time trend $\beta_{c[i]}$. These coefficients are then multiplied by the mean-centered calendar year of the study's midpoint $X_i$.

The model's specification is complemented with the following prior distributions. We assumed that the global intercept parameter, $\alpha_g$, and global slope parameter, $\beta_g$, follow normal distributions. We used weakly informative prior distributions for the country-level and region-level variance parameters of the random intercepts and random slopes, assuming half-normal distributions, and selected the hyperparameters such that the variance was higher across regions than countries, as we expect outcomes to be more similar with countries than within regions. We allowed for correlations between random intercepts and slopes using multivariate normal distributions, and used weakly informative priors for the Cholesky factors, $R_c$ and $R_r$ of the correlation matrices that specify the country-level and region-level variance-covaranice matrices $\Sigma_c$, and $\Sigma_r$.

$$\alpha_s \sim \text{Normal}(0, \sigma_s)$$

$$\begin{bmatrix}\alpha_c \\ \beta_c\end{bmatrix} \sim \text{MVNormal}(\begin{bmatrix}0 \\ 0\end{bmatrix}, \Sigma_c)$$

$$\begin{bmatrix}\alpha_r \\ \beta_r\end{bmatrix} \sim \text{MVNormal}(\begin{bmatrix}0 \\ 0\end{bmatrix}, \Sigma_r)$$

$$\alpha_g \sim \text{Normal}(0, 2)$$

$$\beta_g \sim \text{Normal}(0, 1)$$

$$\sigma_s \sim \text{HalfNormal}(0, 1)$$

$$\sigma_{\alpha_c}, \sigma_{\beta_c} \sim \text{HalfNormal}(0, 1)$$

$$\sigma_{\alpha_r}, \sigma_{\beta_r} \sim \text{HalfNormal}(0, 0.5)$$

$$R_c, R_r \sim \text{LKJ}(1)$$

$$\Sigma_c = \begin{bmatrix} \sigma_{\alpha_c} & 0 \\ 0 & \sigma_{\beta_c} \end{bmatrix} * R_c * \begin{bmatrix} \sigma_{\alpha_c} & 0 \\ 0 & \sigma_{\beta_c} \end{bmatrix}$$

$$\Sigma_r = \begin{bmatrix} \sigma_{\alpha_r} & 0 \\ 0 & \sigma_{\beta_r} \end{bmatrix} * R_r * \begin{bmatrix} \sigma_{\alpha_r} & 0 \\ 0 & \sigma_{\beta_r} \end{bmatrix}$$

*Poisson regression model for counts*

For the meta-regression models of HIV incidence rates among MSM in Africa, the model takes the following form:

$$y_i \sim \text{Poisson}(\lambda_i)$$

$$\log(\lambda_i) = a_i + b_i + \log(\delta_i)$$

Where $y_i$ is the number of HIV acquisitions occurring over follow-up in study $i$, that are Poisson distributed. The log-transformed incidence rate $\lambda_i$ is modeled as the sum of study-specific intercepts $a_i$, the time trend $b_i$, and the offset $\log(\delta_i)$ which corresponds to the log-transformed person-years for study $i$. The remainder of the model follows the same specification as above. The model's specification is complemented with the following prior distributions.

$$\sim \text{Normal}(0, \sigma_s)$$

$$\begin{bmatrix} \alpha_c \\ \beta_c \end{bmatrix} \sim \text{MVNormal}(\begin{bmatrix} 0 \\ 0 \end{bmatrix}, \Sigma_c)$$

$$\begin{bmatrix} \alpha_r \\ \beta_r \end{bmatrix} \sim \text{MVNormal}(\begin{bmatrix} 0 \\ 0 \end{bmatrix}, \Sigma_r)$$

$$\alpha_g \sim \text{Normal}(0, 10)$$

$$\beta_g \sim \text{Normal}(0, 1)$$

$$\sigma_s \sim \text{HalfNormal}(0, 1)$$

$$\sigma_{\alpha_c}, \sigma_{\beta_c} \sim \text{HalfNormal}(0, 1)$$

$$\sigma_{\alpha_r}, \sigma_{\beta_r} \sim \text{HalfNormal}(0, 0.5)$$

$$R_c, R_r \sim \text{LKJ}(1)$$

$$\Sigma_c = \begin{bmatrix} \sigma_{\alpha_c} & 0 \\ 0 & \sigma_{\beta_c} \end{bmatrix} * R_c * \begin{bmatrix} \sigma_{\alpha_c} & 0 \\ 0 & \sigma_{\beta_c} \end{bmatrix}$$

$$\Sigma_r = \begin{bmatrix} \sigma_{\alpha_r} & 0 \\ 0 & \sigma_{\beta_r} \end{bmatrix} * R_r * \begin{bmatrix} \sigma_{\alpha_r} & 0 \\ 0 & \sigma_{\beta_r} \end{bmatrix}$$

**Text S3. Study quality assessment tool.**

| Criteria used to assess the quality and risk of bias of included studies |
|---|
| 1) **Appropriateness of the sampling method to recruit a representative sample of MSM participants (maximum 1 points)** |
| a) RDS/cluster/time-location sampling with or without statistical adjustment for study design, or snowball/chain-referral sampling (1 point) |
| b) Convenience or purposive sampling (0 points) |
| c) Sampling strategy not described (0 points) |
| 2) **Statistical adjustment of outcomes for complex survey design (maximum 1 point)** |
| a) Observations of outcome adjusted for complex sampling design (e.g., RDS-adjusted observations; 1 point) |
| b) Crude observations available only (0 points) |
| 3) **Representativeness of MSM participants based on eligibility criteria used to recruit MSM into the study (maximum 1 point)** |
| a) Eligibility criteria designed to recruit a representative sample of MSM participants from the 'general' population of MSM (e.g., not only high-risk MSM, or definition of MSM based on sexual behaviour with another man over recall periods >3 months; 1 point) |
| b) Study recruited a selected sample of MSM participants or eligibility criteria led to more selected sample of MSM (e.g., high-risk MSM, definitions of MSM based on anal sex over recall periods <3 months; 0 points) |
| c) Eligibility criteria not described (0 points) |
| 4) **Inclusion of transgender women in the study definition of MSM (maximum 1 point)** |
| a) Study did not define transgender women as MSM, or outcome(s) were disaggregated and available among only MSM (1 point) |
| b) Transgender women were defined as MSM, or outcomes were not disaggregated (0 points) |
| c) Unclear whether transgender women were included as MSM (0 points) |
| 5) **Risk of misclassification in ascertainment of the relevant outcome(s) reported (maximum 1 point)** |
| a) Confirmed using biomarkers (for incidence and viral suppression outcomes; 1 point) |
| b) Self-report in confidential interview (for all other outcomes; e.g., ACASI, CAPI, SAQ, PBS; 1 point) |
| c) Self-report in face-to-face interview (0 points) |
| d) Ascertainment method not described (0 points) |

ACASI, audio computer-assisted self-interview; CAPI, computer-assisted personal interview; MSM, gay, bisexual, and other men who have sex with men; PBS, pooling booth survey; RCT, randomized controlled trial; RDS, respondent driven sampling; SAQ, self-administered questionnaire.

**Text S4. Additional results pertaining to engagement in care outcomes.**

Observations of engagement in care (other than current ART use) among MSM living with HIV included reports of ever receiving care ($N_o$=3), ever receiving ART ($N_o$=7), currently receiving care ($N_o$=7), being linked to care within 30 days of diagnosis ($N_o$=2), being linked and retained in care within 3 months of diagnosis ($N_o$=1), and being retained in care in the past 12 ($N_o$=1) or 6 months ($N_o$=1). In 6 studies, ever ART use among HIV aware MSM was reported ($N_o$=6).

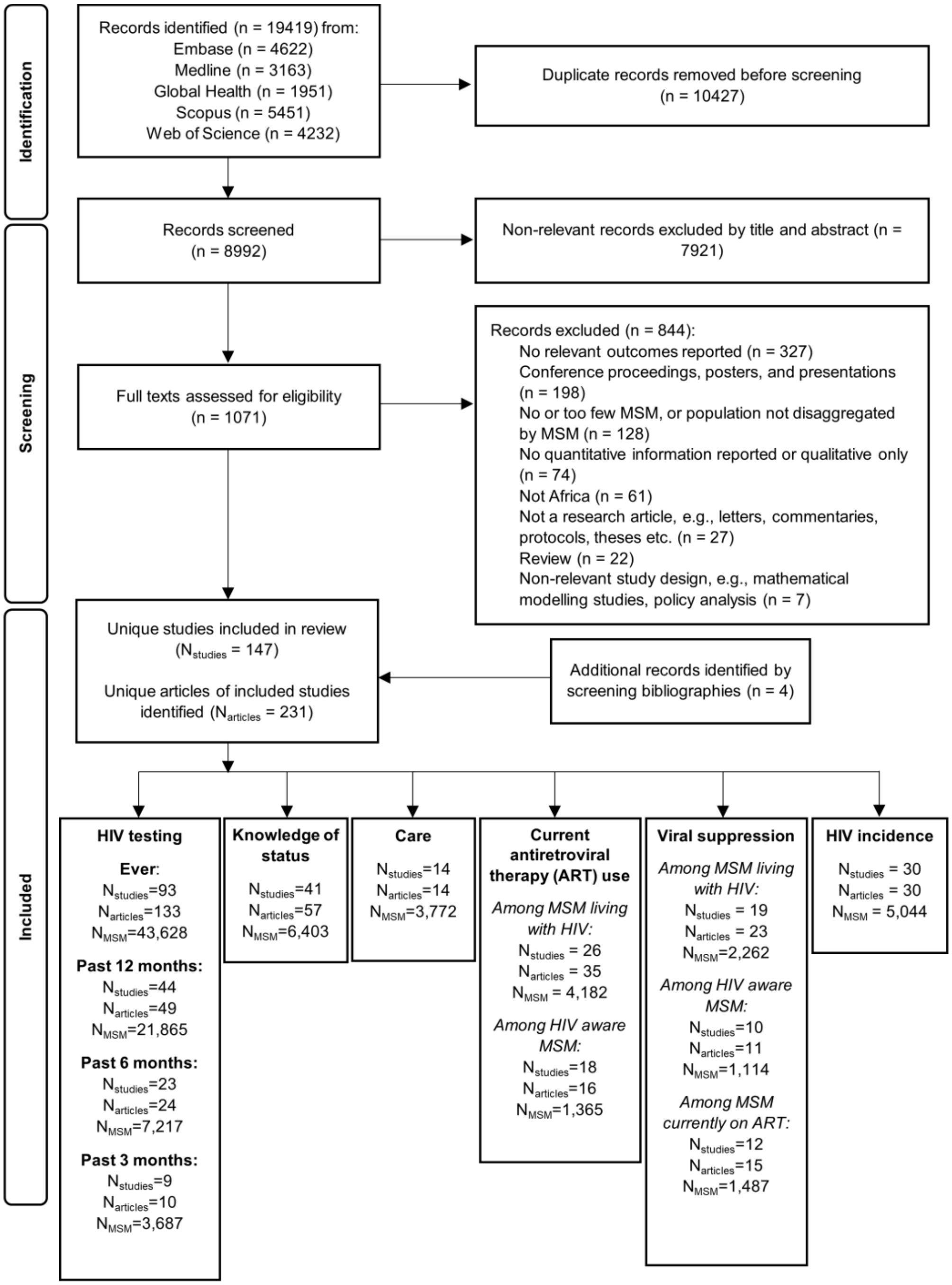


**Figure S1. PRISMA flowchart.** Screening identified 147 unique studies, reported in 231 unique articles, that were included in our analyses of HIV incidence, testing, and treatment cascade outcomes among men who have sex with men (MSM) in Africa.

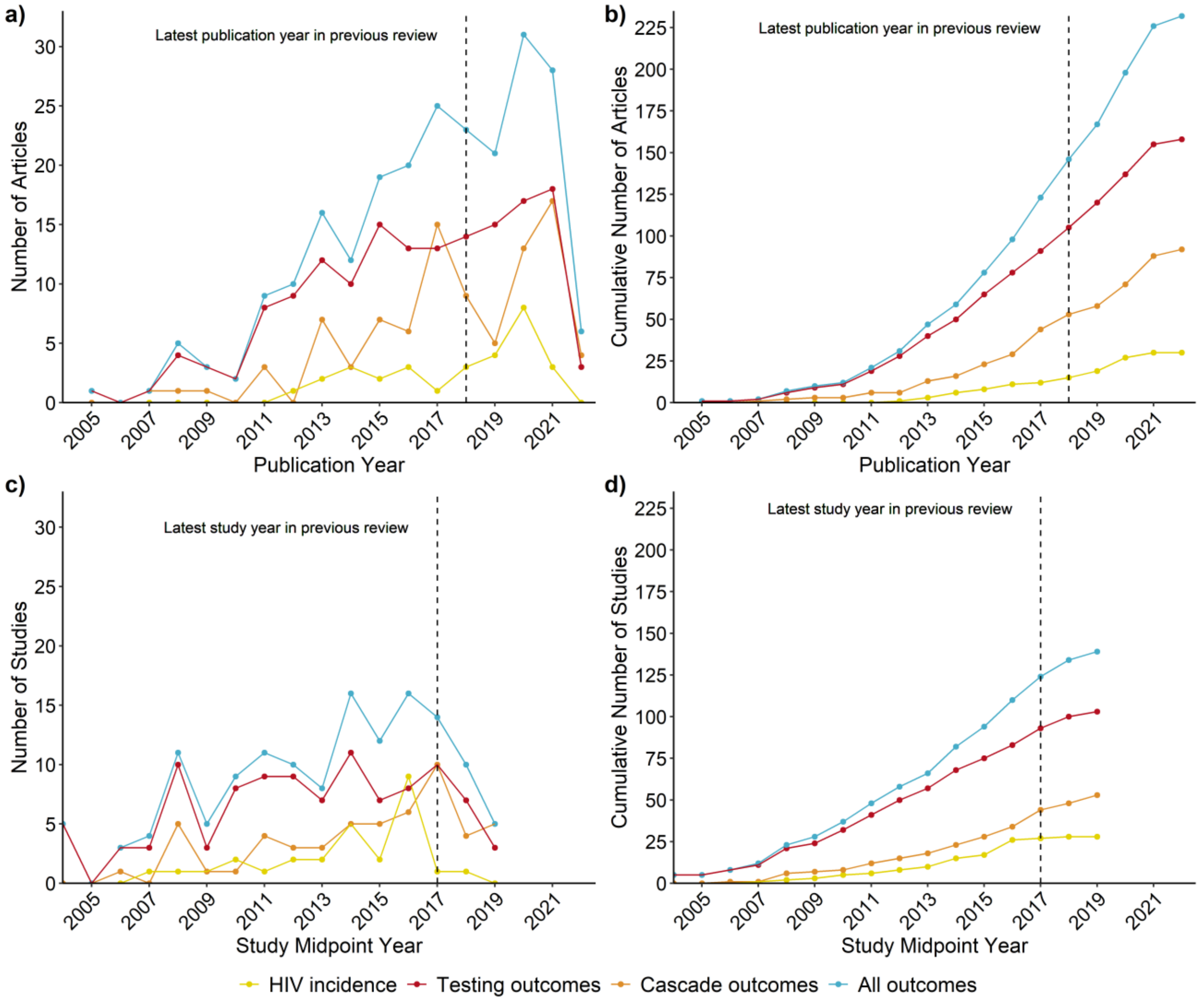


**Figure S2. Number of articles and studies over time.**

(a) The number of unique research articles published over time (by publication year), (b) the cumulative number of unique research articles published over time (by publication year), (c) the number of unique studies conducted over time (by study midpoint year), and (d) the cumulative number of studies conducted over time (by study midpoint year) included in our review reporting HIV incidence rates (yellow lines), HIV testing outcomes (red lines), and HIV treatment cascade outcomes (orange lines). In 5 studies, the study year was not reported. The dashed lines represent our previous systematic review.

| Reference | Study Year | Country | n/N | Proportion, % (95% CI) |
|---|---|---|---|---|
| **Northern Africa** | | | | |
| Elmahy, 2018 | 2016 | Egypt | 159/461 | 34.5 (30.3-38.9) |
| **Eastern Africa** | | | | |
| Kajubi, 2008 | 2004 | Uganda | 19/78 | 24.0 (15.8-34.7) |
| Ntata, 2008 | 2006 | Malawi | 57/97 | 58.8 (48.7-68.1) |
| Sanders, 2007 | 2006 | Kenya | 72/285 | 25.3 (20.6-30.6) |
| Khatib, 2017 | 2007 | Tanzania | 71/376 | 18.8 (15.2-23.1) |
| Fay, 2011 | 2008 | Malawi | 69/196 | 35.2 (28.8-42.1) |
| Hladik, 2012 | 2008 | Uganda | 66/153 | 43.4 (35.8-51.3) |
| Moller, 2015 | 2008 | Kenya | 266/561 | 47.4 (43.3-51.6) |
| Luchters, 2011 | 2008 | Kenya | 283/442 | 64.0 (59.4-68.4)* |
| Nyoni, 2021 | 2009 | Tanzania | 164/271 | 60.5 (54.6-66.2) |
| Chapman, 2011 | 2009 | Rwanda | 55/88 | 62.5 (52.0-72.0)* |
| McKinnon, 2014 | 2010 | Kenya | 435/507 | 85.8 (82.5-88.6) |
| Muraguri, 2015 | 2010 | Kenya | 402/563 | 71.4 (67.5-75.0)* |
| Mmbaga, 2012 | 2011 | Tanzania | 80/150 | 53.3 (45.3-61.2) |
| Khatib, 2017 | 2011 | Tanzania | 132/194 | 68.2 (61.3-74.4) |
| Boothe, 2021 | 2011 | Mozambique | 869/1432 | 60.7 (58.1-63.2) |
| Githuka, 2016 | 2012 | Kenya | 21/35 | 61.3 (44.4-75.8) |
| Romijnders, 2016 | 2012 | Tanzania | 230/296 | 77.7 (72.6-82.1) |
| Hladik, 2017 | 2013 | Uganda | 163/251 | 65.1 (59.0-70.7) |
| Wanyenze, 2016 | 2013 | Uganda | 76/85 | 89.4 (80.9-94.4) |
| Wirtz, 2017 | 2013 | Malawi | 52/114 | 45.9 (37.0-55.1) |
| Mmbaga, 2018 | 2014 | Tanzania | 472/753 | 62.7 (59.2-66.1) |
| Bhattacharjee, 2015 | 2014 | Kenya | 1201/1308 | 91.8 (90.2-93.2) |
| Ross, 2019 | 2015 | Tanzania | 231/231 | 99.8 (96.7-100.0)* |
| Nyblade, 2017 | 2015 | Kenya | 200/232 | 86.2 (81.1-90.1)* |
| Sandfort, 2019 | 2016 | Malawi | 109/122 | 89.3 (82.5-93.7)* |
| Sandfort, 2019 | 2016 | Kenya | 161/169 | 95.3 (90.8-97.6)* |
| Herce, 2018 | 2017 | Malawi | 86/115 | 74.8 (66.1-81.9) |
| Smith, 2021 | 2017 | Kenya | 326/347 | 94.0 (91.0-96.1) |
| Twahirwa Rwema, 2020 | 2018 | Rwanda | 670/736 | 91.0 (88.7-92.9) |
| Okoboi, 2020 | 2018 | Uganda | 204/290 | 70.3 (64.8-75.3) |
| Dijkstra, 2021 | 2019 | Kenya | 422/445 | 94.8 (92.3-96.5) |
| Bhattacharjee, 2020 | 2019 | Kenya | 1164/1200 | 97.0 (95.9-97.8) |
| Parmley, 2022 | 2019 | Zimbabwe | 990/1167 | 84.8 (82.7-86.8) |
| **Southern Africa** | | | | |
| Magesa, 2014 | NR | Tanzania | 42/50 | 84.0 (71.1-91.8) |
| Nel, 2013 | 2004 | South Africa | 754/1045 | 72.2 (69.4-74.8) |
| Lane, 2008 | 2004 | South Africa | 99/147 | 67.3 (59.4-74.4)* |
| Fay, 2011 | 2008 | Namibia | 129/217 | 59.4 (52.8-65.8) |
| Fay, 2011 | 2008 | Botswana | 97/117 | 82.9 (75.0-88.7) |
| Lane, 2011 | 2008 | South Africa | 65/172 | 37.9 (31.0-45.4) |
| Knox, 2011 | 2008 | South Africa | 203/300 | 67.7 (62.2-72.7) |
| Tun, 2012 | 2009 | South Africa | 86/120 | 71.1 (62.4-78.5) |
| Eaton, 2013 | 2010 | South Africa | 89/142 | 62.7 (54.4-70.2) |
| Wagenaar, 2012 | 2010 | South Africa | 382/439 | 87.0 (83.5-89.9) |
| Kendall, 2014 | 2011 | Angola | 125/328 | 38.1 (33.0-43.5) |
| Lane, 2014 | 2012 | South Africa | 436/605 | 72.1 (68.4-75.5) |
| Maleke, 2017 | 2012 | South Africa | 18/23 | 78.3 (57.2-90.7) |
| Siegler, 2015 | 2012 | South Africa | 33/34 | 97.1 (81.9-99.6)* |
| Batist, 2013 | 2012 | South Africa | 91/97 | 93.8 (86.9-97.2) |
| Stahlman, 2015 | 2014 | Lesotho | 366/530 | 69.1 (65.0-72.8)* |
| Sandfort, 2019 | 2016 | South Africa | 279/310 | 90.0 (86.1-92.9)* |
| Stephenson, 2021 | 2017 | SA, Namibia | 226/254 | 89.0 (84.5-92.3) |
| Fearon, 2020 | 2017 | South Africa | 171/181 | 94.5 (90.0-97.0) |
| Scheibe, 2020 | 2017 | South Africa | 726/746 | 97.3 (95.9-98.3)* |
| Herce, 2018 | 2017 | Angola | 307/713 | 43.1 (39.5-46.7)* |
| Montgomery, 2021 | 2018 | South Africa | 180/190 | 94.7 (90.5-97.1) |
| Pillay, 2020 | 2018 | South Africa | 94/96 | 97.9 (92.1-99.5) |
| Jobson, 2018 | NR | South Africa | 272/316 | 86.1 (81.8-89.5)* |

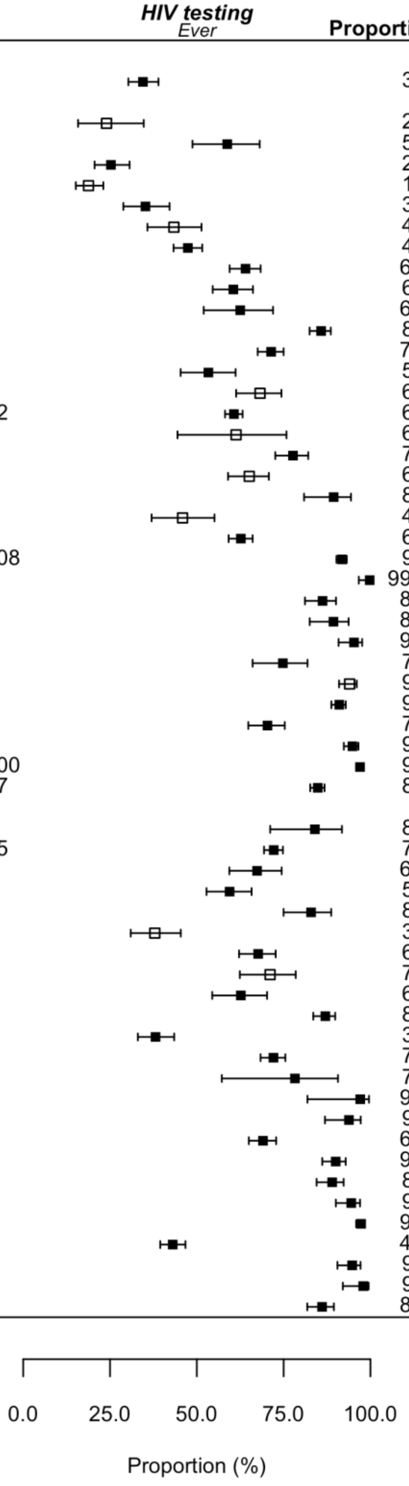

**Ever HIV testing continued…**

| Reference | Study Year | Country | n/N | Proportion, % (95% CI) |
|---|---|---|---|---|
| **Central Africa** | | | | |
| Lorente, 2012 | 2008 | Cameroon | 134/165 | 81.2 (74.5-86.5) |
| Holland, 2015 | 2011 | Cameroon | 413/511 | 80.8 (77.2-84.0) |
| Coulaud, 2016 | 2014 | Burundi | 49/51 | 96.1 (85.6-99.0)* |
| Lillie, 2021 | 2018 | Burundi | 91/363 | 25.1 (20.9-29.8) |
| Longo, 2018 | NR | CAR | 36/396 | 9.1 (6.6-12.3) |
| **Western Africa** | | | | |
| Wade, 2005 | 2004 | Senegal | 50/463 | 10.8 (8.3-14.0) |
| Vuylsteke, 2012 | 2007 | Cote d'Ivoire | 68/96 | 70.8 (61.0-79.0) |
| Eluwa, 2019 | 2007 | Nigeria | 305/897 | 34.0 (31.0-37.2) |
| Stromdahl, 2019 | 2008 | Nigeria | 180/276 | 65.2 (59.4-70.6) |
| Gyamerah, 2020 | 2010 | Ghana | 571/1382 | 41.3 (38.7-43.9)* |
| Adebajo, 2014 | 2010 | Nigeria | 389/708 | 54.9 (51.3-58.6)* |
| Eluwa, 2019 | 2010 | Nigeria | 778/1327 | 58.6 (55.9-61.2) |
| Ekouevi, 2014 | 2011 | Togo | 259/408 | 63.5 (58.7-68.0)* |
| Bakai, 2016 | 2011 | Togo | 446/708 | 63.0 (59.4-66.5)* |
| Hakim, 2015 | 2011 | Cote d'Ivoire | 165/263 | 62.6 (56.6-68.2) |
| Drame, 2013 | 2012 | Senegal | 103/117 | 86.6 (79.2-91.6) |
| Nelson, 2015 | 2012 | Ghana | 93/136 | 68.4 (60.1-75.6) |
| Ruisenor-Escudero, 2019 | 2013 | Togo | 483/683 | 70.7 (67.2-74.0) |
| Girault, 2015 | 2013 | Ghana | 100/166 | 60.2 (52.6-67.4) |
| Goodman, 2016 | 2013 | Burkina Faso | 506/670 | 75.5 (72.1-78.6)* |
| Abubakari, 2021 | 2014 | Ghana | 47/57 | 82.5 (70.4-90.3) |
| Eluwa, 2019 | 2014 | Nigeria | 2217/3432 | 64.6 (63.0-66.2) |
| Tobin-West, 2017 | 2014 | Nigeria | 70/101 | 69.3 (59.7-77.5) |
| Lahuerta, 2018 | 2014 | Mali | 169/236 | 71.6 (65.5-77.0)* |
| Ulanja, 2019 | 2015 | Cote d'Ivoire | 1042/1301 | 80.1 (77.8-82.2) |
| Teclessou, 2017 | 2015 | Togo | 317/466 | 68.0 (63.7-72.1)* |
| Yaya, 2021 | 2016 | Togo | 129/160 | 80.6 (73.8-86.0)* |
| Yaya, 2021 | 2016 | Mali | 233/295 | 79.0 (74.0-83.3)* |
| Yaya, 2021 | 2016 | Cote d'Ivoire | 131/193 | 67.9 (61.0-74.1)* |
| Ramadhani, 2020 | 2016 | Nigeria | 1745/2123 | 82.2 (80.5-83.8) |
| Yaya, 2021 | 2017 | Burkina Faso | 131/168 | 78.0 (71.1-83.6)* |
| Sadio, 2019 | 2017 | Togo | 595/668 | 89.1 (86.5-91.2)* |
| Tun, 2018 | 2017 | Nigeria | 262/319 | 82.1 (77.5-86.0) |
| Inghels, 2021 | 2018 | Cote d'Ivoire | 477/518 | 92.1 (89.4-94.1) |
| Lieber, 2018 | NR | Liberia | 83/107 | 77.6 (68.7-84.5) |

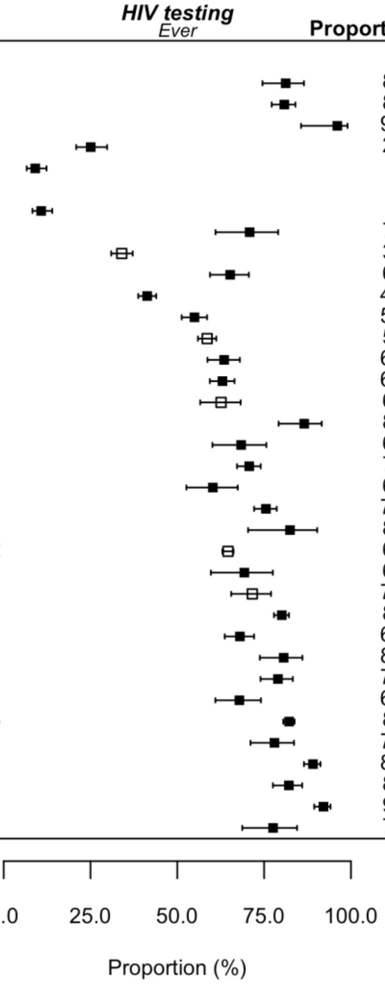


***** observation calculated using available data reported within article

**Figure S3. Forest plot of study proportions of men who have sex with men (MSM) ever tested for HIV, by region of Africa.** Studies reported crude proportions (filled squares) or proportions adjusted for sampling design (e.g., respondent driven sampling, cluster, time-location sampling; unfilled squares).

| Reference | Study Year | Country | n/N | Proportion (95% CI) |
|---|---|---|---|---|
| **Northern Africa** | | | | |
| Valadez, 2013 | 2010 | Libya | 79/174 | 45.6 (38.3-53.0) |
| **Eastern Africa** | | | | |
| Adam, 2009 | 2004 | Mauritius | 8/50 | 16.0 (8.2-28.9) |
| Khatib, 2017 | 2007 | Tanzania | 39/343 | 11.3 (8.4-15.1) |
| Muraguri, 2015 | 2010 | Kenya | 268/563 | 47.6 (43.5-51.7)* |
| Khatib, 2017 | 2011 | Tanzania | 139/251 | 55.3 (49.1-61.3) |
| Horth, 2015 | 2011 | Mozambique | 558/1412 | 39.5 (37.0-42.1) |
| Hladik, 2017 | 2013 | Uganda | 104/147 | 70.9 (63.0-77.7) |
| Wirtz, 2017 | 2013 | Malawi | 19/76 | 24.9 (16.4-35.8) |
| Shangani, 2017 | 2014 | Kenya | 66/89 | 74.2 (64.1-82.2) |
| Ntale, 2019 | 2015 | Rwanda | 385/504 | 76.4 (72.5-79.9) |
| Sandfort, 2019 | 2016 | Malawi | 91/122 | 74.6 (66.1-81.5)* |
| Virkud, 2020 | 2016 | Multiple† | 57/85 | 67.3 (56.6-76.4) |
| Sandfort, 2019 | 2016 | Kenya | 147/169 | 87.0 (81.0-91.3)* |
| Dijkstra, 2021 | 2019 | Kenya | 313/445 | 70.3 (65.9-74.4)* |
| Bhattacharjee, 2020 | 2019 | Kenya | 1008/1184 | 85.1 (83.0-87.0) |
| **Southern Africa** | | | | |
| Burrel, 2010 | 2008 | South Africa | 388/534 | 72.7 (68.7-76.3) |
| Knox, 2011 | 2008 | South Africa | 120/290 | 41.4 (35.8-47.1) |
| Baral, 2011 | 2009 | Lesotho | 128/235 | 54.5 (48.1-60.7) |
| Rao, 2017 | 2011 | eSwatini | 152/290 | 52.4 (46.7-58.1) |
| Kendall, 2014 | 2011 | Angola | 104/328 | 31.7 (26.9-36.9)* |
| Rebe, 2015 | 2012 | South Africa | 107/200 | 53.5 (46.6-60.3) |
| Rao, 2017 | 2014 | eSwatini | 121/136 | 89.0 (82.5-93.2) |
| Lippman, 2018 | 2015 | South Africa | 84/127 | 66.1 (57.5-73.8)* |
| Sandfort, 2019 | 2016 | South Africa | 215/310 | 69.4 (64.0-74.2)* |
| Stephenson, 2022 | 2017 | SA, Namibia | 216/440 | 49.1 (44.4-53.8) |
| Fearon, 2020 | 2017 | South Africa | 118/157 | 75.2 (67.8-81.3) |
| Pillay, 2020 | 2018 | South Africa | 90/96 | 93.8 (86.8-97.2)* |
| **Central Africa** | | | | |
| Park, 2014 | 2011 | Cameroon | 301/503 | 59.8 (55.5-64.0)* |
| Rao, 2017 | 2013 | Cameroon | 188/212 | 88.7 (83.7-92.3) |
| Coulaud, 2016 | 2014 | Burundi | 44/51 | 86.3 (73.9-93.3)* |
| Bowring, 2019 | 2016 | Cameroon | 729/1322 | 55.1 (52.5-57.8)* |
| **Western Africa** | | | | |
| Adam, 2009 | 2006 | Mauritania | 4/26 | 15.4 (5.9-34.5) |
| Eluwa, 2019 | 2007 | Nigeria | 217/298 | 72.9 (67.6-77.6) |
| Gyamerah, 2020 | 2010 | Ghana | 426/1382 | 30.8 (28.4-33.3) |
| Eluwa, 2019 | 2010 | Nigeria | 544/706 | 77.1 (73.9-80.1) |
| Aho, 2014 | 2011 | Cote d'Ivoire | 99/310 | 32.1 (27.1-37.5) |
| Nelson, 2015 | 2012 | Ghana | 80/92 | 87.0 (78.4-92.4)* |
| Girault, 2015 | 2013 | Ghana | 64/166 | 38.6 (31.5-46.2) |
| Eluwa, 2019 | 2014 | Nigeria | 1746/2213 | 78.9 (77.1-80.5) |
| Lahuerta, 2018 | 2014 | Mali | 114/241 | 47.1 (40.9-53.4) |
| Bouscaillou, 2016 | 2014 | Cote d'Ivoire | 15/40 | 37.5 (24.0-53.2) |
| Tun, 2018 | 2017 | Nigeria | 147/319 | 46.1 (40.7-51.6)* |
| Diabate, 2020 | 2018 | Cote d'Ivoire | 176/201 | 87.6 (82.2-91.5) |
| Ahouada, 2020 | 2018 | Benin | 392/400 | 98.0 (96.1-99.0) |

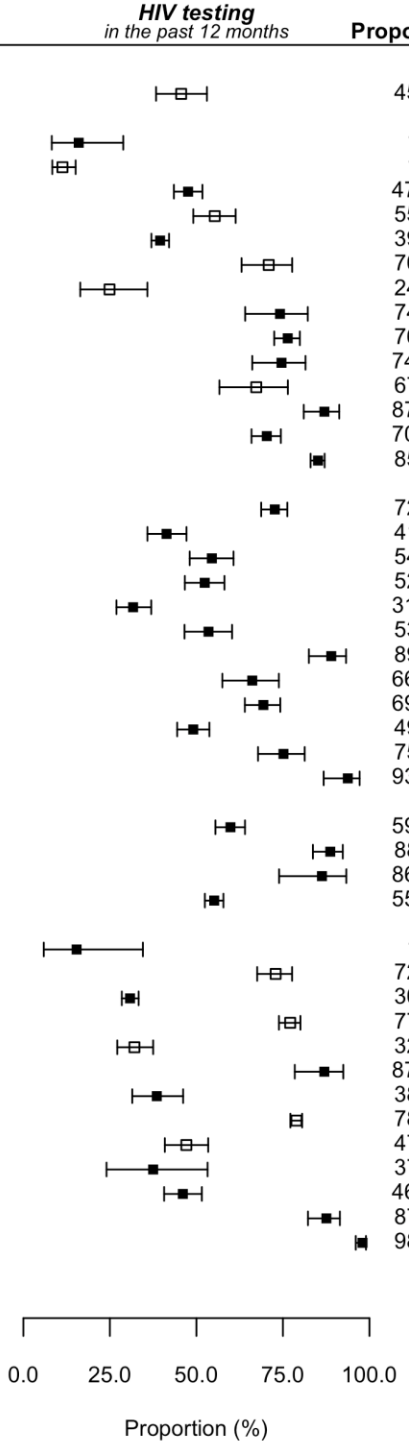


***** observation calculated using available data reported within article

**Figure S4. Forest plot of study proportions of men who have sex with men (MSM) tested for HIV in the past 12 months, by region of Africa.** Studies reported crude proportions (filled squares) or proportions adjusted for sampling design (e.g., respondent driven sampling, cluster, time-location sampling; unfilled squares).

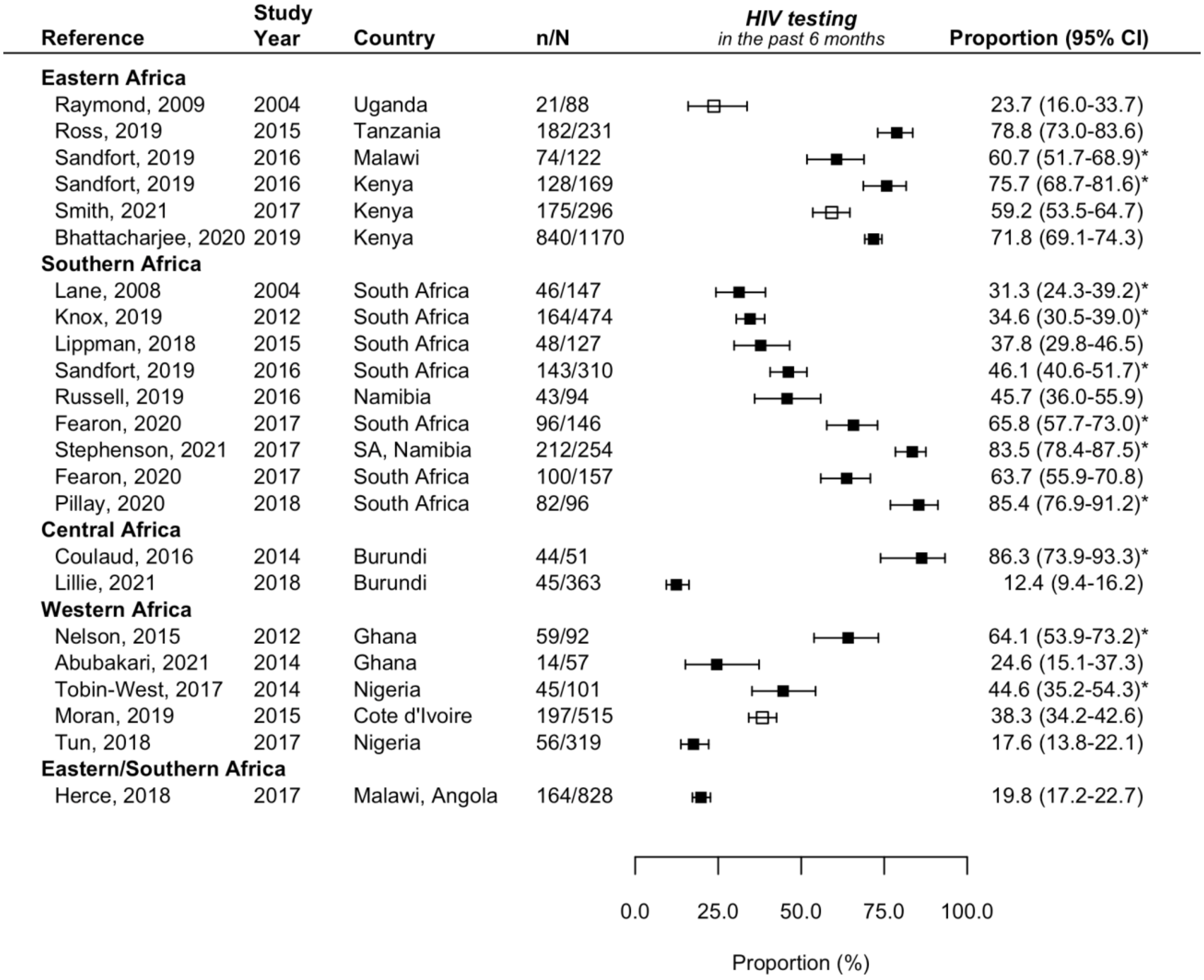


***** observation calculated using available data reported within article

**Figure S5. Forest plot of study proportions of men who have sex with men (MSM) tested for HIV in the past 6 months, by region of Africa.** Studies reported crude proportions (filled squares) or proportions adjusted for sampling design (e.g., respondent driven sampling, cluster, time-location sampling; unfilled squares).

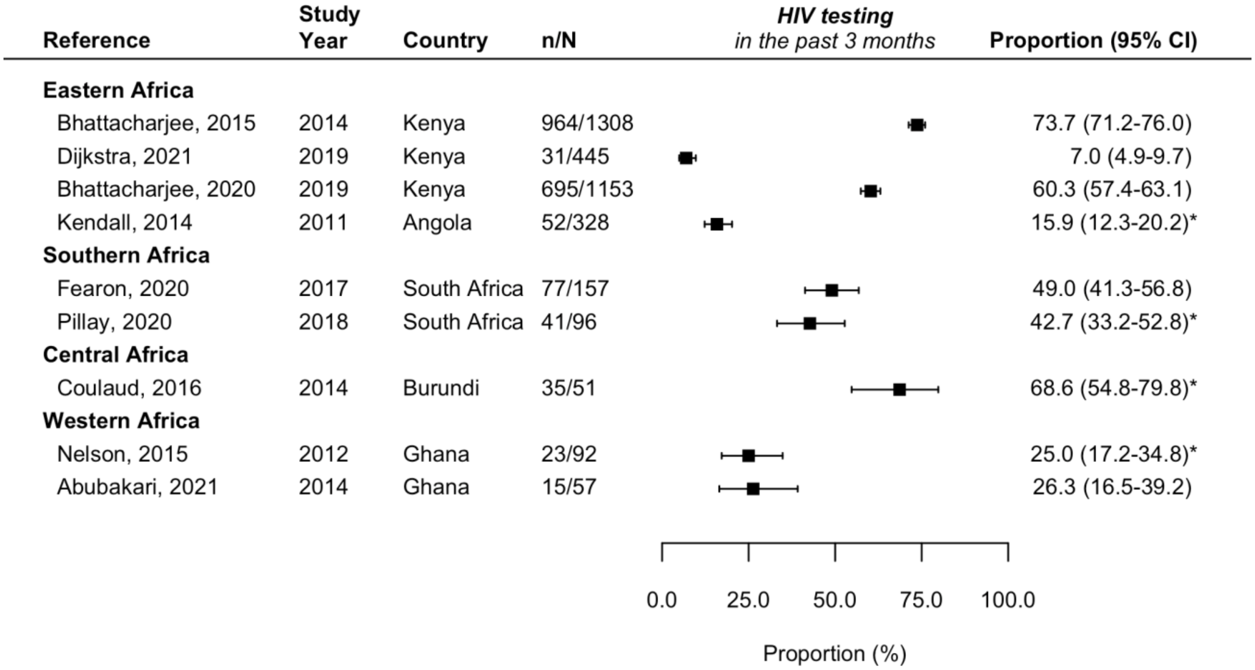


***** observation calculated using available data reported within article

**Figure S6. Forest plot of study proportions of men who have sex with men (MSM) tested for HIV in the past 3 months, by region of Africa.** Studies reported crude proportions (filled squares) or proportions adjusted for sampling design (e.g., respondent driven sampling, cluster, time-location sampling; unfilled squares).

| Reference | Study Year | Country | n/N | Proportion (95% CI) |
|---|---|---|---|---|
| **Eastern Africa** | | | | |
| Sanders, 2007 | 2006 | Kenya | 7/70 | 10.0 (4.8-19.5)* |
| Baral, 2009 | 2008 | Malawi | 2/43 | 4.7 (1.2-16.8) |
| Muraguri, 2015 | 2010 | Kenya | 49/144 | 34.0 (26.8-42.1)* |
| Boothe, 2021 | 2011 | Mozambique | 10/114 | 8.8 (4.8-15.5) |
| Ross, 2014 | 2012 | Tanzania | 5/62 | 8.1 (3.4-18.0) |
| Hladik, 2017 | 2013 | Uganda | 16/79 | 20.3 (12.8-30.5) |
| Wirtz, 2017 | 2013 | Malawi | 3/311 | 0.9 (0.3-2.9) |
| Sandfort, 2019 | 2016 | Malawi | 13/27 | 48.1 (30.4-66.4)* |
| Sandfort, 2019 | 2016 | Kenya | 23/28 | 82.1 (63.6-92.4)* |
| Korhonen, 2018 | 2016 | Kenya | 264/1476 | 17.9 (16.0-19.9) |
| Herce, 2018 | 2017 | Malawi | 1/2 | 50.0 (5.9-94.1) |
| Smith, 2021 | 2017 | Kenya | 93/121 | 76.6 (68.2-83.3) |
| Twahirwa Rwema, 2020 | 2018 | Rwanda | 45/74 | 60.8 (49.3-71.2) |
| Okoboi, 2021 | 2018 | Uganda | 3/12 | 25.0 (8.3-55.2) |
| Dijkstra, 2021 | 2019 | Kenya | 26/56 | 46.4 (33.9-59.4) |
| Bhattacharjee, 2020 | 2019 | Kenya | 76/201 | 37.8 (31.4-44.7) |
| Harris, 2022 | 2019 | Zimbabwe | 180/248 | |
| **Southern Africa** | | | | |
| Baral, 2009 | 2008 | Namibia | 16/27 | 59.3 (40.3-75.8) |
| Baral, 2009 | 2008 | Botswana | 4/23 | 17.4 (6.7-38.2) |
| Lane, 2011 | 2008 | South Africa | 8/69 | 11.6 (5.9-21.5)* |
| Baral, 2011 | 2009 | South Africa | 3/50 | 6.0 (1.9-17.0) |
| Stahlman, 2016 | 2011 | eSwatini | 2/5 | 42.9 (10.5-82.7) |
| Kendall, 2014 | 2011 | Angola | 10/27 | 37.0 (21.2-56.2)* |
| Lane, 2014 | 2012 | South Africa | 40/172 | 23.3 (17.5-30.1) |
| Stahlman, 2016 | 2014 | Lesotho | 16/30 | 53.7 (36.1-70.4) |
| van Liere, 2017 | 2015 | South Africa | 36/43 | 83.7 (69.6-92.0)* |
| Sandfort, 2019 | 2016 | South Africa | 67/128 | 52.3 (43.7-60.8)* |
| Sullivan, 2020 | 2016 | South Africa | 63/125 | 50.4 (41.7-59.1) |
| Fearon, 2020 | 2017 | South Africa | 76/117 | 65.0 (55.9-73.0)* |
| Fearon, 2020 | 2017 | South Africa | 21/36 | |
| Scheibe, 2020 | 2017 | South Africa | 274/320 | 85.6 (81.3-89.1)* |
| **Central Africa** | | | | |
| Herce, 2018 | 2017 | Angola | 4/20 | 20.0 (7.7-42.8) |
| Bowring, 2019 | 2016 | Cameroon | 115/272 | 42.3 (36.5-48.2)* |
| **Western Africa** | | | | |
| Mason, 2013 | 2011 | The Gambia | 1/20 | 5.0 (0.7-28.2)* |
| Hakim, 2015 | 2011 | Cote d'Ivoire | 6/46 | 13.6 (6.3-26.8) |
| Drame, 2013 | 2012 | Senegal | 20/41 | 48.8 (34.0-63.7)* |
| Holland, 2016 | 2013 | Togo | 10/67 | 14.9 (8.2-25.6) |
| Holland, 2016 | 2013 | Burkina Faso | 10/32 | 31.2 (17.7-49.0)* |
| Lyons, 2017 | 2014 | Senegal | 29/219 | 13.2 (9.4-18.4) |
| Hakim, 2017 | 2014 | Mali | 10/79 | 13.3 (7.4-22.7) |
| Tiamiyu, 2020 | 2015 | Nigeria | 529/984 | 53.8 (50.6-56.9) |

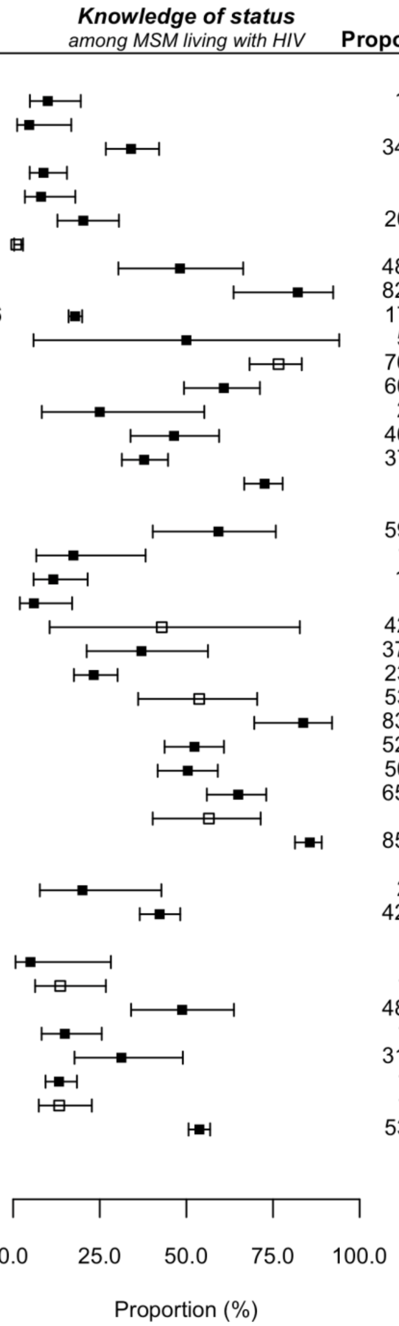


***** observation calculated using available data reported within article

**Figure S7. Forest plot of study proportions of men who have sex with men (MSM) living with HIV who know their status (HIV aware MSM), by region of Africa.** Studies reported crude proportions (filled squares) or proportions adjusted for sampling design (e.g., respondent driven sampling, cluster, time-location sampling; unfilled squares).

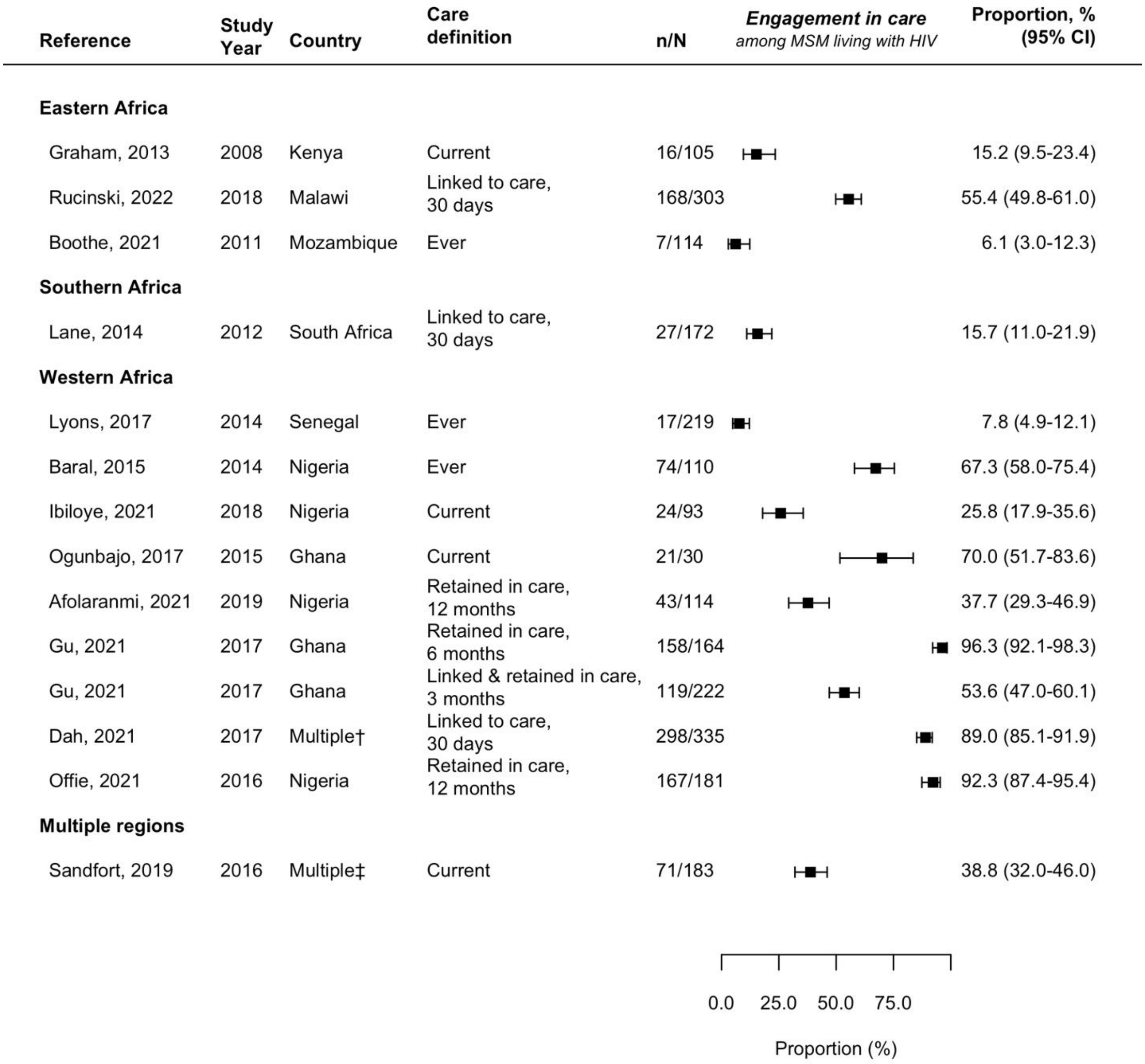


***** observation calculated using available data reported within article

† includes Burkina Faso, Cote d'Ivoire, Mali, Togo

‡ includes Kenya, Malawi, South Africa

**Figure S8. Forest plot of study proportions of men who have sex with men (MSM) living with HIV engaged in care other than current ART use, by region of Africa.**

**(a)**

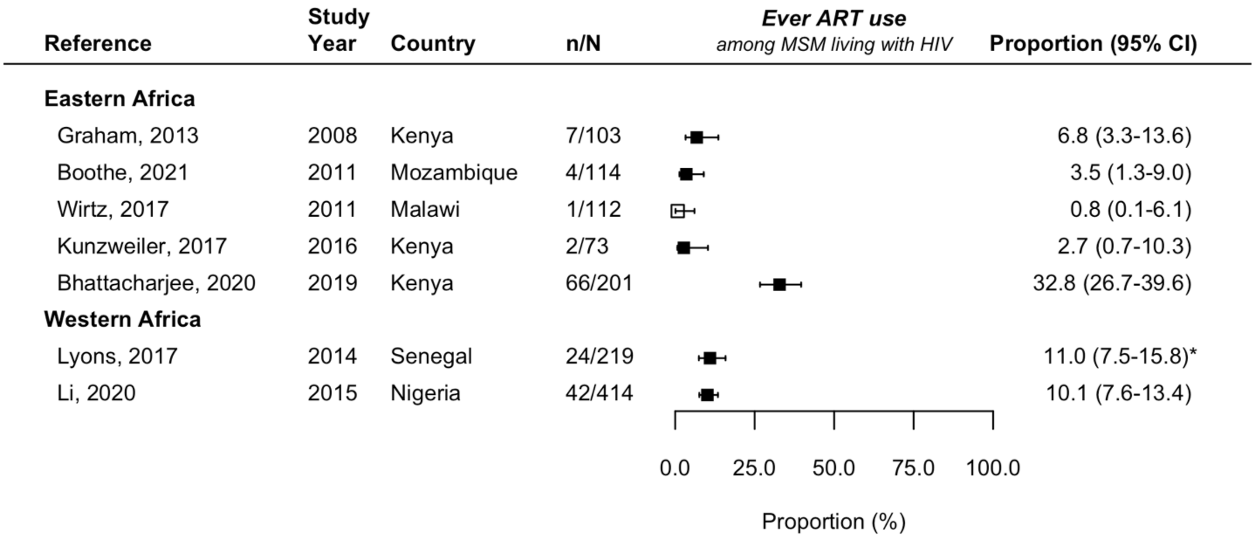


***** observation calculated using available data reported within article

**(b)**

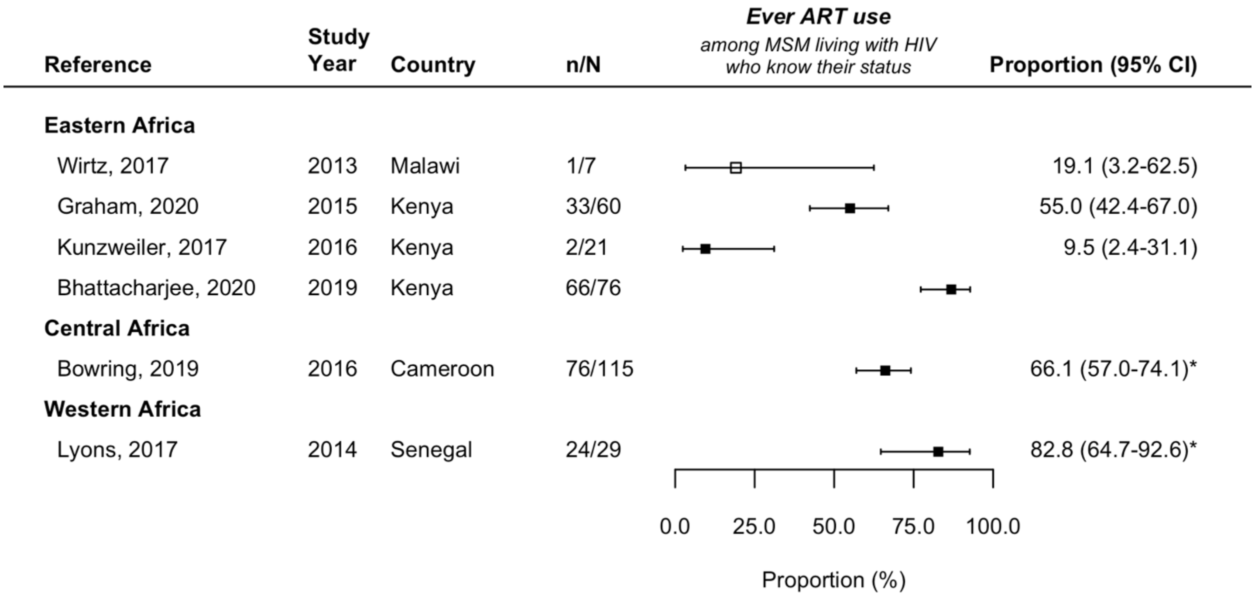


***** observation calculated using available data reported within article

**Figure S9. Forest plot of study proportions of men who have sex with men (MSM) ever on ART, by region of Africa.** Ever ART use among (a) MSM living with HIV, and (b) HIV aware MSM. Studies reported crude proportions (filled squares) or proportions adjusted for sampling design (e.g., respondent driven sampling, cluster, time-location sampling; unfilled squares).

| Reference | Study Year | Country | n/N | Proportion (95% CI) |
|---|---|---|---|---|
| **Eastern Africa** | | | | |
| Boothe, 2021 | 2011 | Mozambique | 4/114 | 3.5 (1.3-9.0) |
| Hladik, 2017 | 2013 | Uganda | 12/79 | 15.2 (8.8-24.9) |
| Sandfort, 2019 | 2016 | Malawi | 10/27 | 37.0 (21.2-56.2)* |
| Sandfort, 2019 | 2016 | Kenya | 19/28 | 67.9 (48.9-82.4)* |
| Kunzweiler, 2019 | 2016 | Kenya | 8/63 | 12.7 (6.5-23.4) |
| Smith, 2021 | 2017 | Kenya | 83/126 | 65.3 (56.6-73.1) |
| Twahirwa Rwema, 2020 | 2018 | Rwanda | 44/74 | 59.5 (48.0-70.0) |
| Dijkstra, 2021 | 2019 | Kenya | 25/56 | 44.6 (32.3-57.7)* |
| Bhattacharjee, 2020 | 2019 | Kenya | 65/201 | 32.3 (26.2-39.1) |
| Harris, 2022 | 2019 | Zimbabwe | 174/248 | 70.2 (64.2-75.5) |
| Gebrebrhan, 2021 | NR | Kenya | 21/31 | 67.7 (49.7-81.7) |
| **Southern Africa** | | | | |
| Lane, 2014 | 2012 | South Africa | 21/172 | 12.2 (8.1-18.0) |
| Rebe, 2015 | 2012 | South Africa | 46/88 | 52.3 (41.9-62.5) |
| Rees, 2017 | 2015 | South Africa | 399/646 | 61.8 (58.0-65.4) |
| Sandfort, 2019 | 2016 | South Africa | 34/128 | 26.6 (19.6-34.9)* |
| Fearon, 2020 | 2017 | South Africa | 39/118 | 33.1 (25.2-42.0)* |
| Fearon, 2020 | 2017 | South Africa | 14/48 | 30.0 (18.8-44.3) |
| Scheibe, 2020 | 2017 | South Africa | 255/274 | |
| **Central Africa** | | | | |
| Bouassa, 2018 | 2010 | CAR | 10/29 | 34.5 (19.7-53.1)* |
| **Western Africa** | | | | |
| Ndiaye, 2013 | 2007 | Senegal | 9/97 | 9.3 (4.9-16.9) |
| Holland, 2016 | 2013 | Togo | 4/67 | 6.0 (2.3-14.9)* |
| Holland, 2016 | 2013 | Burkina Faso | 5/32 | 15.6 (6.7-32.5)* |
| Lyons, 2017 | 2014 | Senegal | 22/219 | 10.0 (6.7-14.8)* |
| Ramadhani, 2020 | 2016 | Nigeria | 638/865 | 73.8 (70.7-76.6)* |
| Yaya, 2021 | 2017 | Multiple† | 164/206 | 79.6 (73.6-84.6) |
| Koyalta, 2021 | 2019 | Mali | 14/16 | 87.5 (61.4-96.9) |

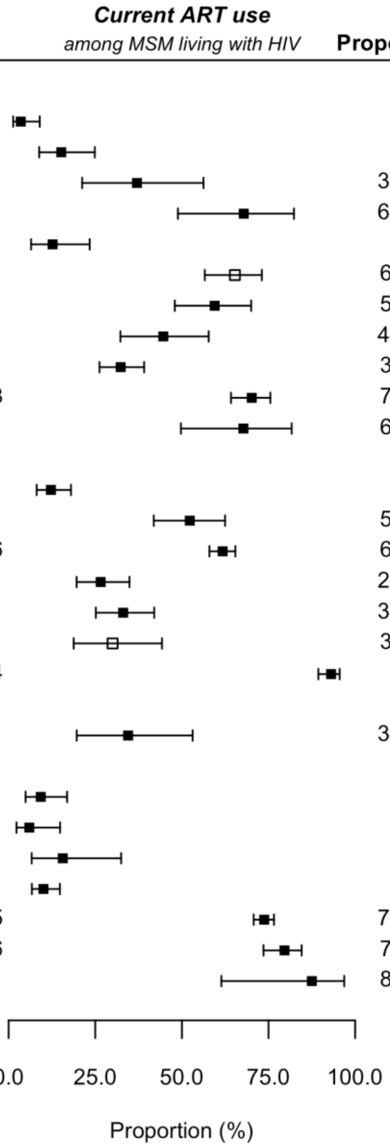


***** observation calculated using available data reported within article

† includes Burkina Faso, Cote d'Ivoire, Mali, and Togo.

**Figure S10. Forest plot of study proportions of men who have sex with men (MSM) living with HIV currently on antiretroviral therapy (ART), by region of Africa.** Studies reported crude proportions (filled squares) or proportions adjusted for sampling design (e.g., respondent driven sampling, cluster, time-location sampling; unfilled squares).

| Reference | Study Year | Country | n/N | Proportion (95% CI) |
|---|---|---|---|---|
| **Eastern Africa** | | | | |
| Hladik, 2017 | 2013 | Uganda | 12/16 | 75.0 (49.2-90.3) |
| Sandfort, 2019 | 2016 | Malawi | 10/13 | 76.9 (47.8-92.4)* |
| Sandfort, 2019 | 2016 | Kenya | 19/23 | 82.6 (61.8-93.3)* |
| Kunzweiler, 2019 | 2016 | Kenya | 8/18 | 44.4 (24.0-67.0)* |
| Smith, 2021 | 2017 | Kenya | 106/122 | 86.9 (79.7-91.8) |
| Twahirwa Rwema, 2020 | 2018 | Rwanda | 44/45 | 97.8 (85.8-99.7) |
| Dijkstra, 2021 | 2019 | Kenya | 25/26 | 96.2 (77.2-99.5) |
| Bhattacharjee, 2020 | 2019 | Kenya | 65/76 | 85.5 (75.7-91.8) |
| Harris, 2022 | 2019 | Zimbabwe | 174/180 | |
| **Southern Africa** | | | | |
| Lane, 2014 | 2012 | South Africa | 21/40 | 52.5 (37.3-67.3) |
| Sandfort, 2019 | 2016 | South Africa | 34/67 | 50.7 (39.0-62.5)* |
| Fearon, 2020 | 2017 | South Africa | 12/23 | 53.2 (33.3-72.1) |
| Cloete, 2008 | NR | South Africa | 25/92 | 27.2 (19.1-37.1) |
| **Western Africa** | | | | |
| Holland, 2016 | 2013 | Togo | 4/10 | 40.0 (15.8-70.3)* |
| Holland, 2016 | 2013 | Burkina Faso | 5/12 | 41.7 (18.5-69.2)* |
| Lyons, 2017 | 2014 | Senegal | 22/29 | 75.9 (57.3-88.0)* |
| Hakim, 2018 | 2014 | Mali | 3/5 | 61.2 (19.2-91.3) |
| Tiamiyu, 2020 | 2015 | Nigeria | 226/493 | 45.8 (41.5-50.3)* |

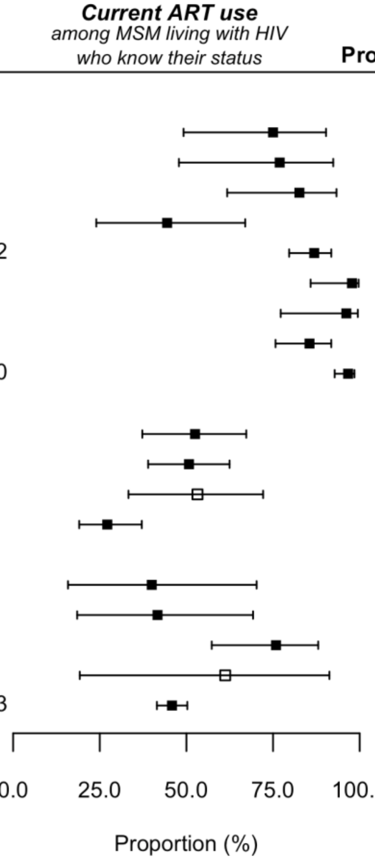


***** observation calculated using available data reported within article

**Figure S11. Forest plot of study proportions of HIV aware men who have sex with men (MSM) currently on antiretroviral therapy (ART), by region of Africa.** Studies reported crude proportions (filled squares) or proportions adjusted for sampling design (e.g., respondent driven sampling, cluster, time-location sampling; unfilled squares).

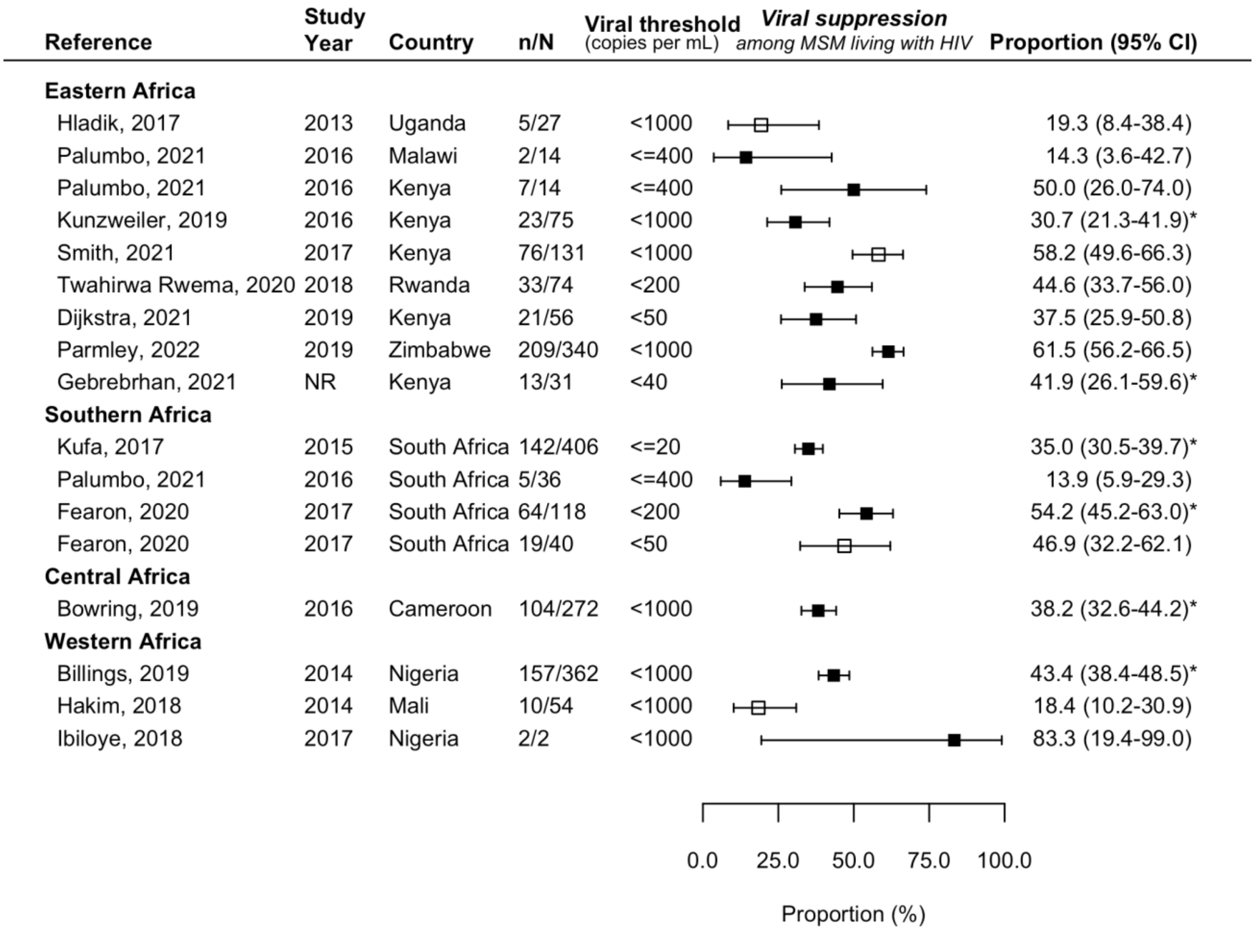


***** observation calculated using available data reported within article

**Figure S12. Forest plot of study proportions of men who have sex with men (MSM) living with HIV virally suppressed, by region of Africa.** Studies reported crude proportions (filled squares) or proportions adjusted for sampling design (e.g., respondent driven sampling, cluster, time-location sampling; unfilled squares).

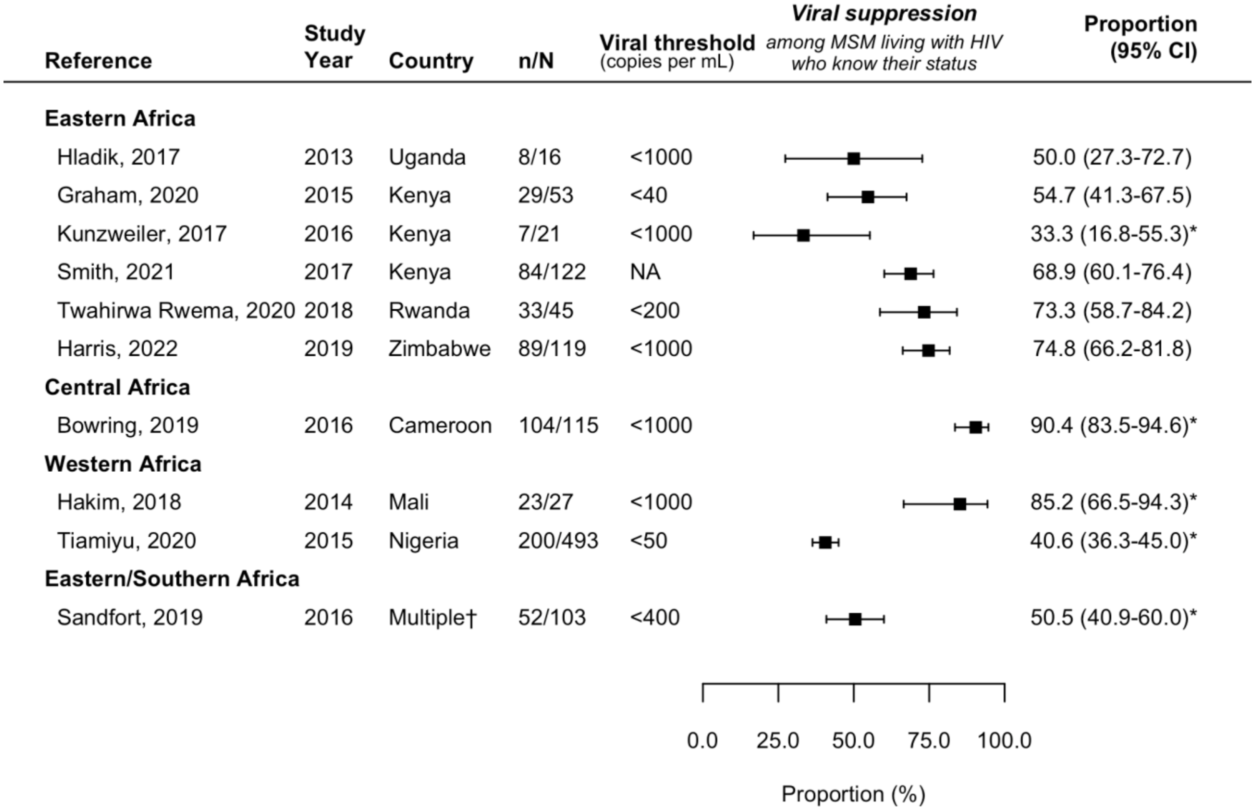


***** observation calculated using available data reported within article

† includes Kenya, Malawi, and South Africa.

**Figure S13. Forest plot of study proportions of HIV aware men who have sex with men (MSM) virally suppressed, by region of Africa.** Studies reported crude proportions (filled squares) or proportions adjusted for sampling design (e.g., respondent driven sampling, cluster, time-location sampling; unfilled squares).

| Reference | Study Year | Country | n/N | Viral threshold (copies per mL) | *Viral suppression* *among MSM on ART* | Proportion (95% CI) |
|---|---|---|---|---|---|---|
| **Eastern Africa** | | | | | | |
| Hladik, 2017 | 2013 | Uganda | 7/12 | <1000 | | 58.3 (30.8-81.5) |
| Smith, 2021 | 2017 | Kenya | 84/106 | NA | | 79.2 (70.5-85.9) |
| Twahirwa Rwema, 2020 | 2018 | Rwanda | 33/44 | <200 | | 75.0 (60.3-85.6) |
| Dijkstra, 2021 | 2019 | Kenya | 21/24 | <50 | | 87.5 (67.6-95.9) |
| Harris, 2022 | 2019 | Zimbabwe | 151/174 | <1000 | | 86.8 (80.9-91.1) |
| Gebrebrhan, 2021 | NR | Kenya | 13/18 | <40 | | 72.2 (48.1-87.9) |
| **Southern Africa** | | | | | | |
| Fearon, 2020 | 2017 | South Africa | 9/11 | <50 | | 77.3 (46.0-93.2) |
| **Western Africa** | | | | | | |
| Lyons, 2017 | 2014 | Senegal | 14/22 | <1000 | | 63.6 (42.3-80.7) |
| Hakim, 2018 | 2014 | Mali | 23/23 | <1000 | | 97.9 (74.1-99.9) |
| Ramadhani, 2020 | 2016 | Nigeria | 377/488 | <1000 | | 77.3 (73.3-80.8)* |
| Ibiloye, 2021 | 2017 | Nigeria | 466/474 | <1000 | | 98.3 (96.7-99.2) |
| **Eastern/Southern Africa** | | | | | | |
| Sandfort, 2019 | 2016 | Multiple† | 52/63 | <400 | | 82.5 (71.2-90.1) |

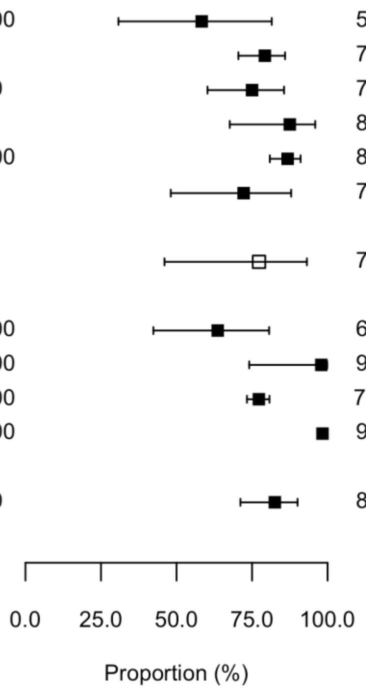


***** observation calculated using available data reported within article

† includes Kenya, Malawi, and South Africa.

**Figure S14. Forest plot of study proportions of men who have sex with men (MSM) currently on antiretroviral therapy (ART) virally suppressed, by region of Africa.** Studies reported crude proportions (filled squares) or proportions adjusted for sampling design (e.g., respondent driven sampling, cluster, time-location sampling; unfilled squares).

| Reference | Study years | Country | Events | Person years | Incidence per 100py (95% CI) |
|---|---|---|---|---|---|
| **Eastern Africa** | | | | | |
| Kamali, 2015 | 2005–2008 | Kenya | 25 | 304 | 8.2 (5.6, 12.2) |
| Kamali, 2015 | 2009–2012 | Kenya | 48 | 692 | 6.9 (5.2, 9.2) |
| Kamali, 2015 | 2006–2006 | Kenya | 5 | 64 | 7.8 (3.3, 18.8) |
| Kamali, 2015 | 2006–2009 | Kenya | 14 | 274 | 5.1 (3.0, 8.6) |
| Kimani, 2019 | 2016–2017 | Kenya | 6 | 162.1 | 3.7 (1.7, 8.2) |
| McKinnon, 2014 | 2009–2012 | Kenya | 28 | 256 | 10.9 (7.6, 15.8) |
| Robb, 2016 | 2009–2015 | Uganda | 4 | 111 | 3.6 (1.4, 9.6) |
| Sandfort, 2021 | 2015–2017 | Malawi | 1 | 74.4 | 1.3 (0.2, 9.5) |
| Sandfort, 2021 | 2015–2017 | Kenya | 3 | 80.1 | 3.7 (1.2, 11.6) |
| Wahome, 2020 | 2017–2019 | Kenya | 9 | 233.7 | 3.9 (2.0, 7.4) |
| Wirtz, 2015 | 2012–2012 | Malawi | 7 | 79.6 | 8.8 (4.2, 18.5)* |
| Wirtz, 2015 | 2012–2013 | Malawi | 0 | 22.1 | 0.0 (0.0, 16.7)* |
| Wirtz, 2015 | 2013–2013 | Malawi | 0 | 23.1 | 0.0 (0.0, 16.7)* |
| **Southern Africa** | | | | | |
| Buchbinder, 2014 | 2007–2010 | South Africa | 2 | 43 | 4.7 (1.2, 18.6) |
| Kamali, 2015 | 2009–2010 | South Africa | 2 | 21 | 9.5 (2.4, 38.1) |
| Lane, 2016 | 2012–2015 | South Africa | 18 | 144.2 | 12.5 (7.9, 19.8) |
| Maenetje, 2019 | 2012–2015 | South Africa | 0 | 20.2 | 0.0 (0.0, 18.3) |
| Sandfort, 2021 | 2015–2017 | South Africa | 10 | 69.3 | 14.4 (7.8, 26.8) |
| Sullivan, 2020 | 2015–2016 | South Africa | 7 | 133.3 | 5.3 (2.5, 11.0) |
| **Central Africa** | | | | | |
| Mbeko Simaleko, 2018 | 2010–2012 | CAR | 12 | 128 | 9.4 (5.3, 16.5)* |
| Mbeko Simaleko, 2020 | 2014–2016 | CAR | 10 | 200.2 | 5.0 (2.7, 9.3)* |
| **Western Africa** | | | | | |
| Couderc, 2017 | 2013–2013 | Mali | 1 | 89.2 | 1.1 (0.2, 8.0) |
| Couderc, 2017 | 2013–2013 | Senegal | 0 | 27 | 0.0 (0.0, 13.7) |
| Couderc, 2017 | 2013–2015 | Cote d'Ivoire | 7 | 44 | 15.9 (7.6, 33.4) |
| Dah, 2021 | 2015–2018 | Cote d'Ivoire | 23 | 159.8 | 14.4 (9.6, 21.7) |
| Dah, 2021 | 2015–2018 | Mali | 35 | 389.7 | 9.0 (6.4, 12.5) |
| Dah, 2021 | 2016–2017 | Togo | 11 | 107.6 | 10.2 (5.7, 18.5) |
| Dah, 2021 | 2016–2017 | Burkina Faso | 9 | 123.7 | 7.3 (3.8, 14.0) |
| Drame, 2013 | 2011–2012 | Senegal | 8 | 50 | 16.0 (8.0, 32.0) |
| Hessou, 2020 | 2016–2016 | Benin | 20 | 172.5 | 11.6 (7.5, 18.0) |
| Hessou, 2020 | 2016–2017 | Benin | 11 | 162.8 | 6.8 (3.7, 12.2) |
| Hessou, 2020 | 2017–2017 | Benin | 3 | 157 | 1.9 (0.6, 5.9) |
| Hessou, 2020 | 2017–2018 | Benin | 0 | 154.5 | 0.0 (0.0, 2.4) |
| Hessou, 2020 | 2018–2018 | Benin | 14 | 151 | 9.3 (5.5, 15.7) |
| Lyons, 2020 | 2015–2017 | Senegal | 8 | 249.5 | 3.2 (1.6, 6.4) |
| Ramadhani, 2020 | 2013–2019 | Nigeria | 65 | 631.2 | 10.3 (8.1, 13.1)* |

*HIV Incidence Rate*

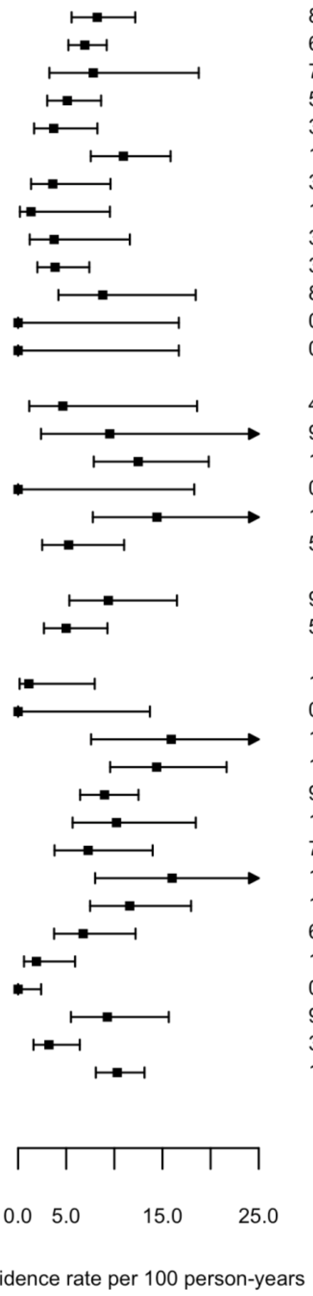


***** observation calculated using available data reported within article

**Figure S15. Forest plot of study observations of HIV incidence among men who have sex with men (MSM), by region of Africa.**

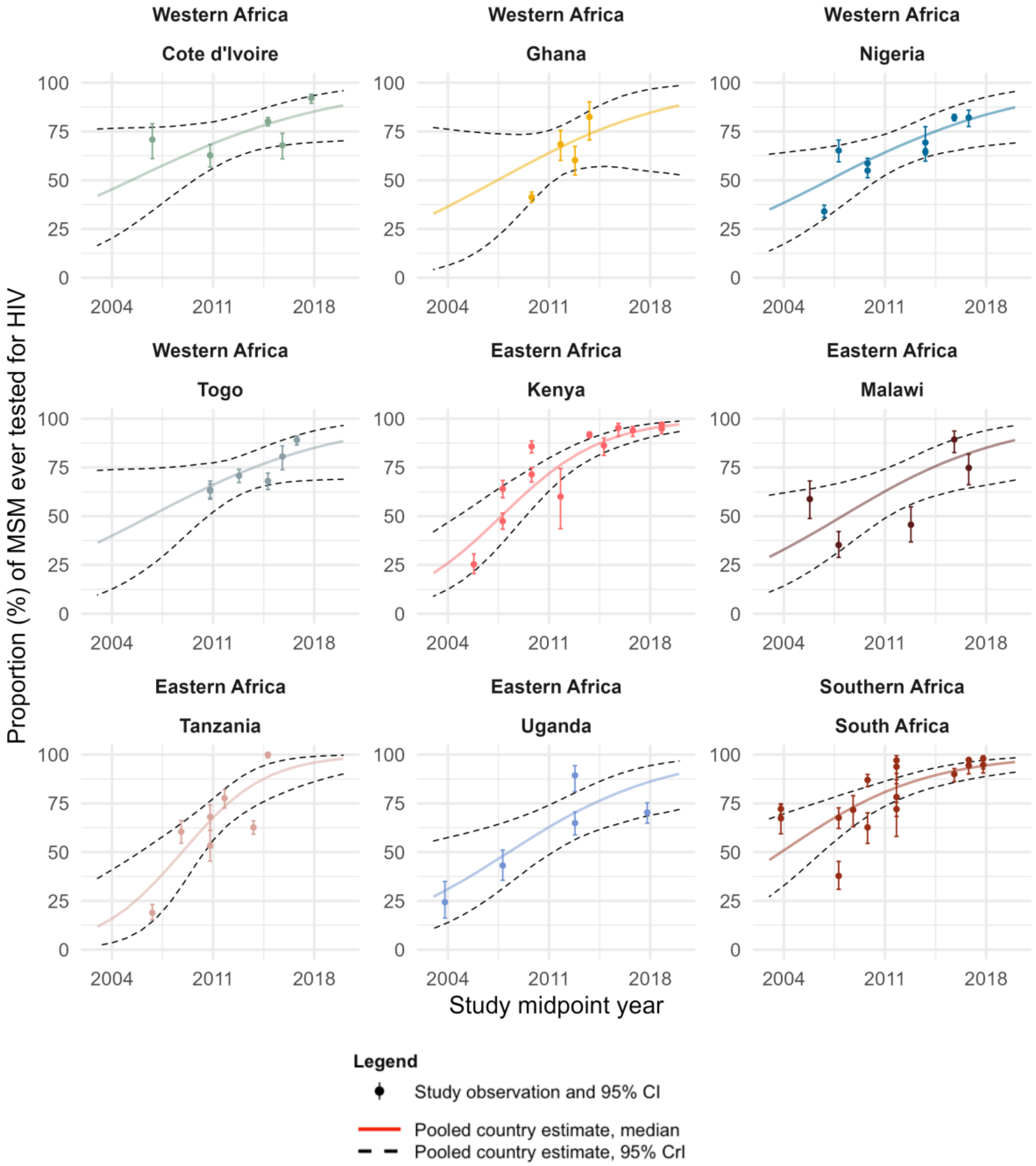


**Figure S16. Ever HIV testing among men who have sex with men (MSM) over time, by country of Africa.** Estimates are shown over the range of available years in each region of Africa for countries with at least 3 observations from different time points. Points represent available study observations and their 95% confidence intervals. The solid and dotted lines represent the estimated country-level proportions and 95% credible intervals (CrI) over time, respectively.

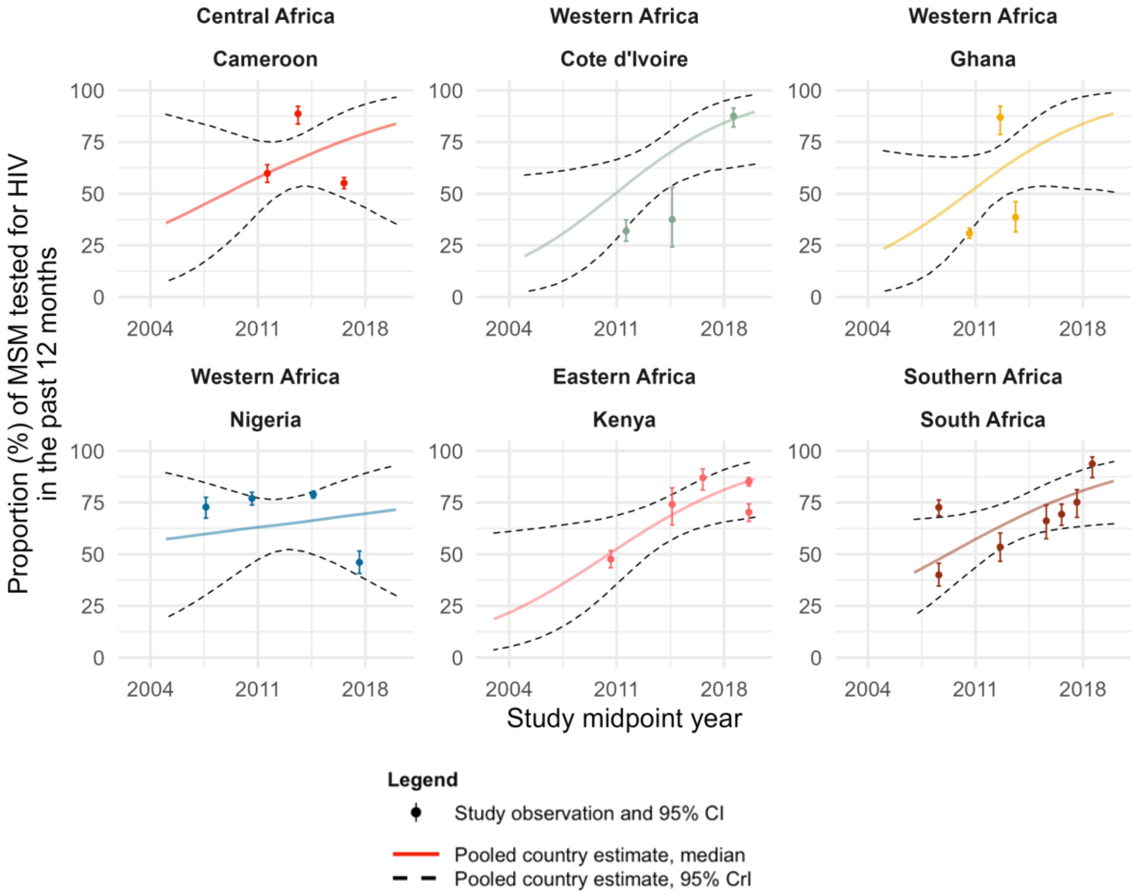


**Figure S17. HIV testing in the past 12 months among men who have sex with men (MSM) over time, by country of Africa.** Estimates are shown over the range of available years in each region of Africa for countries with at least 3 observations from different time points. Points represent available study observations and their 95% confidence intervals. The solid and dotted lines represent the estimated country-level proportions and 95% credible intervals (CrI) over time, respectively.

**Table S2. Estimated time trends in HIV testing in the past 6 months among men who have sex with men (MSM) in Africa and estimated outcomes in 2010 and 2020, overall and by region of Africa.**

| Outcome | Region of Africa | No | Estimate of time trend | 95% CrI | Estimate in 2010 | 95% CrI | Estimate in 2020 | 95% CrI |
|---|---|---|---|---|---|---|---|---|
| **Past 6 months HIV testing (%)** | | | | | | | | |
| Among all MSM* | Overall | 23 | OR=0.85 | 0.40-1.74 | 68% | 7-99% | 31% | 1-96% |
| | Central/Western Africa | 7 | OR=0.64 | 0.41-1.08 | 88% | 37-99% | 7% | 1-60% |
| | Eastern/Southern Africa | 16 | OR=1.08 | 0.76-1.36 | 43% | 18-86% | 65% | 25-86% |

ART, antiretroviral therapy; CrI, credible interval; IRR, incidence rate ratio; MSM, men who have sex with men; OR, odds ratio.

* n = 1 observation from Central/Eastern Africa not shown

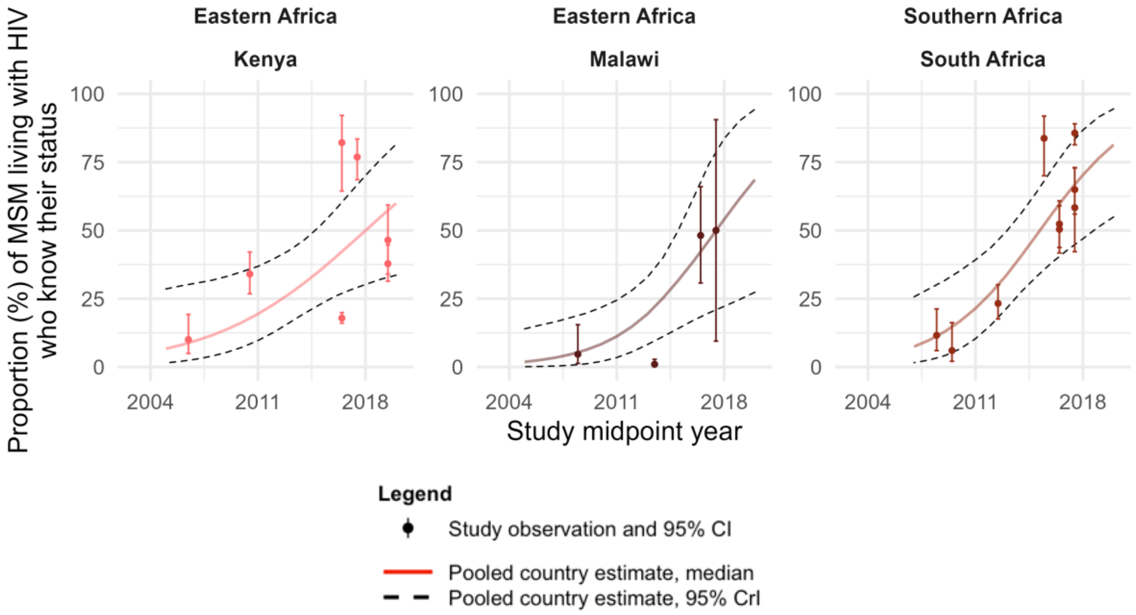


**Figure S18. Knowledge of status (self-reported) among men who have sex with men (MSM) living with HIV over time, by country of Africa.** Estimates are shown over the range of available years in each region of Africa for countries with at least 3 observations from different time points. Points represent available study observations and their 95% confidence intervals. The solid and dotted lines represent the estimated country-level proportions and 95% credible intervals (CrI) over time, respectively.

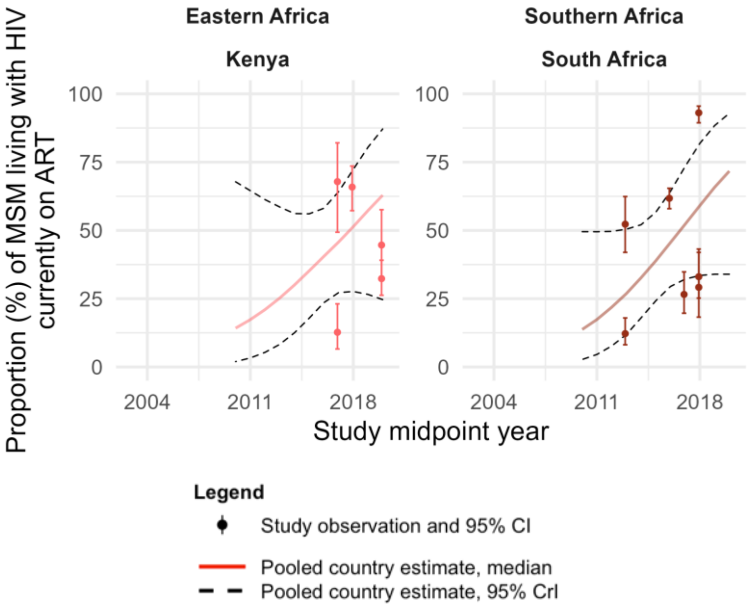


**Figure S19. Current antiretroviral therapy (ART) use among men who have sex with men (MSM) living with HIV over time, by country of Africa.** Estimates are shown over the range of available years in each region of Africa for countries with at least 3 observations from different time points. Points represent available study observations and their 95% confidence intervals. The solid and dotted lines represent the estimated country-level proportions and 95% credible intervals (CrI) over time, respectively.

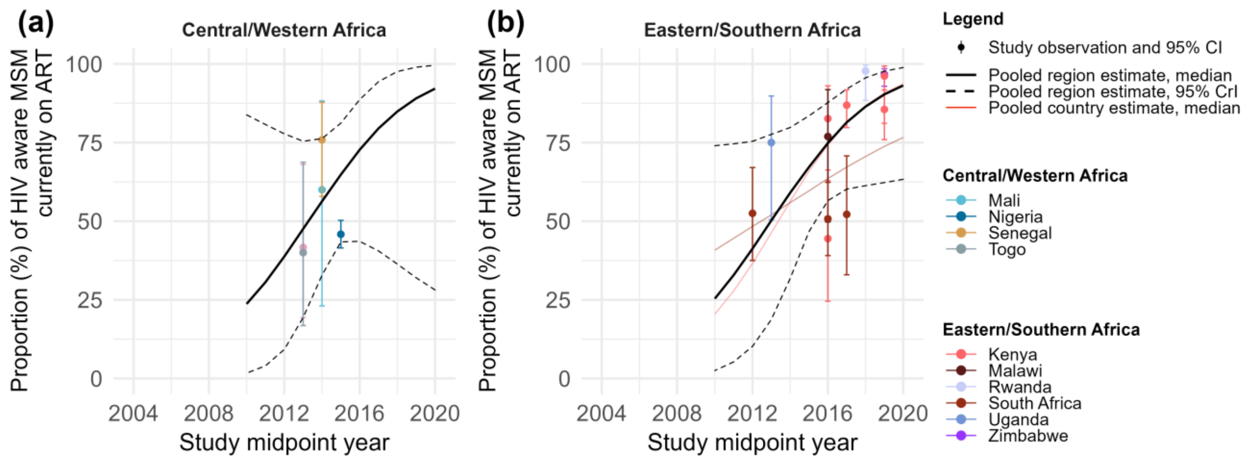


**Figure S20. Current antiretroviral therapy (ART) use among HIV aware men who have sex with men (MSM) over time, by region and country of Africa.** Current ART use among HIV aware MSM in (a) Central/Western Africa, and (b) Eastern/Southern Africa. Points represent available study observations and their 95% confidence intervals, coloured by country in which the study was conducted. The black solid and dotted lines represent the estimated region-level proportions and 95% credible intervals (CrI), respectively. Coloured solid lines represent estimated country-level proportions for countries with at least 3 estimates from 3 different time points (see Figure S20 for individual country-level time trends and 95% CrI).

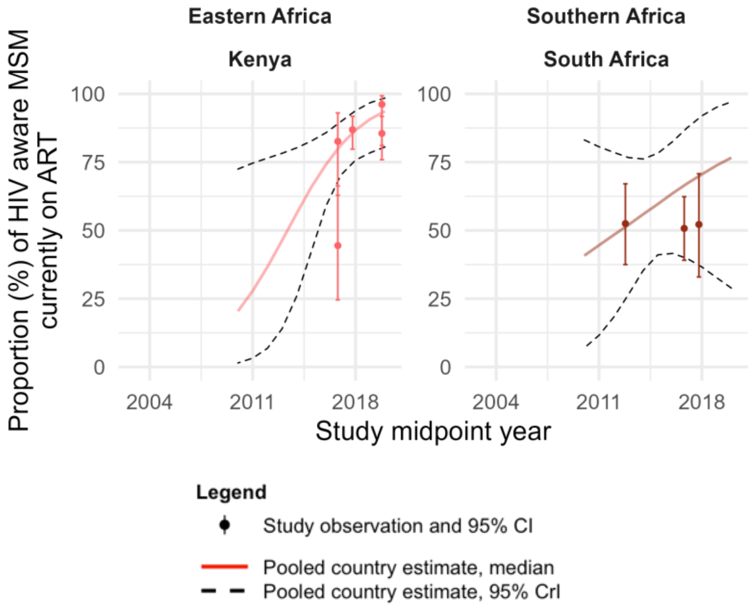


**Figure S21. Current antiretroviral therapy (ART) use among HIV aware men who have sex with men (MSM) over time, by country of Africa.** Estimates are shown over the range of available years in each region of Africa for countries with at least 3 observations from different time points. Points represent available study observations and their 95% confidence intervals. The solid and dotted lines represent the estimated country-level proportions and 95% credible intervals (CrI) over time, respectively.

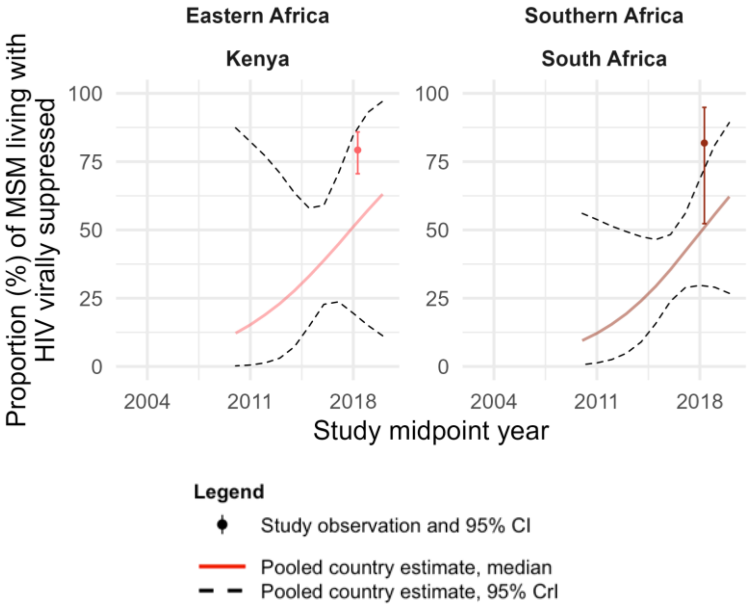


**Figure S22. Viral suppression among men who have sex with men (MSM) living with HIV over time, by country of Africa.** Estimates are shown over the range of available years in each region of Africa for countries with at least 3 observations from different time points. Points represent available study observations and their 95% confidence intervals. The solid and dotted lines represent the estimated country-level proportions and 95% credible intervals (CrI) over time, respectively.

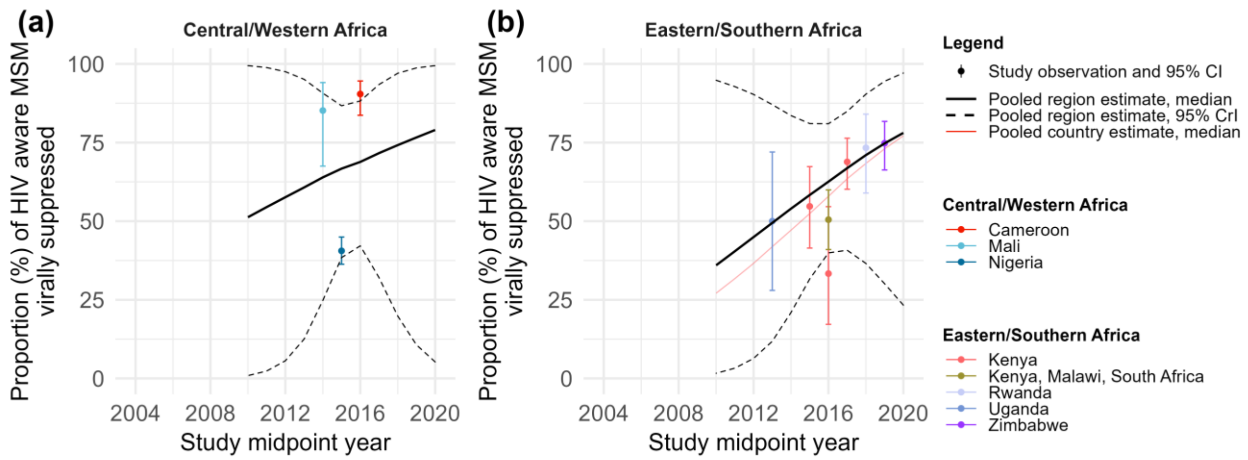


**Figure S23. Viral suppression among HIV aware men who have sex with men (MSM) over time, by region and country of Africa.** Viral suppression among HIV aware MSM in (a) Central/Western Africa, and (b) Eastern/Southern Africa. Points represent available study observations and their 95% confidence intervals, coloured by country in which the study was conducted. The black solid and dotted lines represent the estimated region-level proportions and 95% credible intervals (CrI), respectively. Coloured solid lines represent estimated country-level proportions for countries with at least 3 estimates from 3 different time points (see Figure S22 for individual country-level time trends and 95% CrI).

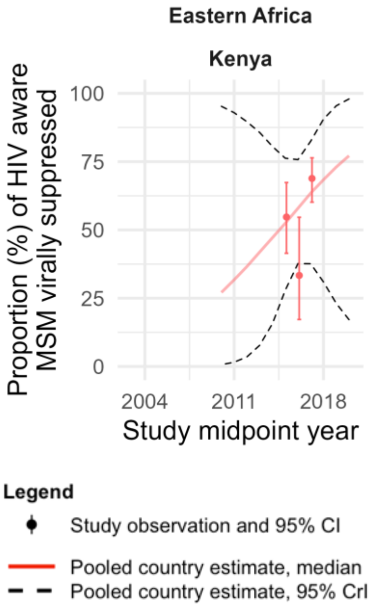


**Figure S24. Viral suppression among HIV aware men who have sex with men (MSM) over time, by country of Africa.** Estimates are shown over the range of available years in each region of Africa for countries with at least 3 observations from different time points. Points represent available study observations and their 95% confidence intervals. The solid and dotted lines represent the estimated country-level proportions and 95% credible intervals (CrI) over time, respectively.

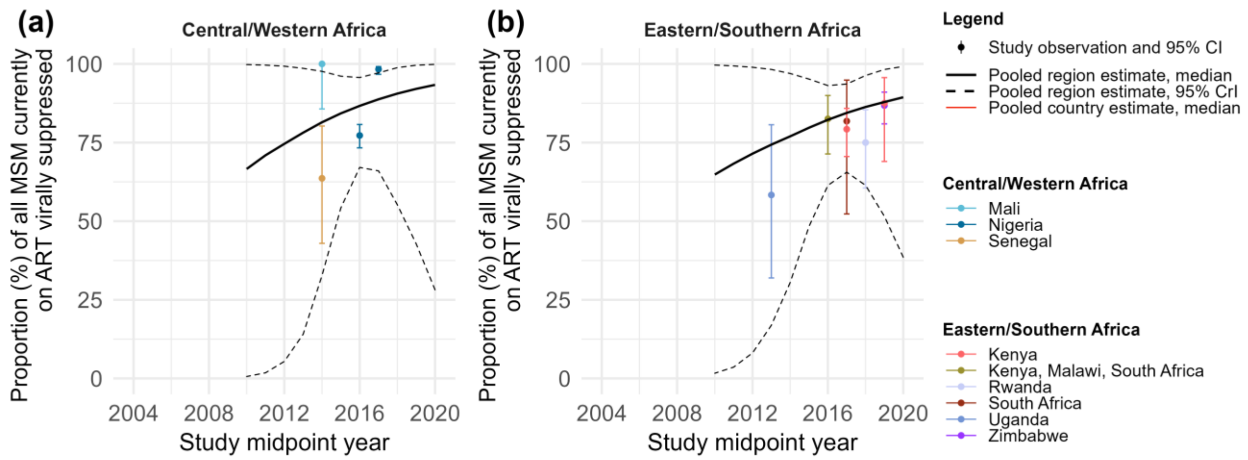


**Figure S25. Viral suppression among men who have sex with men (MSM) currently on antiretroviral therapy (ART) over time, by region and country of Africa.** Viral suppression among MSM currently on ART over time in (a) Central/Western Africa and (b) Eastern/Southern Africa. Points represent available study observations and their 95% confidence intervals, coloured by country in which the study was conducted. The black solid and dotted lines represent the estimated region-level proportions and 95% credible intervals (CrI), respectively. Coloured solid lines represent estimated country-level proportions for countries with at least 3 estimates from 3 different time points (see Figure S22 for individual country-level time trends and 95% CrI).

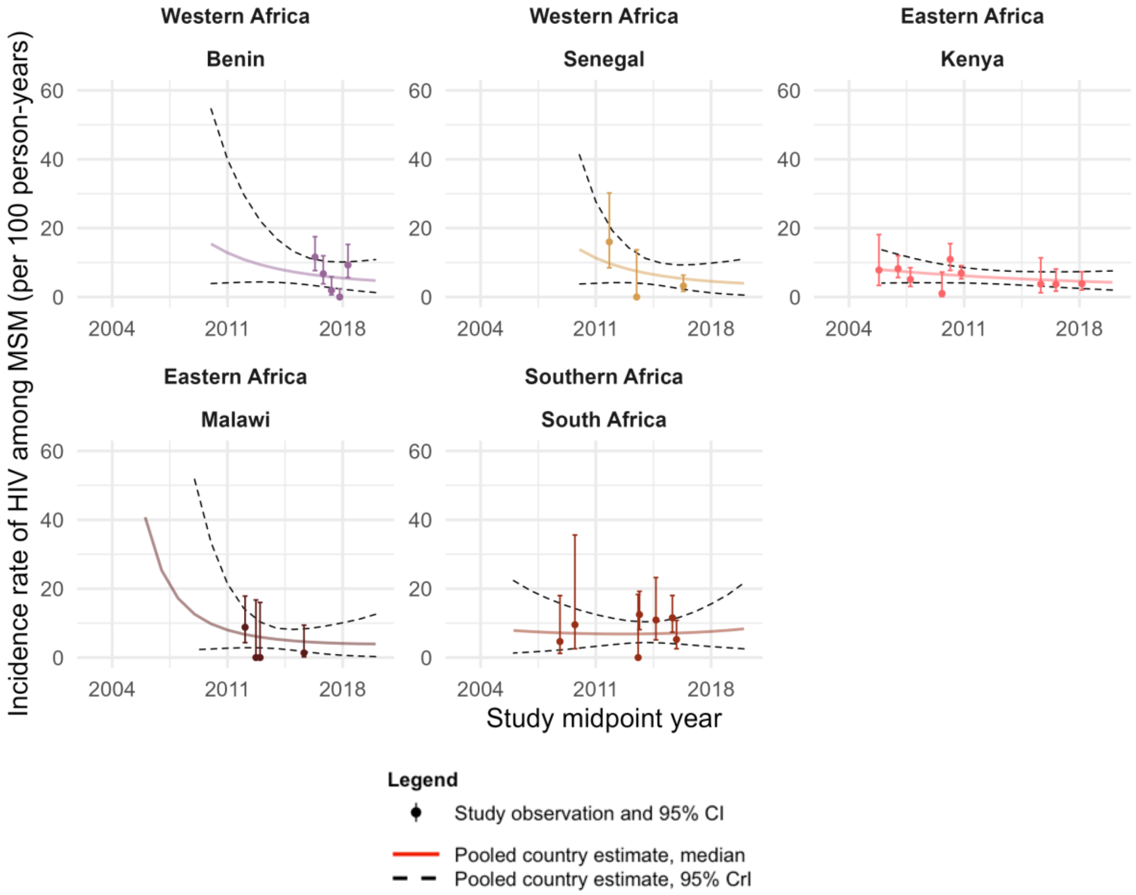


**Figure S26. HIV incidence among men who have sex with men (MSM) over time, by country of Africa.** Estimates are shown over the range of available years in each region of Africa for countries with at least 3 observations from different time points. Points represent available study observations and their 95% confidence intervals. The solid and dotted lines represent the estimated country-level proportions and 95% credible intervals (CrI) over time, respectively.

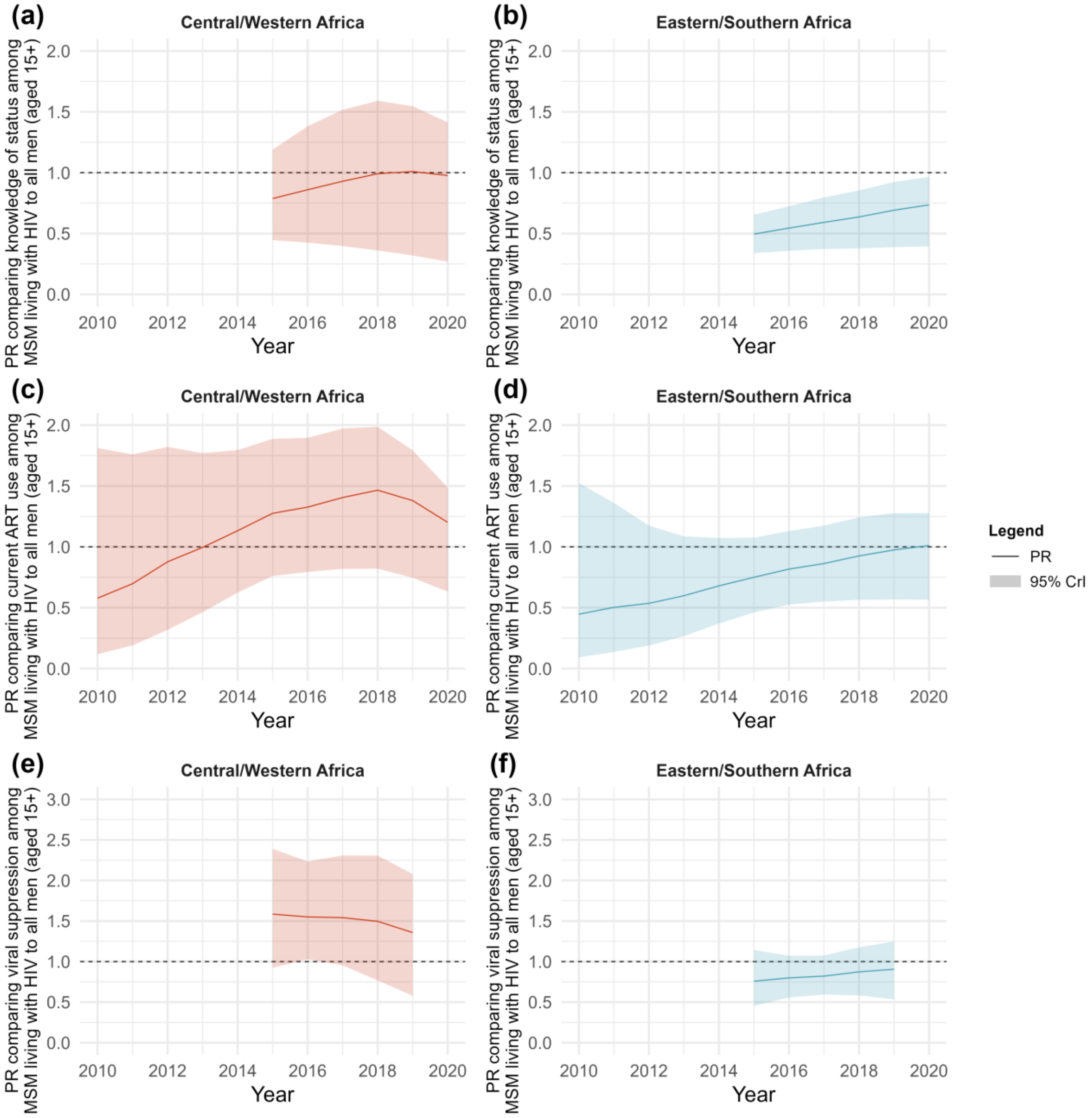


**Figure S27. Prevalence ratios (PR) and 95% credible intervals (CrI) comparing our estimates of HIV treatment cascade outcomes among men who have sex with men (MSM) living with HIV with UNAIDS estimates among all men living with HIV (aged 15+).** PRs comparing estimates of (a) knowledge of status in Central/Western Africa, and (b) knowledge of status in Eastern/Southern Africa, (c) current antiretroviral therapy (ART) use in Central/Western Africa, and (d) current ART use in Eastern/Southern Africa, and (e) viral suppression in Central/Western Africa, and (f) viral suppression in Eastern/Southern Africa among MSM living with HIV with UNAIDS estimates among all men living with HIV (aged 15+). PRs were estimated over the range of years of estimates available for all men.

**Table S3. Study quality assessment of studies included in our review.** Studies received a score ranging from 0-5 for each outcome reported in the study.

| References | Country | Midpoint Year | Outcomes reported | Criterion 1: Appropriateness of the sampling method to recruit a representative sample of MSM participants (maximum 1 point) | Criterion 2: Statistical adjustment of outcomes for complex sampling design (maximum 1 point) | Criterion 3: Generalizability of the results based on the study definition of MSM (maximum 1 point) | Criterion 4: Inclusion of transgender women in the study definition of MSM (maximum 1 point) | Criterion 5: Risk of misclassification in ascertainment of the relevant outcome(s) (maximum 1 point) | Study quality score /5 |
|---|---|---|---|---|---|---|---|---|---|
| **Northern Africa** | | | | | | | | | |
| **Valadez 2013[1]** | Libya | 2010 | HIV testing in the past 12 months | a = 1 point | a = 1 point | a = 1 point | b = 0 points | c = 0 points | 3 |
| **Elmahy 2018[2]** | Egypt | 2016 | HIV testing ever | b = 0 points | b = 0 points | b = 0 points | b = 0 points | b = 1 point | 1 |
| **Central Africa** | | | | | | | | | |
| **Kendall 2014[3]** | Angola | 2011 | HIV testing ever | a = 1 point | b = 0 points | a = 1 point | b = 0 points | c = 0 points | 2 |
| | | | HIV testing in the past 12 months | a = 1 point | b = 0 points | a = 1 point | b = 0 points | c = 0 points | 2 |
| | | | HIV testing in the past 3 months | a = 1 point | b = 0 points | a = 1 point | b = 0 points | c = 0 points | 2 |
| | | | Knowledge of status | a = 1 point | b = 0 points | a = 1 point | b = 0 points | c = 0 points | 2 |
| **Herce 2018[4]** | Angola | 2017 | HIV testing ever | a = 1 point | a = 1 point | a = 1 point | a = 1 point | c = 0 points | 4 |
| | | | HIV testing in the past 6 months | a = 1 point | a = 1 point | a = 1 point | a = 1 point | c = 0 points | 4 |
| | | | Knowledge of status | a = 1 point | a = 1 point | a = 1 point | a = 1 point | c = 0 points | 4 |
| **Lorente, 2012[5]** | Cameroon | 2008 | HIV testing ever | a = 1 point | b = 0 points | a = 1 point | b = 0 points | c = 0 points | 2 |
| **Holland, 2015[6]; Park, 2014[7]** | Cameroon | 2011 | HIV testing ever | a = 1 point | a = 1 point | a = 1 point | b = 0 points | c = 0 points | 3 |
| | | | HIV testing in the past 12 months | a = 1 point | a = 1 point | a = 1 point | b = 0 points | c = 0 points | 3 |
| **Rao 2017[8]** | Cameroon | 2013 | HIV testing in the past 12 months | a = 1 point | b = 0 points | a = 1 point | b = 0 points | c = 0 points | 2 |
| **Bowring 2019[9], Rao 2017[8]** | Cameroon | 2015/ 2016 | HIV testing in the past 12 months | a = 1 point | a = 1 point | a = 1 point | b = 0 points | c = 0 points | 3 |
| | | | Knowledge of status | a = 1 point | b = 0 points | a = 1 point | b = 0 points | c = 0 points | 2 |

| References | Country | Midpoint Year | Outcomes reported | Criterion 1: Appropriateness of the sampling method to recruit a representative sample of MSM participants (maximum 1 point) | Criterion 2: Statistical adjustment of outcomes for complex sampling design (maximum 1 point) | Criterion 3: Generalizability of the results based on the study definition of MSM (maximum 1 point) | Criterion 4: Inclusion of transgender women in the study definition of MSM (maximum 1 point) | Criterion 5: Risk of misclassification in ascertainment of the relevant outcome(s) (maximum 1 point) | Study quality score /5 |
|---|---|---|---|---|---|---|---|---|---|
| | | | Ever ART use | a = 1 point | b = 0 points | a = 1 point | b = 0 points | c = 0 points | 2 |
| | | | Viral suppression | a = 1 point | b = 0 points | a = 1 point | b = 0 points | a = 1 point | 3 |
| **Bouassa 2018[10], Gresenguet 2017[11], Longo 2018[12]** | Central African Republic | 2010 | HIV testing ever | b = 0 points | b = 0 points | b = 0 points | b = 0 points | c = 0 points | 0 |
| | | | Current antiretroviral therapy (ART) use | b = 0 points | b = 0 points | b = 0 points | b = 0 points | c = 0 points | 0 |
| **Mbeko Simaleko 2018[13]** | Central African Republic | 2011 | HIV incidence | b = 0 points | b = 0 points | b = 0 points | c = 0 points | a = 1 point | 1 |
| **Mbeko Simaleko 2020[14]** | Central African Republic | 2015 | HIV incidence | c = 0 points | b = 0 points | c = 0 points | c = 0 points | a = 1 point | 1 |
| **Western Africa** | | | | | | | | | |
| **Hessou 2020[15]** | Benin | 2017 | HIV incidence | a = 1 point | b = 0 points | a = 1 point | b = 0 points | a = 1 point | 3 |
| **Ahouada 2020[16]** | Benin | 2018 | HIV testing in the past 12 months | a = 1 point | b = 0 points | a = 1 point | a = 1 point | c = 0 points | 3 |
| **Goodman, 2016[17], Grosso 2019[18], Holland 2016[19], Kim 2018[20], Poteat 2017[21], Stahlman 2016[22]** | Burkina Faso | 2013 | HIV testing ever | a = 1 point | b = 0 points | a = 1 point | b = 0 points | c = 0 points | 2 |
| | | | Knowledge of status | a = 1 point | b = 0 points | a = 1 point | b = 0 points | c = 0 points | 2 |
| | | | Current ART use | a = 1 point | b = 0 points | a = 1 point | b = 0 points | c = 0 points | 2 |
| **Coulaud 2020[23], Dah 2021[24], Dah 2021[25], Laurent 2021[26], Yaya** | Burkina Faso | 2017 | HIV testing ever | b = 0 points | b = 0 points | b = 0 points | b = 0 points | c = 0 points | 0 |
| | | | Engagement in care | b = 0 points | b = 0 points | b = 0 points | b = 0 points | c = 0 points | 0 |
| | | | HIV incidence | b = 0 points | b = 0 points | b = 0 points | b = 0 points | a = 1 point | 1 |

| References | Country | Midpoint Year | Outcomes reported | Criterion 1: Appropriateness of the sampling method to recruit a representative sample of MSM participants (maximum 1 point) | Criterion 2: Statistical adjustment of outcomes for complex sampling design (maximum 1 point) | Criterion 3: Generalizability of the results based on the study definition of MSM (maximum 1 point) | Criterion 4: Inclusion of transgender women in the study definition of MSM (maximum 1 point) | Criterion 5: Risk of misclassification in ascertainment of the relevant outcome(s) (maximum 1 point) | Study quality score /5 |
|---|---|---|---|---|---|---|---|---|---|
| **2021[27], Yaya 2022[28]** | | | | | | | | | |
| **Vuylsteke 2012[29]** | Côte d'Ivoire | 2007 | HIV testing ever | b = 0 points | b = 0 points | b = 0 points | c = 0 points | c = 0 points | 0 |
| **Aho 2014[30], Hakim 2015[31]** | Côte d'Ivoire | 2011 | HIV testing ever | a = 1 point | a = 1 point | a = 1 point | b = 0 points | c = 0 points | 3 |
| | | | HIV testing in the past 12 months | a = 1 point | a = 1 point | a = 1 point | b = 0 points | c = 0 points | 3 |
| | | | Knowledge of status | a = 1 point | a = 1 point | a = 1 point | b = 0 points | c = 0 points | 3 |
| **Couderc 2017[32]** | Côte d'Ivoire | 2014 | HIV incidence | b = 0 points | b = 0 points | b = 0 points | c = 0 points | a = 1 point | 1 |
| **Bouscaillou 2016[33]** | Côte d'Ivoire | 2014 | HIV testing in the past 12 months | a = 1 point | b = 0 points | b = 0 points | b = 0 points | c = 0 points | 1 |
| **Moran 2019[34], Ulanja 2019[35]** | Côte d'Ivoire | 2015 | HIV testing ever | a = 1 point | a = 1 point | a = 1 point | b = 0 points | c = 0 points | 3 |
| | | | HIV testing in the past 6 months | a = 1 point | a = 1 point | a = 1 point | b = 0 points | c = 0 points | 3 |
| **Coulaud 2020[23], Dah 2021[24], Dah 2021[25], Laurent 2021[26], Yaya 2021[27], Yaya 2022[28]** | Côte d'Ivoire | 2016 | HIV testing ever | b = 0 points | b = 0 points | b = 0 points | b = 0 points | c = 0 points | 0 |
| | | | Engagement in care | b = 0 points | b = 0 points | b = 0 points | b = 0 points | c = 0 points | 0 |
| | | | Current ART use | b = 0 points | b = 0 points | b = 0 points | b = 0 points | c = 0 points | 0 |
| | | | HIV incidence | b = 0 points | b = 0 points | b = 0 points | b = 0 points | a = 1 point | 1 |
| **Diabate 2020[36]** | Côte d'Ivoire | 2018 | HIV testing in the past 12 months | a = 1 point | b = 0 points | a = 1 point | c = 0 points | c = 0 points | 2 |
| **Inghels 2021[37]** | Côte d'Ivoire | 2018 | HIV testing ever | a = 1 point | a = 1 point | a = 1 point | c = 0 points | c = 0 points | 3 |
| **Gyamerah 2020[38]** | Ghana | 2010 | HIV testing ever | a = 1 point | b = 0 points | a = 1 point | b = 0 points | c = 0 points | 2 |
| | | | HIV testing in the past 12 months | a = 1 point | b = 0 points | a = 1 point | b = 0 points | c = 0 points | 2 |
| | Ghana | 2012 | HIV testing ever | a = 1 point | b = 0 points | a = 1 point | b = 0 points | b = 1 point | 3 |

| References | Country | Midpoint Year | Outcomes reported | Criterion 1: Appropriateness of the sampling method to recruit a representative sample of MSM participants (maximum 1 point) | Criterion 2: Statistical adjustment of outcomes for complex sampling design (maximum 1 point) | Criterion 3: Generalizability of the results based on the study definition of MSM (maximum 1 point) | Criterion 4: Inclusion of transgender women in the study definition of MSM (maximum 1 point) | Criterion 5: Risk of misclassification in ascertainment of the relevant outcome(s) (maximum 1 point) | Study quality score /5 |
|---|---|---|---|---|---|---|---|---|---|
| **Kushwaha 2017[39], Nelson 2015[40]** | | | HIV testing in the past 12 months | a = 1 point | b = 0 points | a = 1 point | b = 0 points | b = 1 point | 3 |
| | | | HIV testing in the past 6 months | a = 1 point | b = 0 points | a = 1 point | b = 0 points | b = 1 point | 3 |
| | | | HIV testing in the past 3 months | a = 1 point | b = 0 points | a = 1 point | b = 0 points | b = 1 point | 3 |
| **Girault 2015[41]** | Ghana | 2013 | HIV testing ever | a = 1 point | b = 0 points | a = 1 point | b = 0 points | c = 0 points | 2 |
| | | | HIV testing in the past 12 months | a = 1 point | a = 1 point | a = 1 point | b = 0 points | c = 0 points | 3 |
| **Abubakari 2021[42]** | Ghana | 2014 | HIV testing ever | a = 1 point | b = 0 points | b = 0 points | a = 1 point | c = 0 points | 2 |
| | | | HIV testing in the past 12 months | a = 1 point | b = 0 points | b = 0 points | a = 1 point | c = 0 points | 2 |
| | | | HIV testing in the past 6 months | a = 1 point | b = 0 points | b = 0 points | a = 1 point | c = 0 points | 2 |
| **Ogunbajo 2017[43]** | Ghana | 2015 | Engagement in care | b = 0 points | b = 0 points | a = 1 point | b = 0 points | c = 0 points | 1 |
| **Gu 2021[44]** | Ghana | 2017 | Engagement in care | b = 0 points | b = 0 points | a = 1 point | a = 1 point | b = 1 point | 3 |
| **Lieber 2018[45]** | Liberia | NR | HIV testing ever | b = 0 points | b = 0 points | a = 1 point | b = 0 points | c = 0 points | 1 |
| **Couderc 2017[32]** | Mali | 2013 | HIV incidence | b = 0 points | b = 0 points | b = 0 points | c = 0 points | a = 1 point | 1 |
| **Hakim 2017[46], Hakim 2018[47], Lahuerta 2018[48], Knox 2021[49]** | Mali | 2014 | HIV testing ever | a = 1 point | a = 1 point | a = 1 point | b = 0 points | c = 0 points | 3 |
| | | | HIV testing in the past 12 months | a = 1 point | a = 1 point | a = 1 point | b = 0 points | c = 0 points | 3 |
| | | | Knowledge of status | a = 1 point | a = 1 point | a = 1 point | b = 0 points | c = 0 points | 3 |
| | | | Current ART use | a = 1 point | a = 1 point | a = 1 point | b = 0 points | c = 0 points | 3 |
| | | | Viral suppression | a = 1 point | a = 1 point | a = 1 point | b = 0 points | a = 1 point | 4 |
| | Mali | 2016 | HIV testing ever | b = 0 points | b = 0 points | b = 0 points | b = 0 points | c = 0 points | 0 |

| References | Country | Midpoint Year | Outcomes reported | Criterion 1: Appropriateness of the sampling method to recruit a representative sample of MSM participants (maximum 1 point) | Criterion 2: Statistical adjustment of outcomes for complex sampling design (maximum 1 point) | Criterion 3: Generalizability of the results based on the study definition of MSM (maximum 1 point) | Criterion 4: Inclusion of transgender women in the study definition of MSM (maximum 1 point) | Criterion 5: Risk of misclassification in ascertainment of the relevant outcome(s) (maximum 1 point) | Study quality score /5 |
|---|---|---|---|---|---|---|---|---|---|
| **Coulaud 2020[23], Dah 2021[24], Dah 2021[25], Laurent 2021[26], Yaya 2021[27], Yaya 2022[28]** | | | Engagement in care | b = 0 points | b = 0 points | b = 0 points | b = 0 points | c = 0 points | 0 |
| | | | Current ART use | b = 0 points | b = 0 points | b = 0 points | b = 0 points | c = 0 points | 0 |
| | | | HIV incidence | b = 0 points | b = 0 points | b = 0 points | b = 0 points | a = 1 point | 1 |
| **Koyalta 2021[50]** | Mali | 2019 | Current ART use | b = 0 points | b = 0 points | b = 0 points | c = 0 points | c = 0 points | 0 |
| **Adam 2009[51]** | Mauritania | 2006 | HIV testing in the past 12 months | c = 0 points | b = 0 points | c = 0 points | c = 0 points | d = 0 points | 0 |
| **Adam 2009[51], Merrigan 2011[52]** | Nigeria | 2007 | HIV testing ever | a = 1 point | a = 1 point | a = 1 point | b = 0 points | c = 0 points | 3 |
| | | | HIV testing in the past 12 months | a = 1 point | a = 1 point | a = 1 point | b = 0 points | c = 0 points | 3 |
| **Stromdahl 2012[53], Stromdahl 2019[54]** | Nigeria | 2008 | HIV testing ever | b = 0 points | b = 0 points | a = 1 point | b = 0 points | c = 0 points | 1 |
| **Eluwa 2015[55], Eluwa 2019[56]** | Nigeria | 2010 | HIV testing ever | a = 1 point | a = 1 point | a = 1 point | b = 0 points | c = 0 points | 3 |
| | | | HIV testing in the past 12 months | a = 1 point | a = 1 point | a = 1 point | b = 0 points | c = 0 points | 3 |
| **Adebajo 2014[57], Sheehy 2014[58], Vu 2013[59], Vu 2013[60]** | Nigeria | 2010 | HIV testing ever | a = 1 point | a = 1 point | a = 1 point | b = 0 points | c = 0 points | 3 |

| References | Country | Midpoint Year | Outcomes reported | Criterion 1: Appropriateness of the sampling method to recruit a representative sample of MSM participants (maximum 1 point) | Criterion 2: Statistical adjustment of outcomes for complex sampling design (maximum 1 point) | Criterion 3: Generalizability of the results based on the study definition of MSM (maximum 1 point) | Criterion 4: Inclusion of transgender women in the study definition of MSM (maximum 1 point) | Criterion 5: Risk of misclassification in ascertainment of the relevant outcome(s) (maximum 1 point) | Study quality score /5 |
|---|---|---|---|---|---|---|---|---|---|
| **Baral 2015[61], Billings 2019[62], Charurat 2015[63], Crowell 2017[64], Crowell 2019[65], Kayode 2020[66], Li 2020[67], Nowak 2016[68], Nowak 2017[69], Nowak 2019[70], Nowak 2019[71], Nowak 2020[72], Olawore 2021[73], Ramadhani 2017[74], Ramadhani 2018[75], Ramadhani 2020[76], Robbins 2020[77], Rodriguez-Hart 2016[78], Rodriguez-Hart 2018[79], Schwartz 2015[80], Stahlman 2017[81], Tiamiyu 2020[82]** | Nigeria | 2013 | HIV testing ever | a = 1 point | b = 0 points | a = 1 point | b = 0 points | c = 0 points | 2 |
| | | | Knowledge of status | a = 1 point | b = 0 points | a = 1 point | b = 0 points | c = 0 points | 2 |
| | | | Engagement in care | a = 1 point | b = 0 points | a = 1 point | b = 0 points | c = 0 points | 2 |
| | | | Ever ART use | a = 1 point | b = 0 points | a = 1 point | b = 0 points | c = 0 points | 2 |
| | | | Current ART use | a = 1 point | b = 0 points | a = 1 point | b = 0 points | c = 0 points | 2 |
| | | | Viral suppression | a = 1 point | b = 0 points | a = 1 point | b = 0 points | a = 1 point | 3 |
| | | | HIV incidence | a = 1 point | b = 0 points | a = 1 point | a = 1 point | a = 1 point | 4 |
| | Nigeria | 2014 | HIV testing ever | b = 0 points | b = 0 points | a = 1 point | b = 0 points | a = 1 point | 2 |

| References | Country | Midpoint Year | Outcomes reported | Criterion 1: Appropriateness of the sampling method to recruit a representative sample of MSM participants (maximum 1 point) | Criterion 2: Statistical adjustment of outcomes for complex sampling design (maximum 1 point) | Criterion 3: Generalizability of the results based on the study definition of MSM (maximum 1 point) | Criterion 4: Inclusion of transgender women in the study definition of MSM (maximum 1 point) | Criterion 5: Risk of misclassification in ascertainment of the relevant outcome(s) (maximum 1 point) | Study quality score /5 |
|---|---|---|---|---|---|---|---|---|---|
| **Tobin-West 2017[83]** | | | HIV testing in the past 6 months | b = 0 points | b = 0 points | a = 1 point | b = 0 points | a = 1 point | 2 |
| **Eluwa 2019[56]** | Nigeria | 2014 | HIV testing ever | a = 1 point | a = 1 point | a = 1 point | c = 0 points | c = 0 points | 3 |
| | | | HIV testing in the past 12 months | a = 1 point | a = 1 point | a = 1 point | c = 0 points | c = 0 points | 3 |
| **Offie 2021[84]** | Nigeria | 2016 | Engagement in care | b = 0 points | b = 0 points | b = 0 points | c = 0 points | c = 0 points | 0 |
| **Tun 2018[85]** | Nigeria | 2017 | HIV testing ever | a = 1 point | b = 0 points | a = 1 point | b = 0 points | c = 0 points | 2 |
| | | | HIV testing in the past 12 months | a = 1 point | b = 0 points | a = 1 point | b = 0 points | c = 0 points | 2 |
| **Ibiloye 2018[86]** | Nigeria | 2017 | HIV testing ever | b = 0 points | b = 0 points | b = 0 points | c = 0 points | d = 0 points | 0 |
| **Ibiloye 2021[87]** | Nigeria | 2017 | Engagement in care | b = 0 points | b = 0 points | c = 0 points | a = 1 point | d = 0 points | 1 |
| | | | Viral suppression | b = 0 points | b = 0 points | c = 0 points | a = 1 point | a = 1 point | 2 |
| **Ibiloye 2021[88]** | Nigeria | 2018 | Engagement in care | c = 0 points | b = 0 points | c = 0 points | c = 0 points | d = 0 points | 0 |
| **Afolaranmi 2021[89]** | Nigeria | 2019 | Engagement in care | a = 1 point | b = 0 points | b = 0 points | c = 0 points | c = 0 points | 1 |
| **Ndiaye 2013[90], Wade 2005[91]** | Senegal | 2004/ 2005 | HIV testing ever | a = 1 point | b = 0 points | a = 1 point | b = 0 points | c = 0 points | 2 |
| | | | Current ART use | a = 1 point | b = 0 points | a = 1 point | c = 0 points | c = 0 points | 2 |
| **Drame 2013[92]** | Senegal | 2012 | HIV testing ever | c = 0 points | b = 0 points | b = 0 points | b = 0 points | d = 0 points | 0 |
| | | | Knowledge of status | c = 0 points | b = 0 points | b = 0 points | b = 0 points | d = 0 points | 0 |
| | | | HIV incidence | c = 0 points | b = 0 points | b = 0 points | b = 0 points | a = 1 point | 1 |
| **Couderc 2017[32]** | Senegal | 2013 | HIV incidence | b = 0 points | b = 0 points | b = 0 points | c = 0 points | a = 1 point | 1 |
| **Lyons 2017[93], Lyons 2020[94]** | Senegal | 2015 | Knowledge of status | b = 0 points | b = 0 points | a = 1 point | b = 0 points | c = 0 points | 1 |

| References | Country | Midpoint Year | Outcomes reported | Criterion 1: Appropriateness of the sampling method to recruit a representative sample of MSM participants (maximum 1 point) | Criterion 2: Statistical adjustment of outcomes for complex sampling design (maximum 1 point) | Criterion 3: Generalizability of the results based on the study definition of MSM (maximum 1 point) | Criterion 4: Inclusion of transgender women in the study definition of MSM (maximum 1 point) | Criterion 5: Risk of misclassification in ascertainment of the relevant outcome(s) (maximum 1 point) | Study quality score /5 |
|---|---|---|---|---|---|---|---|---|---|
| | | | Engagement in care | b = 0 points | b = 0 points | a = 1 point | b = 0 points | c = 0 points | 1 |
| | | | Ever ART use | b = 0 points | b = 0 points | a = 1 point | b = 0 points | c = 0 points | 1 |
| | | | Current ART use | b = 0 points | b = 0 points | a = 1 point | b = 0 points | c = 0 points | 1 |
| | | | Viral suppression | b = 0 points | b = 0 points | a = 1 point | b = 0 points | a = 1 point | 2 |
| | | | HIV incidence | b = 0 points | b = 0 points | a = 1 point | b = 0 points | a = 1 point | 2 |
| **Mason 2013[95], Poteat 2017[21], Stahlman 2016[22]** | The Gambia | 2011 | Knowledge of status | a = 1 point | b = 0 points | a = 1 point | b = 0 points | c = 0 points | 2 |
| **Ekouevi 2014[96]** | Togo | 2011 | HIV testing ever | a = 1 point | b = 0 points | a = 1 point | c = 0 points | c = 0 points | 2 |
| **Bakai 2016[97]** | Togo | 2011 | HIV testing ever | a = 1 point | b = 0 points | c = 0 points | b = 0 points | c = 0 points | 1 |
| **Grosso 2019[18], Holland 2016[19], Poteat 2017[21], Ruiseñor-Escudero 2017[98], Ruiseñor-Escudero 2019[99], Ruiseñor-Escudero 2019[100], Stahlman 2016[22]** | Togo | 2013 | HIV testing ever | a = 1 point | b = 0 points | a = 1 point | b = 0 points | c = 0 points | 2 |
| | | | Knowledge of status | a = 1 point | b = 0 points | a = 1 point | b = 0 points | c = 0 points | 2 |
| | | | Current ART use | a = 1 point | b = 0 points | a = 1 point | b = 0 points | c = 0 points | 2 |
| **Teclessou 2017[101]** | Togo | 2015 | HIV testing ever | a = 1 point | b = 0 points | a = 1 point | a = 1 point | c = 0 points | 3 |
| | Togo | 2016 | HIV testing ever | b = 0 points | b = 0 points | b = 0 points | b = 0 points | c = 0 points | 0 |

| References | Country | Midpoint Year | Outcomes reported | Criterion 1: Appropriateness of the sampling method to recruit a representative sample of MSM participants (maximum 1 point) | Criterion 2: Statistical adjustment of outcomes for complex sampling design (maximum 1 point) | Criterion 3: Generalizability of the results based on the study definition of MSM (maximum 1 point) | Criterion 4: Inclusion of transgender women in the study definition of MSM (maximum 1 point) | Criterion 5: Risk of misclassification in ascertainment of the relevant outcome(s) (maximum 1 point) | Study quality score /5 |
|---|---|---|---|---|---|---|---|---|---|
| **Coulaud 2020[23], Dah 2021[24], Dah 2021[25], Laurent 2021[26], Yaya 2021[27], Yaya 2022[28]** | | | Engagement in care | b = 0 points | b = 0 points | b = 0 points | b = 0 points | c = 0 points | 0 |
| | | | Current ART use | b = 0 points | b = 0 points | b = 0 points | b = 0 points | c = 0 points | 0 |
| | | | HIV incidence | b = 0 points | b = 0 points | b = 0 points | b = 0 points | a = 1 point | 1 |
| **Sadio 2019[102]** | Togo | 2017 | HIV testing ever | a = 1 point | b = 0 points | a = 1 point | c = 0 points | c = 0 points | 2 |
| **Eastern Africa** | | | | | | | | | |
| **Coulaud 2016[103]** | Burundi | 2014 | HIV testing ever | b = 0 points | b = 0 points | b = 0 points | b = 0 points | b = 1 point | 1 |
| | | | HIV testing in the past 12 months | b = 0 points | b = 0 points | b = 0 points | b = 0 points | b = 1 point | 1 |
| | | | HIV testing in the past 6 months | b = 0 points | b = 0 points | b = 0 points | b = 0 points | b = 1 point | 1 |
| | | | HIV testing in the past 3 months | b = 0 points | b = 0 points | b = 0 points | b = 0 points | b = 1 point | 1 |
| **Lillie 2021[104]** | Burundi | 2018 | HIV testing ever | b = 0 points | b = 0 points | a = 1 point | a = 1 point | c = 0 points | 2 |
| | | | HIV testing in the past 6 months | b = 0 points | b = 0 points | a = 1 point | a = 1 point | c = 0 points | 2 |
| **Gebrebrhan 2021[105]** | Kenya | NR | Current ART use | b = 0 points | b = 0 points | b = 0 point | b = 0 points | d = 0 points | 0 |
| | | | Viral suppression | b = 0 points | b = 0 points | b = 0 point | b = 0 points | a = 1 point | 1 |
| **Sanders 2007[106]** | Kenya | 2006 | HIV testing ever | b = 0 points | b = 0 points | b = 0 points | b = 0 points | c = 0 points | 0 |
| | | | Knowledge of status | b = 0 points | b = 0 points | b = 0 points | b = 0 points | c = 0 points | 0 |
| **Kamali 2015[107], Price 2012[108]** | Kenya | 2007 | HIV incidence | b = 0 points | b = 0 points | c = 0 points | c = 0 points | a = 1 point | 1 |
| **Luchters 2011[109]** | Kenya | 2008 | HIV testing ever | a = 1 point | b = 0 points | b = 0 points | b = 0 points | c = 0 points | 1 |
| **Graham 2013[110]** | Kenya | 2008 | Engagement in care | a = 1 point | b = 0 points | a = 1 point | b = 0 points | c = 0 points | 2 |
| | | | Current ART use | a = 1 point | b = 0 points | a = 1 point | b = 0 points | c = 0 points | 2 |

| References | Country | Midpoint Year | Outcomes reported | Criterion 1: Appropriateness of the sampling method to recruit a representative sample of MSM participants (maximum 1 point) | Criterion 2: Statistical adjustment of outcomes for complex sampling design (maximum 1 point) | Criterion 3: Generalizability of the results based on the study definition of MSM (maximum 1 point) | Criterion 4: Inclusion of transgender women in the study definition of MSM (maximum 1 point) | Criterion 5: Risk of misclassification in ascertainment of the relevant outcome(s) (maximum 1 point) | Study quality score /5 |
|---|---|---|---|---|---|---|---|---|---|
| **Mdodo 2016[111]** | Kenya | 2010 | HIV incidence | a = 1 point | b = 0 points | b = 0 points | c = 0 points | a = 1 point | 2 |
| **Muraguri 2015[112]** | Kenya | 2010 | HIV testing ever | a = 1 point | a = 1 point | b = 0 points | b = 0 points | c = 0 points | 2 |
| | | | HIV testing in the past 12 months | a = 1 point | a = 1 point | b = 0 points | b = 0 points | c = 0 points | 2 |
| | | | Knowledge of status | a = 1 point | a = 1 point | b = 0 points | b = 0 points | c = 0 points | 2 |
| **McKinnon 2014[113]** | Kenya | 2010 | HIV testing ever | b = 0 points | b = 0 points | b = 0 points | c = 0 points | c = 0 points | 0 |
| | | | HIV incidence | b = 0 points | b = 0 points | b = 0 points | c = 0 points | a = 1 point | 1 |
| **Kamali 2015[107], Moller 2015[114], Price 2012[108], Sanders 2013[115], Wahome 2018[116], Wahome 2020[117]** | Kenya | 2011 | HIV testing ever | b = 0 points | b = 0 points | b = 0 points | b = 0 points | c = 0 points | 0 |
| | | | HIV incidence | b = 0 points | b = 0 points | b = 0 points | c = 0 points | a = 1 point | 1 |
| **Githuka 2016[118]** | Kenya | 2012 | HIV testing ever | a = 1 point | a = 1 point | a = 1 point | c = 0 points | c = 0 points | 3 |
| **Shangani 2017[119]** | Kenya | 2014 | HIV testing in the past 12 months | a = 1 point | b = 0 points | a = 1 point | b = 0 points | c = 0 points | 2 |
| **Bhattacharjee 2015[120], Musyoki 2018[121]** | Kenya | 2014/ 2015 | HIV testing ever | a = 1 point | b = 0 points | c = 0 points | b = 0 points | b = 1 point | 2 |
| | | | HIV testing in the past 3 months | a = 1 point | b = 0 points | c = 0 points | b = 0 points | b = 1 point | 2 |
| | | | Viral suppression | b = 0 points | b = 0 points | b = 0 points | b = 0 points | a = 1 point | 1 |
| **Nyblade 2017[122]** | Kenya | 2015 | HIV testing ever | a = 1 point | b = 0 points | b = 0 points | c = 0 points | c = 0 points | 1 |
| **Kimani 2019[123]** | Kenya | 2016 | HIV incidence | b = 0 points | b = 0 points | c = 0 points | a = 1 point | a = 1 point | 2 |
| **Korhonen 2018[124], Kunzweiler 2017[125],** | Kenya | 2016 | Knowledge of status | a = 1 point | b = 0 points | a = 1 point | c = 0 points | b = 1 point | 3 |
| | | | Ever ART use | a = 1 point | b = 0 points | a = 1 point | b = 0 points | b = 1 point | 3 |
| | | | Current ART use | a = 1 point | b = 0 points | [128]a = 1 point | c = 0 points | b = 1 point | 3 |

| References | Country | Midpoint Year | Outcomes reported | Criterion 1: Appropriateness of the sampling method to recruit a representative sample of MSM participants (maximum 1 point) | Criterion 2: Statistical adjustment of outcomes for complex sampling design (maximum 1 point) | Criterion 3: Generalizability of the results based on the study definition of MSM (maximum 1 point) | Criterion 4: Inclusion of transgender women in the study definition of MSM (maximum 1 point) | Criterion 5: Risk of misclassification in ascertainment of the relevant outcome(s) (maximum 1 point) | Study quality score /5 |
|---|---|---|---|---|---|---|---|---|---|
| **Kunzweiler 2018[126], Kunzweiler 2018[127]** | | | Viral suppression | a = 1 point | b = 0 points | a = 1 point | b = 0 points | a = 1 point | 3 |
| **Fogel 2019[128], Palumbo 2021[129], Sandfort 2019[130], Sandfort 2021[131], Sivay 2020[132], Zhang 2018[133]** | Kenya | 2016 | HIV testing ever | b = 0 points | b = 0 points | a = 1 point | b = 0 points | c = 0 points | 1 |
| | | | HIV testing in the past 12 months | b = 0 points | b = 0 points | a = 1 point | b = 0 points | c = 0 points | 1 |
| | | | HIV testing in the past 6 months | b = 0 points | b = 0 points | a = 1 point | b = 0 points | c = 0 points | 1 |
| | | | Knowledge of status | b = 0 points | b = 0 points | a = 1 point | b = 0 points | c = 0 points | 1 |
| | | | Engagement in care | b = 0 points | b = 0 points | a = 1 point | b = 0 points | c = 0 points | 1 |
| | | | Current ART use | b = 0 points | b = 0 points | a = 1 point | b = 0 points | c = 0 points | 1 |
| | | | Viral suppression | b = 0 points | b = 0 points | a = 1 point | b = 0 points | a = 1 point | 2 |
| | | | HIV incidence | b = 0 points | b = 0 points | a = 1 point | b = 0 points | a = 1 point | 2 |
| **Fearon 2020[134], Smith 2021[135], Smith 2021[136]** | Kenya | 2017 | HIV testing ever | a = 1 point | b = 0 points | a = 1 point | a = 1 point | b = 1 point | 4 |
| | | | HIV testing in the past 6 months | a = 1 point | b = 0 points | a = 1 point | b = 0 points | b = 1 point | 3 |
| | | | Knowledge of status | a = 1 point | b = 0 points | a = 1 point | b = 0 points | b = 1 point | 3 |
| | | | Engagement in care | a = 1 point | b = 0 points | a = 1 point | b = 0 points | b = 1 point | 3 |
| | | | Current ART use | a = 1 point | b = 0 points | a = 1 point | b = 0 points | b = 1 point | 3 |
| | | | Viral suppression | a = 1 point | b = 0 points | a = 1 point | b = 0 points | a = 1 point | 3 |

| References | Country | Midpoint Year | Outcomes reported | Criterion 1: Appropriateness of the sampling method to recruit a representative sample of MSM participants (maximum 1 point) | Criterion 2: Statistical adjustment of outcomes for complex sampling design (maximum 1 point) | Criterion 3: Generalizability of the results based on the study definition of MSM (maximum 1 point) | Criterion 4: Inclusion of transgender women in the study definition of MSM (maximum 1 point) | Criterion 5: Risk of misclassification in ascertainment of the relevant outcome(s) (maximum 1 point) | Study quality score /5 |
|---|---|---|---|---|---|---|---|---|---|
| **Sanders 2013[115], Wahome 2020[117]** | Kenya | 2018 | HIV incidence | b = 0 points | b = 0 points | b = 0 points | c = 0 points | a = 1 point | 1 |
| **Bhattacharjee 2020[137]** | Kenya | 2019 | HIV testing ever | a = 1 point | b = 0 points | a = 1 point | b = 0 points | c = 0 points | 2 |
| | | | HIV testing in the past 12 months | a = 1 point | b = 0 points | a = 1 point | b = 0 points | c = 0 points | 2 |
| | | | HIV testing in the past 6 months | a = 1 point | b = 0 points | a = 1 point | b = 0 points | c = 0 points | 2 |
| | | | HIV testing in the past 3 months | a = 1 point | b = 0 points | a = 1 point | b = 0 points | c = 0 points | 2 |
| | | | Knowledge of status | a = 1 point | b = 0 points | a = 1 point | b = 0 points | c = 0 points | 2 |
| | | | Engagement in care | a = 1 point | b = 0 points | a = 1 point | b = 0 points | c = 0 points | 2 |
| | | | Current ART use | a = 1 point | b = 0 points | a = 1 point | b = 0 points | c = 0 points | 2 |
| **Dijkstra 2021[138]** | Kenya | 2019 | HIV testing ever | b = 0 points | b = 0 points | a = 1 point | b = 0 points | c = 0 points | 1 |
| | | | HIV testing in the past 12 months | b = 0 points | b = 0 points | a = 1 point | b = 0 points | c = 0 points | 1 |
| | | | HIV testing in the past 3 months | b = 0 points | b = 0 points | a = 1 point | b = 0 points | c = 0 points | 1 |
| | | | Knowledge of status | b = 0 points | b = 0 points | a = 1 point | b = 0 points | c = 0 points | 1 |
| | | | Current ART use | b = 0 points | b = 0 points | a = 1 point | b = 0 points | c = 0 points | 1 |
| | | | Viral suppression | b = 0 points | b = 0 points | a = 1 point | b = 0 points | a = 1 point | 2 |

| References | Country | Midpoint Year | Outcomes reported | Criterion 1: Appropriateness of the sampling method to recruit a representative sample of MSM participants (maximum 1 point) | Criterion 2: Statistical adjustment of outcomes for complex sampling design (maximum 1 point) | Criterion 3: Generalizability of the results based on the study definition of MSM (maximum 1 point) | Criterion 4: Inclusion of transgender women in the study definition of MSM (maximum 1 point) | Criterion 5: Risk of misclassification in ascertainment of the relevant outcome(s) (maximum 1 point) | Study quality score /5 |
|---|---|---|---|---|---|---|---|---|---|
| **Virkud 2020[139]** | Kenya, Rwanda, Tanzania, Uganda (cross-border areas) | 2016 | HIV testing in the past 12 months | b = 0 points | b = 0 points | a = 1 point | c = 0 points | c = 0 points | 1 |
| **Ntata 2008[140]** | Malawi | 2006 | HIV testing ever | a = 1 point | b = 0 points | c = 0 points | c = 0 points | c = 0 points | 1 |
| **Baral 2009[141], Beyrer 2010[142], Fay 2011[143]** | Malawi | 2008 | HIV testing ever | a = 1 point | b = 0 points | a = 1 point | b = 0 points | c = 0 points | 2 |
| | | | Knowledge of status | a = 1 point | b = 0 points | a = 1 point | b = 0 points | c = 0 points | 2 |
| | | | Current ART use | a = 1 point | b = 0 points | a = 1 point | b = 0 points | c = 0 points | 2 |
| **Poteat 2017[21], Stahlman 2016[144], Wirtz 2013[145], Wirtz 2015[146], Wirtz 2017[147]** | Malawi | 2013 | HIV testing ever | a = 1 point | a = 1 point | a = 1 point | b = 0 points | c = 0 points | 3 |
| | | | HIV testing in the past 12 months | a = 1 point | a = 1 point | a = 1 point | b = 0 points | c = 0 points | 3 |
| | | | Knowledge of status | a = 1 point | a = 1 point | a = 1 point | b = 0 points | c = 0 points | 3 |
| | | | Ever ART use | a = 1 point | a = 1 point | a = 1 point | b = 0 points | c = 0 points | 3 |
| | | | HIV incidence | a = 1 point | b = 0 points | a = 1 point | b = 0 points | a = 1 point | 3 |
| **Fogel 2019[128], Palumbo 2021[129], Sandfort 2019[130], Sandfort 2021[131], Sivay 2020[132], Zhang 2018[133]** | Malawi | 2016 | HIV testing ever | b = 0 points | b = 0 points | a = 1 point | b = 0 points | c = 0 points | 1 |
| | | | HIV testing in the past 12 months | b = 0 points | b = 0 points | a = 1 point | b = 0 points | c = 0 points | 1 |
| | | | HIV testing in the past 6 months | b = 0 points | b = 0 points | a = 1 point | b = 0 points | c = 0 points | 1 |
| | | | Knowledge of status | b = 0 points | b = 0 points | a = 1 point | b = 0 points | c = 0 points | 1 |
| | | | Engagement in care | b = 0 points | b = 0 points | a = 1 point | b = 0 points | c = 0 points | 1 |

| References | Country | Midpoint Year | Outcomes reported | Criterion 1: Appropriateness of the sampling method to recruit a representative sample of MSM participants (maximum 1 point) | Criterion 2: Statistical adjustment of outcomes for complex sampling design (maximum 1 point) | Criterion 3: Generalizability of the results based on the study definition of MSM (maximum 1 point) | Criterion 4: Inclusion of transgender women in the study definition of MSM (maximum 1 point) | Criterion 5: Risk of misclassification in ascertainment of the relevant outcome(s) (maximum 1 point) | Study quality score /5 |
|---|---|---|---|---|---|---|---|---|---|
| | | | Current ART use | b = 0 points | b = 0 points | a = 1 point | b = 0 points | c = 0 points | 1 |
| | | | Viral suppression | b = 0 points | b = 0 points | a = 1 point | b = 0 points | a = 1 point | 2 |
| | | | HIV incidence | b = 0 points | b = 0 points | a = 1 point | b = 0 points | a = 1 point | 2 |
| **Herce 2018[4]** | Malawi | 2017 | HIV testing ever | a = 1 point | a = 1 point | b = 0 points | a = 1 point | c = 0 points | 3 |
| | | | HIV testing in the past 6 months | a = 1 point | a = 1 point | b = 0 points | a = 1 point | c = 0 points | 3 |
| **Rucinski 2022[148]** | Malawi | 2018 | Engagement in care | b = 0 points | b = 0 points | c = 0 points | c = 0 points | d = 0 points | 0 |
| **Adam 2009[51]** | Mauritius | 2004 | HIV testing in the past 12 months | c = 0 points | b = 0 points | c = 0 points | c = 0 points | d = 0 points | 0 |
| **Boothe 2021[149], Boothe 2021[150], Horth 2015[151], Sathane 2016[152]** | Mozambique | 2011 | HIV testing ever | a = 1 point | b = 0 points | a = 1 point | b = 0 points | c = 0 points | 2 |
| | | | HIV testing in the past 12 months | a = 1 point | a = 1 point | a = 1 point | b = 0 points | c = 0 points | 3 |
| | | | Knowledge of status | a = 1 point | b = 0 points | a = 1 point | b = 0 points | c = 0 points | 2 |
| | | | Engagement in care | a = 1 point | b = 0 points | a = 1 point | b = 0 points | c = 0 points | 2 |
| | | | Ever ART use | a = 1 point | b = 0 points | a = 1 point | b = 0 points | c = 0 points | 2 |
| | | | Current ART use | a = 1 point | b = 0 points | a = 1 point | b = 0 points | c = 0 points | 2 |
| **Chapman 2011[153]** | Rwanda | 2009 | HIV testing ever | a = 1 point | b = 0 points | a = 1 point | b = 0 points | c = 0 points | 2 |
| **Ntale 2019[154]** | Rwanda | 2015 | HIV testing in the past 12 months | a = 1 point | b = 0 points | a = 1 point | c = 0 points | c = 0 points | 2 |
| **Twahirwa Rwema 2020[155]** | Rwanda | 2018 | HIV testing ever | a = 1 point | b = 0 points | a = 1 point | b = 0 points | c = 0 points | 2 |
| | | | Knowledge of status | a = 1 point | b = 0 points | a = 1 point | b = 0 points | c = 0 points | 2 |
| | | | Current ART use | a = 1 point | b = 0 points | a = 1 point | b = 0 points | c = 0 points | 2 |
| | | | Viral suppression | a = 1 point | b = 0 points | a = 1 point | b = 0 points | a = 1 point | 3 |

| References | Country | Midpoint Year | Outcomes reported | Criterion 1: Appropriateness of the sampling method to recruit a representative sample of MSM participants (maximum 1 point) | Criterion 2: Statistical adjustment of outcomes for complex sampling design (maximum 1 point) | Criterion 3: Generalizability of the results based on the study definition of MSM (maximum 1 point) | Criterion 4: Inclusion of transgender women in the study definition of MSM (maximum 1 point) | Criterion 5: Risk of misclassification in ascertainment of the relevant outcome(s) (maximum 1 point) | Study quality score /5 |
|---|---|---|---|---|---|---|---|---|---|
| **Magesa 2014[156]** | Tanzania | NR | HIV testing ever | a = 1 point | b = 0 points | a = 1 point | b = 0 points | c = 0 points | 2 |
| **Dahoma 2011[157], Johnston 2010[158], Khatib 2017[159]** | Tanzania | 2007 | HIV testing ever | a = 1 point | a = 1 point | b = 0 points | b = 0 points | c = 0 points | 2 |
| | | | HIV testing in the past 12 months | a = 1 point | a = 1 point | b = 0 points | b = 0 points | c = 0 points | 2 |
| **Nyoni 2012[160], Nyoni 2013[161]** | Tanzania | 2009 | HIV testing ever | a = 1 point | b = 0 points | a = 1 point | b = 0 points | c = 0 points | 2 |
| **Khatib 2017[159]** | Tanzania | 2011 | HIV testing ever | a = 1 point | a = 1 point | b = 0 points | b = 0 points | c = 0 points | 2 |
| | | | HIV testing in the past 12 months | a = 1 point | a = 1 point | b = 0 points | b = 0 points | c = 0 points | 2 |
| **Mmbaga 2012[162]** | Tanzania | 2011 | HIV testing ever | a = 1 point | b = 0 points | a = 1 point | c = 0 points | c = 0 points | 2 |
| **Ahaneku 2016[163], Anderson 2015[164], Romijnders 2016[165], Ross 2014[166]** | Tanzania | 2012 | HIV testing ever | a = 1 point | b = 0 points | a = 1 point | b = 0 points | b = 1 point | 3 |
| | | | Knowledge of status | a = 1 point | b = 0 points | a = 1 point | b = 0 points | b = 1 point | 3 |
| **Mmbaga 2018[167]** | Tanzania | 2014 | HIV testing ever | a = 1 point | b = 0 points | a = 1 point | b = 0 points | c = 0 points | 2 |
| **Ross 2019[168]** | Tanzania | 2015 | HIV testing ever | b = 0 points | b = 0 points | a = 1 point | c = 0 points | c = 0 points | 1 |
| | | | HIV testing in the past 6 months | b = 0 points | b = 0 points | a = 1 point | c = 0 points | c = 0 points | 1 |
| **Kajubi 2008[169], Raymond 2009[170]** | Uganda | 2004 | HIV testing ever | a = 1 point | a = 1 point | b = 0 points | b = 0 points | c = 0 points | 2 |
| | | | HIV testing in the past 6 months | a = 1 point | a = 1 point | b = 0 points | b = 0 points | c = 0 points | 2 |

| References | Country | Midpoint Year | Outcomes reported | Criterion 1: Appropriateness of the sampling method to recruit a representative sample of MSM participants (maximum 1 point) | Criterion 2: Statistical adjustment of outcomes for complex sampling design (maximum 1 point) | Criterion 3: Generalizability of the results based on the study definition of MSM (maximum 1 point) | Criterion 4: Inclusion of transgender women in the study definition of MSM (maximum 1 point) | Criterion 5: Risk of misclassification in ascertainment of the relevant outcome(s) (maximum 1 point) | Study quality score /5 |
|---|---|---|---|---|---|---|---|---|---|
| **Hladik 2012[171]** | Uganda | 2008 | HIV testing ever | a = 1 point | a = 1 point | b = 0 points | b = 0 points | b = 1 point | 3 |
| **Robb 2016[172]** | Uganda | 2012 | HIV incidence | b = 0 points | b = 0 points | b = 0 points | c = 0 points | a = 1 point | 1 |
| **Wanyenze 2016[173]** | Uganda | 2013 | HIV testing ever | a = 1 point | b = 0 points | b = 0 points | b = 0 points | c = 0 points | 1 |
| **Hladik 2017[174]** | Uganda | 2013 | HIV testing ever | a = 1 point | a = 1 point | a = 1 point | b = 0 points | b = 1 point | 4 |
| | | | HIV testing in the past 12 months | a = 1 point | a = 1 point | a = 1 point | b = 0 points | b = 1 point | 4 |
| | | | Knowledge of status | a = 1 point | b = 0 points | a = 1 point | b = 0 points | b = 1 point | 3 |
| | | | Current ART use | a = 1 point | b = 0 points | a = 1 point | b = 0 points | b = 1 point | 3 |
| | | | Viral suppression | a = 1 point | a = 1 point | a = 1 point | b = 0 points | a = 1 point | 4 |
| **Okoboi 2020[175], Okoboi 2021[176]** | Uganda | 2018 | HIV testing ever | a = 1 point | b = 0 points | c = 0 points | c = 0 points | c = 0 points | 1 |
| | | | Knowledge of status | a = 1 point | b = 0 points | c = 0 points | c = 0 points | c = 0 points | 1 |
| **Harris 2022[177], Parmley 2022[178], Parmley 2022[179]** | Zimbabwe | 2019 | HIV testing ever | a = 1 point | b = 0 points | a = 1 point | b = 0 points | c = 0 points | 2 |
| | | | Knowledge of status | a = 1 point | b = 0 points | a = 1 point | b = 0 points | c = 0 points | 2 |
| | | | Current ART use | a = 1 point | b = 0 points | a = 1 point | a = 1 point | c = 0 points | 3 |
| | | | Viral suppression | a = 1 point | b = 0 points | a = 1 point | b = 0 points | a = 1 point | 3 |
| **Southern Africa** | | | | | | | | | |
| **Baral 2009[141], Beyrer 2010[142], Fay 2011[143]** | Botswana | 2008 | HIV testing ever | a = 1 point | b = 0 points | a = 1 point | b = 0 points | c = 0 points | 2 |
| | | | Knowledge of status | a = 1 point | b = 0 points | a = 1 point | b = 0 points | c = 0 points | 2 |
| | | | Current ART use | a = 1 point | b = 0 points | a = 1 point | b = 0 points | c = 0 points | 2 |

| References | Country | Midpoint Year | Outcomes reported | Criterion 1: Appropriateness of the sampling method to recruit a representative sample of MSM participants (maximum 1 point) | Criterion 2: Statistical adjustment of outcomes for complex sampling design (maximum 1 point) | Criterion 3: Generalizability of the results based on the study definition of MSM (maximum 1 point) | Criterion 4: Inclusion of transgender women in the study definition of MSM (maximum 1 point) | Criterion 5: Risk of misclassification in ascertainment of the relevant outcome(s) (maximum 1 point) | Study quality score /5 |
|---|---|---|---|---|---|---|---|---|---|
| **Baral 2013[180], Brown 2016[181], Grover 2016[182], Poteat 2017[21], Rao 2017[8], Risher 2013[183], Stahlman 2015[184], Stahlman 2016[144]** | eSwatini | 2011 | HIV testing in the past 12 months | a = 1 point | a = 1 point | a = 1 point | b = 0 points | c = 0 points | 3 |
| | | | Knowledge of status | a = 1 point | a = 1 point | a = 1 point | b = 0 points | c = 0 points | 3 |
| **Rao 2017[8]** | eSwatini | 2014 | HIV testing in the past 12 months | a = 1 point | b = 0 points | a = 1 point | b = 0 points | c = 0 points | 2 |
| **Baral 2011[185]** | Lesotho | 2009 | HIV testing in the past 12 months | a = 1 point | b = 0 points | a = 1 point | a = 1 point | c = 0 points | 3 |
| **Poteat 2017[21], Stahlman 2015[184], Stahlman 2015[186], Stahlman 2016[144], Wendi 2016[187]** | Lesotho | 2014 | HIV testing ever | a = 1 point | a = 1 point | a = 1 point | b = 0 points | c = 0 points | 3 |
| | | | Knowledge of status | a = 1 point | a = 1 point | a = 1 point | b = 0 points | c = 0 points | 3 |
| **Baral 2009[141], Beyrer 2010[142], Fay 2011[143]** | Namibia | 2008 | HIV testing ever | a = 1 point | b = 0 points | a = 1 point | b = 0 points | c = 0 points | 2 |
| | | | Knowledge of status | a = 1 point | b = 0 points | a = 1 point | b = 0 points | c = 0 points | 2 |
| | | | Current ART use | a = 1 point | b = 0 points | a = 1 point | b = 0 points | c = 0 points | 2 |
| **Russell 2019[188]** | Namibia | 2016 | HIV testing in the past 6 months | b = 0 points | b = 0 points | c = 0 points | a = 1 point | c = 0 points | 1 |
| **Cloete 2008[189]** | South Africa | NR | Current ART use | b = 0 points | b = 0 points | c = 0 points | c = 0 points | b = 1 point | 1 |
| **Jobson 2018[190]** | South Africa | NR | HIV testing ever | a = 1 point | b = 0 points | b = 0 points | b = 0 points | b = 1 point | 2 |
| **Lane 2008[191]** | South Africa | 2004 | HIV testing ever | b = 0 points | b = 0 points | a = 1 point | b = 0 points | c = 0 points | 1 |

| References | Country | Midpoint Year | Outcomes reported | Criterion 1: Appropriateness of the sampling method to recruit a representative sample of MSM participants (maximum 1 point) | Criterion 2: Statistical adjustment of outcomes for complex sampling design (maximum 1 point) | Criterion 3: Generalizability of the results based on the study definition of MSM (maximum 1 point) | Criterion 4: Inclusion of transgender women in the study definition of MSM (maximum 1 point) | Criterion 5: Risk of misclassification in ascertainment of the relevant outcome(s) (maximum 1 point) | Study quality score /5 |
|---|---|---|---|---|---|---|---|---|---|
| | | | HIV testing in the past 6 months | b = 0 points | b = 0 points | a = 1 point | b = 0 points | c = 0 points | 1 |
| **Nel 2013[192], Sandfort 2008[193]** | South Africa | 2004 | HIV testing ever | b = 0 points | b = 0 points | b = 0 points | c = 0 points | c = 1 point | 1 |
| **Burrell 2010[194]** | South Africa | 2008 | HIV testing in the past 12 months | b = 0 points | b = 0 points | b = 0 points | b = 0 points | b = 1 point | 1 |
| **Knox 2011[195], Knox 2013[196]** | South Africa | 2008 | HIV testing ever | b = 0 points | b = 0 points | a = 1 point | b = 0 points | b = 1 point | 2 |
| | | | HIV testing in the past 12 months | b = 0 points | b = 0 points | a = 1 point | b = 0 points | b = 1 point | 2 |
| **Arnold 2013[197], Lane 2011[198]** | South Africa | 2008 | HIV testing ever | a = 1 point | b = 0 points | a = 1 point | b = 0 points | c = 0 points | 2 |
| | | | Knowledge of status | a = 1 point | b = 0 points | a = 1 point | b = 0 points | c = 0 points | 2 |
| **Baral 2011[199]** | South Africa | 2009 | Knowledge of status | b = 0 points | b = 0 points | a = 1 point | a = 1 point | c = 0 points | 2 |
| **Buchbinder 2014[200], Buchbinder 2014[201]** | South Africa | 2009 | HIV incidence | b = 0 points | b = 0 points | b = 0 points | b = 0 points | a = 1 point | 1 |
| **Tun 2012[202]** | South Africa | 2009 | HIV testing ever | a = 1 point | a = 1 point | c = 0 points | b = 0 points | c = 0 points | 2 |
| **Eaton 2013[203]** | South Africa | 2010 | HIV testing ever | b = 0 points | b = 0 points | b = 0 points | b = 0 points | b = 1 point | 1 |
| **Kamali 2015[107]** | South Africa | 2010 | HIV incidence | b = 0 points | b = 0 points | c = 0 points | c = 0 points | a = 1 point | 1 |
| **Stephenson 2012[204], Wagenaar 2012[205]** | South Africa | 2010 | HIV testing ever | b = 0 points | b = 0 points | a = 1 point | b = 0 points | b = 1 point | 2 |
| **Batist 2013[206]** | South Africa | 2012 | HIV testing ever | b = 0 points | b = 0 points | b = 0 points | b = 0 points | b = 1 point | 1 |
| **Knox 2019[207]** | South Africa | 2012 | HIV testing in the past 6 months | a = 1 point | b = 0 points | a = 1 point | c = 0 points | c = 0 points | 2 |
| **Maleke 2017[208]** | South Africa | 2012 | HIV testing ever | a = 1 point | b = 0 points | b = 0 points | c = 0 points | c = 0 points | 1 |

| References | Country | Midpoint Year | Outcomes reported | Criterion 1: Appropriateness of the sampling method to recruit a representative sample of MSM participants (maximum 1 point) | Criterion 2: Statistical adjustment of outcomes for complex sampling design (maximum 1 point) | Criterion 3: Generalizability of the results based on the study definition of MSM (maximum 1 point) | Criterion 4: Inclusion of transgender women in the study definition of MSM (maximum 1 point) | Criterion 5: Risk of misclassification in ascertainment of the relevant outcome(s) (maximum 1 point) | Study quality score /5 |
|---|---|---|---|---|---|---|---|---|---|
| **Rebe 2015[209]** | South Africa | 2012 | HIV testing in the past 12 months | b = 0 points | b = 0 points | b = 0 points | b = 0 points | c = 0 points | 0 |
| | | | Current ART use | b = 0 points | b = 0 points | b = 0 points | b = 0 points | c = 0 points | 0 |
| **Siegler 2015[210]** | South Africa | 2012 | HIV testing ever | a = 1 point | b = 0 points | a = 1 point | b = 0 points | c = 0 points | 2 |
| **Lane 2014[211], Lane 2016[212]** | South Africa | 2012 | HIV testing ever | a = 1 point | a = 1 point | a = 1 point | b = 0 points | c = 0 points | 3 |
| | | | Knowledge of status | a = 1 point | b = 0 points | a = 1 point | b = 0 points | c = 0 points | 2 |
| | | | Engagement in care | a = 1 point | b = 0 points | a = 1 point | b = 0 points | c = 0 points | 2 |
| | | | Current ART use | a = 1 point | b = 0 points | a = 1 point | b = 0 points | c = 0 points | 2 |
| | | 2015 | HIV incidence | a = 1 point | b = 0 points | a = 1 point | c = 0 points | a = 1 point | 3 |
| **Maenetje 2019[213]** | South Africa | 2013 | HIV incidence | b = 0 points | b = 0 points | b = 0 points | c = 0 points | a = 1 point | 1 |
| **Kufa 2017[214]** | South Africa | 2015 | Viral suppression | a = 1 point | b = 0 points | a = 1 point | b = 0 points | c = 0 points | 2 |
| **Rees 2017[215], van Liere 2017[216]** | South Africa | 2015 | Knowledge of status | b = 0 points | b = 0 points | b = 0 points | c = 0 points | c = 0 points | 0 |
| | | | Current ART use | b = 0 points | b = 0 points | b = 0 points | c = 0 points | c = 0 points | 0 |
| **Chen 2020[217], Lippman 2018[218], Lippman 2018[219], Radebe 2020[220]** | South Africa | 2015/ 2016 | HIV testing ever | a = 1 point | b = 0 points | a = 1 point | b = 0 points | c = 0 points | 2 |
| | | | HIV testing in the past 12 months | a = 1 point | b = 0 points | a = 1 point | b = 0 points | c = 0 points | 2 |
| | | | HIV testing in the past 6 months | a = 1 point | b = 0 points | a = 1 point | b = 0 points | c = 0 points | 2 |
| | | | HIV incidence | a = 1 point | b = 0 points | a = 1 point | b = 0 points | a = 1 point | 3 |
| **Fogel 2018[128], Palumbo 2021[129], Sandfort 2019[130], Sandfort 2021[131], Sivay** | South Africa | 2016 | HIV testing ever | b = 0 points | b = 0 points | a = 1 point | b = 0 points | c = 0 points | 1 |
| | | | HIV testing in the past 6 months | b = 0 points | b = 0 points | a = 1 point | b = 0 points | c = 0 points | 1 |
| | | | Knowledge of status | b = 0 points | b = 0 points | a = 1 point | b = 0 points | c = 0 points | 1 |
| | | | Engagement in care | b = 0 points | b = 0 points | a = 1 point | b = 0 points | c = 0 points | 1 |

| References | Country | Midpoint Year | Outcomes reported | Criterion 1: Appropriateness of the sampling method to recruit a representative sample of MSM participants (maximum 1 point) | Criterion 2: Statistical adjustment of outcomes for complex sampling design (maximum 1 point) | Criterion 3: Generalizability of the results based on the study definition of MSM (maximum 1 point) | Criterion 4: Inclusion of transgender women in the study definition of MSM (maximum 1 point) | Criterion 5: Risk of misclassification in ascertainment of the relevant outcome(s) (maximum 1 point) | Study quality score /5 |
|---|---|---|---|---|---|---|---|---|---|
| **2020[132], Zhang 2018[133]** | | | Current ART use | b = 0 points | b = 0 points | a = 1 point | b = 0 points | c = 0 points | 1 |
| | | | Viral suppression | b = 0 points | b = 0 points | a = 1 point | b = 0 points | a = 1 point | 2 |
| | | | HIV incidence | b = 0 points | b = 0 points | a = 1 point | b = 0 points | a = 1 point | 2 |
| **Sullivan 2020[221]** | South Africa | 2016 | Knowledge of status | b = 0 points | b = 0 points | a = 1 point | b = 0 points | b = 1 point | 2 |
| | | | HIV incidence | b = 0 points | b = 0 points | a = 1 point | a = 1 point | a = 1 point | 3 |
| **Fearon 2020[222]** | South Africa | 2017 | HIV testing ever | a = 1 point | a = 1 point | a = 1 point | b = 0 points | b = 1 point | 4 |
| | | | HIV testing in the past 12 months | a = 1 point | a = 1 point | a = 1 point | b = 0 points | b = 1 point | 4 |
| | | | HIV testing in the past 6 months | a = 1 point | a = 1 point | a = 1 point | b = 0 points | b = 1 point | 4 |
| | | | HIV testing in the past 3 months | a = 1 point | a = 1 point | a = 1 point | b = 0 points | b = 1 point | 4 |
| | | | Knowledge of status | a = 1 point | a = 1 point | a = 1 point | b = 0 points | b = 1 point | 4 |
| | | | Current ART use | a = 1 point | a = 1 point | a = 1 point | b = 0 points | b = 1 point | 4 |
| | | | Viral suppression | a = 1 point | a = 1 point | a = 1 point | b = 0 points | a = 1 point | 4 |
| **Fearon 2020[134]** | South Africa | 2017 | HIV testing in the past 6 months | a = 1 point | b = 0 points | a = 1 point | b = 0 points | b = 1 point | 3 |
| | | | Knowledge of status | a = 1 point | b = 0 points | a = 1 point | b = 0 points | b = 1 point | 3 |
| | | | Current ART use | a = 1 point | b = 0 points | a = 1 point | b = 0 points | b = 1 point | 3 |
| | | | Viral suppression | a = 1 point | b = 0 points | a = 1 point | b = 0 points | a = 1 point | 3 |
| **Scheibe 2020[223]** | South Africa | 2017 | HIV testing ever | b = 0 points | b = 0 points | a = 1 point | b = 0 points | c = 0 points | 1 |
| | | | Knowledge of status | b = 0 points | b = 0 points | a = 1 point | b = 0 points | c = 0 points | 1 |
| | | | Current ART use | b = 0 points | b = 0 points | a = 1 point | b = 0 points | c = 0 points | 1 |
| **Pillay 2020[224]** | South Africa | 2018 | HIV testing ever | b = 0 points | b = 0 points | b = 0 points | b = 0 points | c = 0 points | 0 |

| References | Country | Midpoint Year | Outcomes reported | Criterion 1: Appropriateness of the sampling method to recruit a representative sample of MSM participants (maximum 1 point) | Criterion 2: Statistical adjustment of outcomes for complex sampling design (maximum 1 point) | Criterion 3: Generalizability of the results based on the study definition of MSM (maximum 1 point) | Criterion 4: Inclusion of transgender women in the study definition of MSM (maximum 1 point) | Criterion 5: Risk of misclassification in ascertainment of the relevant outcome(s) (maximum 1 point) | Study quality score /5 |
|---|---|---|---|---|---|---|---|---|---|
| | | | HIV testing in the past 12 months | b = 0 points | b = 0 points | b = 0 points | b = 0 points | c = 0 points | 0 |
| | | | HIV testing in the past 6 months | b = 0 points | b = 0 points | b = 0 points | b = 0 points | c = 0 points | 0 |
| | | | HIV testing in the past 3 months | b = 0 points | b = 0 points | b = 0 points | b = 0 points | c = 0 points | 0 |
| **Minnis 2020[225], Montgomery 2021[226]** | South Africa | 2018 | HIV testing ever | b = 0 points | b = 0 points | b = 0 points | c = 0 points | c = 0 points | 0 |
| **Metheny 2022[227], Stephenson 2021[228], Stephenson 2022[229]** | South Africa, Namibia | 2017 | HIV testing ever | b = 0 points | b = 0 points | b = 0 points | c = 0 points | c = 0 points | 0 |
| | | | HIV testing in the past 6 months | b = 0 points | b = 0 points | b = 0 points | c = 0 points | c = 0 points | 0 |

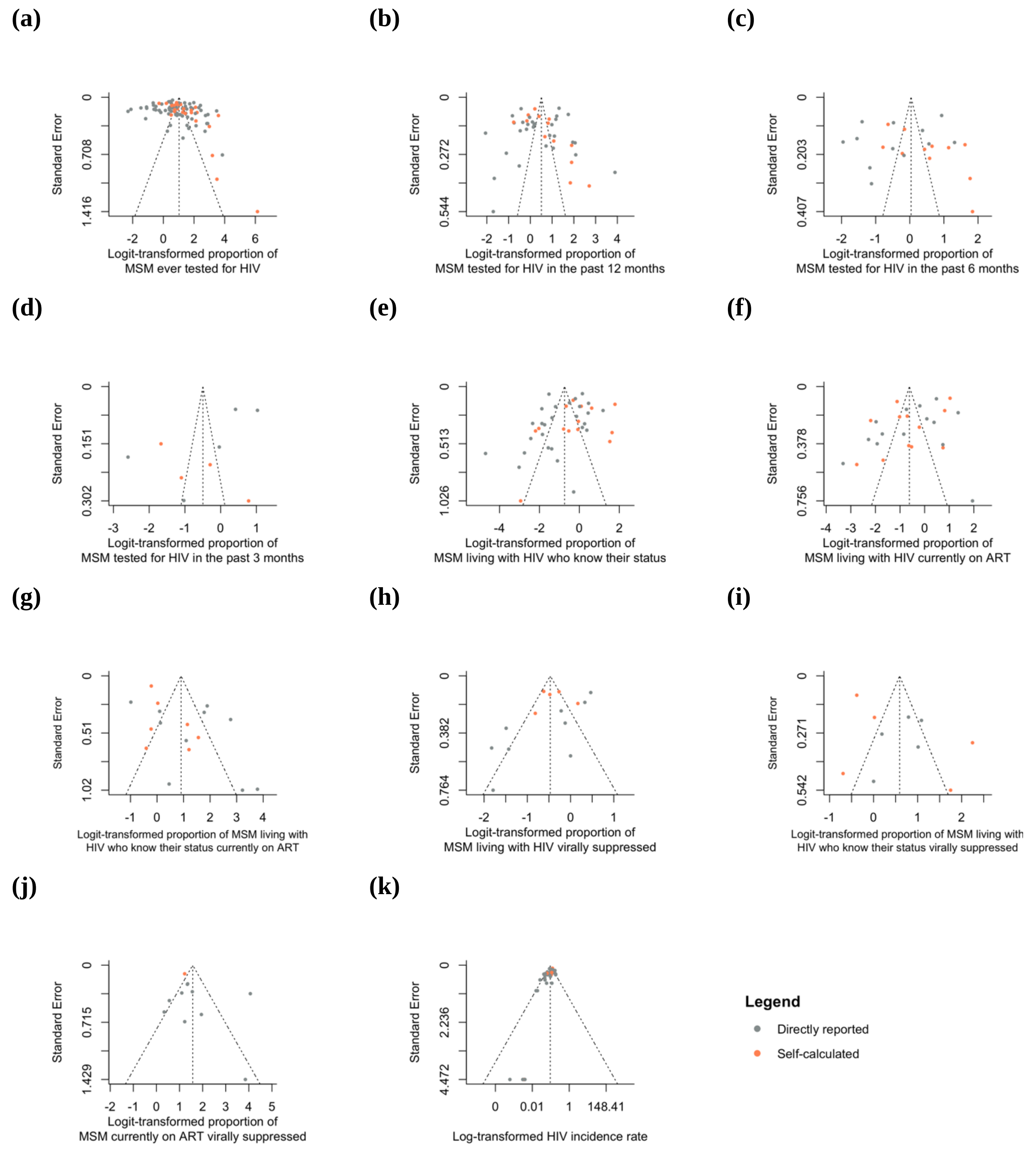


**Figure S28. Funnel plots of HIV testing, treatment cascade, and HIV incidence outcomes among men who have sex with men (MSM) in Africa.** Funnel plots of (a) ever HIV testing, (b) HIV testing in the past 12 months, (c) HIV testing in the past 6 months, (d) HIV testing in the past 3 months, (e) knowledge of status among MSM living with HIV, (f) current antiretroviral therapy (ART) use among MSM living with HIV, (g) current ART use among HIV aware MSM, (h) viral suppression among MSM living with HIV, (i) viral suppression among HIV aware MSM, (j) viral suppression among MSM currently on ART, and (k) HIV incidence among MSM. Points represent study observations that were either directly reported in articles (grey points) or that we calculated from available information reported in articles (orange points).

**References:**


1. Valadez JJ, Berendes S, Jeffery C, et al. Filling the Knowledge Gap: Measuring HIV Prevalence and Risk Factors among Men Who Have Sex with Men and Female Sex Workers in Tripoli, Libya. *PLoS One* 2013; **8**(6): e66701.
2. Elmahy AG. Reaching Egyptian Gays Using Social Media: A Comprehensive Health Study and a Framework for Future Research. *J Homosex* 2018; **65**(13): 1867-76.
3. Kendall C, Kerr LRFS, Mota RMS, et al. Population size, HIV, and behavior among MSM in Luanda, Angola: challenges and findings in the first ever HIV and syphilis biological and behavioral survey. *Journal of acquired immune deficiency syndromes (1999)* 2014; **66**(5): 544.
4. Herce ME, Miller WM, Bula A, et al. Achieving the first 90 for key populations in sub-Saharan Africa through venue-based outreach: challenges and opportunities for HIV prevention based on PLACE study findings from Malawi and Angola. *J Int AIDS Soc* 2018; **21 Suppl 5**(Suppl Suppl 5): e25132.
5. Lorente N, Henry E, Fugon L, et al. Proximity to HIV is associated with a high rate of HIV testing among men who have sex with men living in Douala, Cameroon. *AIDS care* 2012; **24**(8): 1020-7.
6. Holland CE, Papworth E, Billong SC, et al. Access to HIV services at non-governmental and community-based organizations among men who have sex with men (MSM) in Cameroon: An integrated biological and behavioral surveillance analysis. *PLoS One* 2015; **10**(4): e0122881.
7. Park JN, Papworth E, Billong SC, et al. Correlates of prior HIV testing among men who have sex with men in Cameroon: a cross-sectional analysis. *BMC Public Health* 2014; **14**: 1220.
8. Rao A, Stahlman S, Hargreaves J, et al. Sampling Key Populations for HIV Surveillance: Results From Eight Cross-Sectional Studies Using Respondent-Driven Sampling and Venue-Based Snowball Sampling. *JMIR Public Health Surveill* 2017; **3**(4): e72.
9. Bowring AL, Ketende S, Rao A, et al. Characterising unmet HIV prevention and treatment needs among young female sex workers and young men who have sex with men in Cameroon: a cross-sectional analysis. *Lancet Child Adolesc Health* 2019; **3**(7): 482-91.
10. Mboumba Bouassa RS, Mbeko Simaleko M, Camengo SP, et al. Unusual and unique distribution of anal high-risk human papillomavirus (HR-HPV) among men who have sex with men living in the Central African Republic. *PLoS One* 2018; **13**(5): e0197845.
11. Grésenguet G, de Dieu Longo J, Tonen-Wolyec S, Bouassa R-SM, Belec L. Acceptability and usability evaluation of finger-stick whole blood HIV self-test as an HIV screening tool adapted to the general public in the Central African Republic. *The open AIDS journal* 2017; **11**: 101.
12. Longo JDD, Diemer HSC, Tonen-Wolyec S, et al. HIV self-testing in Central Africa: stakes and challenges. *Health sciences and disease* 2018; **19**(2).
13. Mbeko Simaleko M, Longo JDD, Magloire C-PS, et al. Persistent high-risk behavior and escalating HIV, syphilis and hepatitis B incidences among men who have sex with men living in Bangui, Central African Republic. *Pan African Medical Journal* 2018; **29**(1): 1-11.
14. Mbeko Simaleko M, Camengo Police SM, Longo JDD, Piette D, Humblet CP, Gresenguet G. Efficacité de la Combinaison d'Interventions de Prévention chez les Hommes Ayant des Rapports Sexuels avec des Hommes à Bangui (République Centrafricaine). *Health sciences and disease* 2020; **21**(7): 94-9.
15. Hessou SP, Glele-Ahanhanzo Y, Adekpedjou R, et al. HIV incidence and risk contributing factors among men who have sex with men in Benin: A prospective cohort study. *PloS one* 2020; **15**(6): e0233624.
16. Ahouada C, Diabaté S, Mondor M, et al. Acceptability of pre-exposure prophylaxis for HIV prevention: facilitators, barriers and impact on sexual risk behaviors among men who have sex with men in Benin. *BMC Public Health* 2020; **20**(1): 1267.
17. Goodman SH, Grosso AL, Ketende SC, et al. Examining the correlates of sexually transmitted infection testing among men who have sex with men in Ouagadougou and Bobo-Dioulasso, Burkina Faso. *Sexually transmitted diseases* 2016; **43**(5): 302-9.

18. Grosso AL, Ketende SC, Stahlman S, et al. Development and reliability of metrics to characterize types and sources of stigma among men who have sex with men and female sex workers in Togo and Burkina Faso. *BMC infectious diseases* 2019; **19**(1): 1-17.
19. Holland CE, Kouanda S, Lougué M, et al. Using Population-Size Estimation and Cross-sectional Survey Methods to Evaluate HIV Service Coverage Among Key Populations in Burkina Faso and Togo. *Public Health Rep* 2016; **131**(6): 773-82.
20. Kim H-Y, Grosso A, Ky-Zerbo O, et al. Stigma as a barrier to health care utilization among female sex workers and men who have sex with men in Burkina Faso. *Annals of epidemiology* 2018; **28**(1): 13-9.
21. Poteat T, Ackerman B, Diouf D, et al. HIV prevalence and behavioral and psychosocial factors among transgender women and cisgender men who have sex with men in 8 African countries: A cross-sectional analysis. *PLoS medicine* 2017; **14**(11): e1002422.
22. Stahlman S, Grosso A, Ketende S, et al. Suicidal ideation among MSM in three West African countries: associations with stigma and social capital. *International Journal of Social Psychiatry* 2016; **62**(6): 522-31.
23. Coulaud P-J, Sagaon-Teyssier L, Mimi M, et al. Changes in risky sexual behaviours among West African MSM enrolled in a quarterly HIV testing and counselling prevention programme (CohMSM ANRS 12324–Expertise France). *Sexually Transmitted Infections* 2020; **96**(2): 115-20.
24. Dah TTE, Yaya I, Mensah E, et al. Rapid antiretroviral therapy initiation and its effect on treatment response in MSM in West Africa. *Aids* 2021; **35**(13): 2201-10.
25. Dah TTE, Yaya I, Sagaon-Teyssier L, et al. Adherence to quarterly HIV prevention services and its impact on HIV incidence in men who have sex with men in West Africa (CohMSM ANRS 12324–Expertise France). *BMC Public Health* 2021; **21**(1): 1-13.
26. Laurent C, Keita BD, Yaya I, et al. HIV pre-exposure prophylaxis for men who have sex with men in west Africa: a multicountry demonstration study. *The Lancet HIV* 2021; **8**(7): e420-e8.
27. Yaya I, Boyer V, Ehlan PA, et al. Heterogeneity in the Prevalence of High-Risk Human Papillomavirus Infection in Human Immunodeficiency Virus-Negative and Human Immunodeficiency Virus-Positive Men Who Have Sex With Men in West Africa. *Clin Infect Dis* 2021; **73**(12): 2184-92.
28. Yaya I, Diallo F, Kouamé MJ, et al. Decrease in incidence of sexually transmitted infections symptoms in men who have sex with men enrolled in a quarterly HIV prevention and care programme in West Africa (CohMSM ANRS 12324-Expertise France). *Sex Transm Infect* 2022; **98**(2): 85-94.
29. Vuylsteke B, Semde G, Sika L, et al. High prevalence of HIV and sexually transmitted infections among male sex workers in Abidjan, Cote d'Ivoire: need for services tailored to their needs. *Sex Transm Infect* 2012; **88**(4): 288-93.
30. Aho J, Hakim A, Vuylsteke B, et al. Exploring risk behaviors and vulnerability for HIV among men who have sex with men in Abidjan, Cote d'Ivoire: poor knowledge, homophobia and sexual violence. *PLoS One* 2014; **9**(6): e99591.
31. Hakim AJ, Aho J, Semde G, et al. The epidemiology of HIV and prevention needs of men who have sex with men in Abidjan, Cote d'Ivoire. *PloS one* 2015; **10**(4): e0125218.
32. Couderc C, Keita BD, Anoma C, et al. Is PrEP needed for MSM in West Africa? HIV incidence in a prospective multicountry cohort. *JAIDS Journal of Acquired Immune Deficiency Syndromes* 2017; **75**(3): e80-e2.
33. Bouscaillou J, Evanno J, Prouté M, et al. Prevalence and risk factors associated with HIV and tuberculosis in people who use drugs in Abidjan, Ivory Coast. *Int J Drug Policy* 2016; **30**: 116-23.
34. Moran A, Scheim A, Lyons C, et al. Characterizing social cohesion and gender identity as risk determinants of HIV among cisgender men who have sex with men and transgender women in Côte d'Ivoire. *Ann Epidemiol* 2020; **42**: 25-32.
35. Ulanja MB, Lyons C, Ketende S, et al. The relationship between depression and sexual health service utilization among men who have sex with men (MSM) in Côte d'Ivoire, West Africa. *BMC Int Health Hum Rights* 2019; **19**(1): 11.

36. Diabaté S, Kra O, Biékoua YJ, et al. Pre-exposure prophylaxis among men who have sex with men in Côte d'Ivoire: a quantitative study of acceptability. *AIDS Care* 2021; **33**(9): 1228-36.
37. Inghels M, Kouassi AK, Niangoran S, et al. Telephone peer recruitment and interviewing during a respondent-driven sampling (RDS) survey: feasibility and field experience from the first phone-based RDS survey among men who have sex with men in Côte d'Ivoire. *BMC Med Res Methodol* 2021; **21**(1): 25.
38. Gyamerah AO, Taylor KD, Atuahene K, et al. Stigma, discrimination, violence, and HIV testing among men who have sex with men in four major cities in Ghana. *AIDS Care* 2020; **32**(8): 1036-44.
39. Kushwaha S, Lalani Y, Maina G, et al. "But the moment they find out that you are MSM…": a qualitative investigation of HIV prevention experiences among men who have sex with men (MSM) in Ghana's health care system. *BMC Public Health* 2017; **17**(1): 1-18.
40. Nelson LE, Wilton L, Agyarko-Poku T, et al. The association of HIV stigma and HIV/STD knowledge with sexual risk behaviors among adolescent and adult men who have sex with men in Ghana, West Africa. *Research in nursing & health* 2015; **38**(3): 194-206.
41. Girault P, Green K, Clement NF, Rahman YAA, Adams B, Wambugu S. Piloting a social networks strategy to increase HIV testing and counseling among men who have sex with men in Greater Accra and Ashanti Region, Ghana. *AIDS and Behavior* 2015; **19**(11): 1990-2000.
42. Abubakari GM, Nelson LE, Ogunbajo A, et al. Implementation and evaluation of a culturally grounded group-based HIV prevention programme for men who have sex with men in Ghana. *Glob Public Health* 2021; **16**(7): 1028-45.
43. Ogunbajo A, Kershaw T, Kushwaha S, Boakye F, Wallace-Atiapah ND, Nelson LE. Barriers, Motivators, and Facilitators to Engagement in HIV Care Among HIV-Infected Ghanaian Men Who have Sex with Men (MSM). *AIDS Behav* 2018; **22**(3): 829-39.
44. Gu LY, Zhang N, Mayer KH, et al. Autonomy-Supportive Healthcare Climate and HIV-Related Stigma Predict Linkage to HIV Care in Men Who Have Sex With Men in Ghana, West Africa. *J Int Assoc Provid AIDS Care* 2021; **20**: 2325958220978113.
45. Lieber M, Reynolds CW, Lieb W, McGill S, Beddoe AM. Human Papillomavirus Knowledge, Attitudes, Practices, and Prevalence Among Men Who Have Sex With Men in Monrovia, Liberia. *J Low Genit Tract Dis* 2018; **22**(4): 326-32.
46. Hakim A, Patnaik P, Telly N, et al. High Prevalence of Concurrent Male-Male Partnerships in the Context of Low Human Immunodeficiency Virus Testing Among Men Who Have Sex With Men in Bamako, Mali. *Sex Transm Dis* 2017; **44**(9): 565-70.
47. Hakim AJ, Coy K, Patnaik P, et al. An urgent need for HIV testing among men who have sex with men and transgender women in Bamako, Mali: Low awareness of HIV infection and viral suppression among those living with HIV. *PLoS One* 2018; **13**(11): e0207363.
48. Lahuerta M, Patnaik P, Ballo T, et al. HIV prevalence and related risk factors in men who have sex with men in Bamako, Mali: findings from a bio-behavioral survey using respondent-driven sampling. *AIDS and Behavior* 2018; **22**(7): 2079-88.
49. Knox J, Patnaik P, Hakim AJ, et al. Prevalence of condomless anal intercourse and associated risk factors among men who have sex with men in Bamako, Mali. *International journal of STD & AIDS* 2021; **32**(3): 218-27.
50. Koyalta D, Mboumba Bouassa RS, Maiga AI, et al. High Prevalence of Anal Oncogenic Human Papillomavirus Infection in Young Men Who Have Sex with Men Living in Bamako, Mali. *Infect Agent Cancer* 2021; **16**(1): 51.
51. Adam PC, de Wit JB, Toskin I, et al. Estimating levels of HIV testing, HIV prevention coverage, HIV knowledge, and condom use among men who have sex with men (MSM) in low-income and middle-income countries. *J Acquir Immune Defic Syndr* 2009; **52 Suppl 2**: S143-51.
52. Merrigan M, Azeez A, Afolabi B, et al. HIV prevalence and risk behaviours among men having sex with men in Nigeria. *Sexually transmitted infections* 2011; **87**(1): 65-70.

53. Strömdahl S, Onigbanjo Williams A, Eziefule B, et al. Associations of consistent condom use among men who have sex with men in Abuja, Nigeria. *AIDS research and human retroviruses* 2012; **28**(12): 1756-62.
54. Strömdahl S, Onigbanjo Williams A, Eziefule B, et al. An assessment of stigma and human right violations among men who have sex with men in Abuja, Nigeria. *BMC International Health and Human Rights* 2019; **19**(1): 1-7.
55. Eluwa GI, Sylvia A, Luchters S, Ahonsi B. HIV risk perception and risk behaviors among men who have sex with men in Nigeria. *Journal of AIDS & Clinical Research* 2015; **6**(7): 1.
56. Eluwa GI, Adebajo SB, Eluwa T, Ogbanufe O, Ilesanmi O, Nzelu C. Rising HIV prevalence among men who have sex with men in Nigeria: a trend analysis. *BMC Public Health* 2019; **19**(1): 1-10.
57. Adebajo S, Obianwu O, Eluwa G, et al. Comparison of audio computer assisted self-interview and face-to-face interview methods in eliciting HIV-related risks among men who have sex with men and men who inject drugs in Nigeria. *PloS one* 2014; **9**(1): e81981.
58. Sheehy M, Tun W, Vu L, Adebajo S, Obianwu O, Karlyn A. High levels of bisexual behavior and factors associated with bisexual behavior among men having sex with men (MSM) in Nigeria. *Aids Care* 2014; **26**(1): 116-22.
59. Vu L, Adebajo S, Tun W, et al. High HIV prevalence among men who have sex with men in Nigeria: implications for combination prevention. *JAIDS Journal of Acquired Immune Deficiency Syndromes* 2013; **63**(2): 221-7.
60. Vu L, Andrinopoulos K, Tun W, Adebajo S. High levels of unprotected anal intercourse and never testing for HIV among men who have sex with men in Nigeria: evidence from a cross-sectional survey for the need for innovative approaches to HIV prevention. *Sexually transmitted infections* 2013; **89**(8): 659-65.
61. Baral SD, Ketende S, Schwartz S, et al. Evaluating respondent-driven sampling as an implementation tool for universal coverage of antiretroviral studies among men who have sex with men living with HIV. *J Acquir Immune Defic Syndr* 2015; **68 Suppl 2**(0 2): S107-13.
62. Billings E, Kijak GH, Sanders-Buell E, et al. New Subtype B Containing HIV-1 Circulating Recombinant of sub-Saharan Africa Origin in Nigerian Men Who Have Sex With Men. *J Acquir Immune Defic Syndr* 2019; **81**(5): 578-84.
63. Charurat M, Emmanuel B, Akolo C, et al. Uptake of treatment as prevention for HIV and continuum of care among HIV-positive men who have sex with men in Nigeria. *Journal of acquired immune deficiency syndromes (1999)* 2015; **68**(Suppl 2): S114.
64. Crowell TA, Keshinro B, Baral SD, et al. Stigma, access to healthcare, and HIV risks among men who sell sex to men in Nigeria. *Journal of the International AIDS Society* 2017; **20**(1): 21489.
65. Crowell TA, Baral SD, Schwartz S, et al. Time to change the paradigm: limited condom and lubricant use among Nigerian men who have sex with men and transgender women despite availability and counseling. *Annals of epidemiology* 2019; **31**: 11-9. e3.
66. Kayode BO, Mitchell A, Ndembi N, et al. Retention of a cohort of men who have sex with men and transgender women at risk for and living with HIV in Abuja and Lagos, Nigeria: a longitudinal analysis. *Journal of the International AIDS Society* 2020; **23**: e25592.
67. Li Y, Liu H, Ramadhani HO, et al. Genetic clustering analysis for HIV infection among MSM in Nigeria: implications for intervention. *Aids* 2020; **34**(2): 227-36.
68. Nowak RG, Gravitt PE, He X, et al. Prevalence of anal high-risk human papillomavirus infections among HIV-positive and HIV-negative men who have sex with men (MSM) in Nigeria. *Sexually transmitted diseases* 2016; **43**(4): 243.
69. Nowak RG, Bentzen SM, Ravel J, et al. Rectal microbiota among HIV-uninfected, untreated HIV, and treated HIV-infected men who have sex with men (MSM) in Nigeria. *AIDS (London, England)* 2017; **31**(6): 857.
70. Nowak RG, Mitchell A, Crowell TA, et al. Individual and sexual network predictors of HIV incidence among men who have sex with men in Nigeria. *Journal of acquired immune deficiency syndromes (1999)* 2019; **80**(4): 444.

71. Nowak RG, Bentzen SM, Ravel J, et al. Anal microbial patterns and oncogenic human papillomavirus in a pilot study of Nigerian men who have sex with men at risk for or living with HIV. *AIDS research and human retroviruses* 2019; **35**(3): 267-75.
72. Nowak RG, Nnaji CH, Dauda W, et al. Satisfaction with high-resolution anoscopy for anal cancer screening among men who have sex with men: a cross-sectional survey in Abuja, Nigeria. *BMC cancer* 2020; **20**(1): 1-9.
73. Olawore O, Crowell TA, Ketende SC, et al. Individual and partnership characteristics associated with consistent condom use in a cohort of cisgender men who have sex with men and transgender women in Nigeria. *BMC public health* 2021; **21**(1): 1-17.
74. Ramadhani HO, Liu H, Nowak RG, et al. Sexual partner characteristics and incident rectal Neisseria gonorrhoeae and Chlamydia trachomatis infections among gay men and other men who have sex with men (MSM): a prospective cohort in Abuja and Lagos, Nigeria. *Sexually transmitted infections* 2017; **93**(5): 348-55.
75. Ramadhani HO, Ndembi N, Nowak RG, et al. Individual and network factors associated with HIV care continuum outcomes among Nigerian MSM accessing healthcare services. *Journal of acquired immune deficiency syndromes (1999)* 2018; **79**(1): e7.
76. Ramadhani HO, Crowell TA, Nowak RG, et al. Association of age with healthcare needs and engagement among Nigerian men who have sex with men and transgender women: cross-sectional and longitudinal analyses from an observational cohort. *Journal of the International AIDS Society* 2020; **23**: e25599.
77. Robbins SJ, Dauda W, Kokogho A, et al. Oral sex practices among men who have sex with men and transgender women at risk for and living with HIV in Nigeria. *PloS one* 2020; **15**(9): e0238745.
78. Rodriguez-Hart C, Liu H, Nowak RG, et al. Serosorting and sexual risk for HIV infection at the ego-alter dyadic level: an egocentric sexual network study among MSM in Nigeria. *AIDS and Behavior* 2016; **20**(11): 2762-71.
79. Rodriguez-Hart C, Bradley C, German D, et al. The synergistic impact of sexual stigma and psychosocial well-being on HIV testing: a mixed-methods study among Nigerian men who have sex with men. *AIDS and Behavior* 2018; **22**(12): 3905-15.
80. Schwartz SR, Nowak RG, Orazulike I, et al. The immediate effect of the Same-Sex Marriage Prohibition Act on stigma, discrimination, and engagement on HIV prevention and treatment services in men who have sex with men in Nigeria: analysis of prospective data from the TRUST cohort. *The lancet HIV* 2015; **2**(7): e299-e306.
81. Stahlman S, Nowak RG, Liu H, et al. Online sex-seeking among men who have sex with men in Nigeria: implications for online intervention. *AIDS and Behavior* 2017; **21**(11): 3068-77.
82. Tiamiyu AB, Lawlor J, Hu F, et al. HIV status disclosure by Nigerian men who have sex with men and transgender women living with HIV: a cross-sectional analysis at enrollment into an observational cohort. *BMC Public Health* 2020; **20**(1): 1282.
83. Tobin-West C, Nwajagu S, Maduka O, Oranu E, Onyekwere V, Tamuno I. Exploring the HIV-risk practices of men who have sex with men in Port Harcourt city, Nigeria. *Annals of Tropical Medicine and Public Health* 2017; **10**(3).
84. Offie DC, Obeagu EI, Akueshi C, et al. Facilitators and Barriers to Retention in HIV Care among HIV Infected MSM Attending Community Health Center Yaba, Lagos Nigeria. *Journal of Pharmaceutical Research International* 2021.
85. Tun W, Vu L, Dirisu O, et al. Uptake of HIV self-testing and linkage to treatment among men who have sex with men (MSM) in Nigeria: A pilot programme using key opinion leaders to reach MSM. *J Int AIDS Soc* 2018; **21 Suppl 5**(Suppl Suppl 5): e25124.
86. Ibiloye O, Decroo T, Eyona N, Eze P, Agada P. Characteristics and early clinical outcomes of key populations attending comprehensive community-based HIV care: Experiences from Nasarawa State, Nigeria. *PLoS One* 2018; **13**(12): e0209477.

87. Ibiloye O, Jwanle P, Masquillier C, et al. Long-term retention and predictors of attrition for key populations receiving antiretroviral treatment through community-based ART in Benue State Nigeria: A retrospective cohort study. *PLoS One* 2021; **16**(11): e0260557.
88. Ibiloye O, Akande P, Plang J, et al. Community health worker-led ART delivery improved scheduled antiretroviral drug refill among men who have sex with men in Lagos State, Nigeria. *Int Health* 2021; **13**(2): 196-8.
89. Afolaranmi TO, Hassan ZI, Ugwu OJ, et al. Retention in HIV care and its predictors among HIV-infected men who have sex with men in Plateau state, North Central Nigeria. *J Family Med Prim Care* 2021; **10**(4): 1596-601.
90. Ndiaye HD, Tchiakpe E, Vidal N, et al. HIV type 1 subtype C remains the predominant subtype in men having sex with men in Senegal. *AIDS Res Hum Retroviruses* 2013; **29**(9): 1265-72.
91. Wade AS, Kane CT, Diallo PAN, et al. HIV infection and sexually transmitted infections among men who have sex with men in Senegal. *Aids* 2005; **19**(18): 2133-40.
92. Dramé FM, Crawford EE, Diouf D, Beyrer C, Baral SD. A pilot cohort study to assess the feasibility of HIV prevention science research among men who have sex with men in Dakar, Senegal. *Journal of the International AIDS Society* 2013; **16**: 18753.
93. Lyons CE, Ketende S, Diouf D, et al. Potential Impact of Integrated Stigma Mitigation Interventions in Improving HIV/AIDS Service Delivery and Uptake for Key Populations in Senegal. *J Acquir Immune Defic Syndr* 2017; **74 Suppl 1**(Suppl 1): S52-s9.
94. Lyons CE, Olawore O, Turpin G, et al. Intersectional stigmas and HIV-related outcomes among a cohort of key populations enrolled in stigma mitigation interventions in Senegal. *AIDS (London, England)* 2020; **34**(Suppl 1): S63.
95. Mason K, Ketende S, Peitzmeier S, et al. A cross-sectional analysis of population demographics, HIV knowledge and risk behaviors, and prevalence and associations of HIV among men who have sex with men in the Gambia. *AIDS Res Hum Retroviruses* 2013; **29**(12): 1547-52.
96. Ekouevi DK, Dagnra CY, Goilibe KB, et al. HIV seroprevalence and associated factors among men who have sex with men in Togo. *Rev Epidemiol Sante Publique* 2014; **62**(2): 127-34.
97. Bakai TA, Ekouevi DK, Tchounga BK, et al. Condom use and associated factors among men who have sex with men in Togo, West Africa. *Pan African Medical Journal* 2016; **23**(1).
98. Ruiseñor-Escudero H, Grosso A, Ketende S, et al. Using a social ecological framework to characterize the correlates of HIV among men who have sex with men in Lomé, Togo. *AIDS care* 2017; **29**(9): 1169-77.
99. Ruiseñor-Escudero H, Lyons C, Ketende S, et al. Prevalence and factors associated to disclosure of same-sex practices to family members and health care workers among men who have sex with men in Togo. *AIDS care* 2019.
100. Ruiseñor-Escudero H, Lyons C, Ketende S, et al. Consistent condom use among men who have sex with men in Lome and Kara, Togo. *AIDS research and human retroviruses* 2019; **35**(6): 519-28.
101. Teclessou JN, Akakpo SA, Ekouevi KD, Koumagnanou G, Singo-Tokofai A, Pitche PV. Evolution of HIV prevalence and behavioral factors among MSM in Togo between 2011 and 2015. *Pan Afr Med J* 2017; **28**: 191.
102. Sadio AJ, Gbeasor-Komlanvi FA, Konu YR, et al. Prevalence of HIV infection and hepatitis B and factors associated with them among men who had sex with men in Togo in 2017. *Med Sante Trop* 2019; **29**(3): 294-301.
103. Coulaud P-j, Mujimbere G, Nitunga A, et al. An assessment of health interventions required to prevent the transmission of HIV infection among men having sex with men in Bujumbura, Burundi. *Journal of community health* 2016; **41**(5): 1033-43.
104. Lillie T, Boyee D, Kamariza G, Nkunzimana A, Gashobotse D, Persaud N. Increasing Testing Options for Key Populations in Burundi Through Peer-Assisted HIV Self-Testing: Descriptive Analysis of Routine Programmatic Data. *JMIR Public Health Surveill* 2021; **7**(9): e24272.

105. Gebrebrhan H, Kambaran C, Sivro A, et al. Rectal microbiota diversity in Kenyan MSM is inversely associated with frequency of receptive anal sex, independent of HIV status. *Aids* 2021; **35**(7): 1091-101.
106. Sanders EJ, Graham SM, Okuku HS, et al. HIV-1 infection in high risk men who have sex with men in Mombasa, Kenya. *Aids* 2007; **21**(18): 2513-20.
107. Kamali A, Price MA, Lakhi S, et al. Creating an African HIV clinical research and prevention trials network: HIV prevalence, incidence and transmission. *PloS one* 2015; **10**(1): e0116100.
108. Price MA, Rida W, Mwangome M, et al. Identifying at-risk populations in Kenya and South Africa: HIV incidence in cohorts of men who report sex with men, sex workers, and youth. *JAIDS Journal of Acquired Immune Deficiency Syndromes* 2012; **59**(2): 185-93.
109. Luchters S, Geibel S, Syengo M, et al. Use of AUDIT, and measures of drinking frequency and patterns to detect associations between alcohol and sexual behaviour in male sex workers in Kenya. *BMC public health* 2011; **11**(1): 1-8.
110. Graham SM, Mugo P, Gichuru E, et al. Adherence to antiretroviral therapy and clinical outcomes among young adults reporting high-risk sexual behavior, including men who have sex with men, in coastal Kenya. *AIDS Behav* 2013; **17**(4): 1255-65.
111. Mdodo R, Gust D, Otieno FO, et al. Investigation of HIV incidence rates in a high-risk, high-prevalence Kenyan population: potential lessons for intervention trials and programmatic strategies. *Journal of the International Association of Providers of AIDS Care (JIAPAC)* 2016; **15**(1): 42-50.
112. Muraguri N, Tun W, Okal J, et al. HIV and STI prevalence and risk factors among male sex workers and other men who have sex with men in Nairobi, Kenya. *Journal of acquired immune deficiency syndromes (1999)* 2015; **68**(1): 91.
113. McKinnon LR, Gakii G, Juno JA, et al. High HIV risk in a cohort of male sex workers from Nairobi, Kenya. *Sexually transmitted infections* 2014; **90**(3): 237-42.
114. Möller LM, Stolte IG, Geskus RB, et al. Changes in sexual risk behavior among MSM participating in a research cohort in coastal Kenya. *AIDS (London, England)* 2015; **29**(0 3): S211.
115. Sanders EJ, Okuku HS, Smith AD, et al. High HIV-1 incidence, correlates of HIV-1 acquisition, and high viral loads following seroconversion among men who have sex with men in Coastal Kenya. *AIDS (London, England)* 2013; **27**(3): 437.
116. Wahome E, Thiong'o AN, Mwashigadi G, et al. An empiric risk score to guide PrEP targeting among MSM in coastal Kenya. *AIDS and Behavior* 2018; **22**(1): 35-44.
117. Wahome EW, Graham SM, Thiong'o AN, et al. PrEP uptake and adherence in relation to HIV-1 incidence among Kenyan men who have sex with men. *EClinicalMedicine* 2020; **26**: 100541.
118. Githuka G, Hladik W, Mwalili S, et al. Populations at increased risk for HIV infection in Kenya: results from a national population-based household survey, 2012. *J Acquir Immune Defic Syndr* 2014; **66 Suppl 1**(Suppl 1): S46-56.
119. Shangani S, Naanyu V, Mwangi A, et al. Factors associated with HIV testing among men who have sex with men in Western Kenya: a cross-sectional study. *Int J STD AIDS* 2017; **28**(2): 179-87.
120. Bhattacharjee P, McClarty LM, Musyoki H, et al. Monitoring HIV prevention programme outcomes among key populations in Kenya: findings from a national survey. *PLoS One* 2015; **10**(8): e0137007.
121. Musyoki H, Bhattacharjee P, Blanchard AK, et al. Changes in HIV prevention programme outcomes among key populations in Kenya: data from periodic surveys. *PLoS One* 2018; **13**(9): e0203784.
122. Nyblade L, Reddy A, Mbote D, et al. The relationship between health worker stigma and uptake of HIV counseling and testing and utilization of non-HIV health services: the experience of male and female sex workers in Kenya. *AIDS Care* 2017; **29**(11): 1364-72.
123. Kimani M, van der Elst EM, Chiro O, et al. Pr EP interest and HIV-1 incidence among MSM and transgender women in coastal Kenya. *Journal of the International AIDS Society* 2019; **22**(6): e25323.

124. Korhonen C, Kimani M, Wahome E, et al. Depressive symptoms and problematic alcohol and other substance use in 1476 gay, bisexual, and other MSM at three research sites in Kenya. *Aids* 2018; **32**(11): 1507-15.
125. Kunzweiler CP, Bailey RC, Okall DO, Graham SM, Mehta SD, Otieno FO. Factors Associated With Prevalent HIV Infection Among Kenyan MSM: The Anza Mapema Study. *J Acquir Immune Defic Syndr* 2017; **76**(3): 241-9.
126. Kunzweiler CP, Bailey RC, Mehta SD, et al. Factors associated with viral suppression among HIV-positive Kenyan gay and bisexual men who have sex with men. *AIDS Care* 2018; **30**(sup5): S76-s88.
127. Kunzweiler CP, Bailey RC, Okall DO, Graham SM, Mehta SD, Otieno FO. Depressive symptoms, alcohol and drug use, and physical and sexual abuse among men who have sex with men in Kisumu, Kenya: the Anza Mapema Study. *AIDS and Behavior* 2018; **22**(5): 1517-29.
128. Fogel JM, Sandfort T, Zhang Y, et al. Accuracy of Self-Reported HIV Status Among African Men and Transgender Women Who Have Sex with Men Who were Screened for Participation in a Research Study: HPTN 075. *AIDS and Behavior* 2019; **23**(1): 289-94.
129. Palumbo PJ, Zhang Y, Clarke W, et al. Uptake of antiretroviral treatment and viral suppression among men who have sex with men and transgender women in sub-Saharan Africa in an observational cohort study: HPTN 075. *Int J Infect Dis* 2021; **104**: 465-70.
130. Sandfort TGM, Dominguez K, Kayange N, et al. HIV testing and the HIV care continuum among sub-Saharan African men who have sex with men and transgender women screened for participation in HPTN 075. *PLoS One* 2019; **14**(5): e0217501.
131. Sandfort TG, Mbilizi Y, Sanders EJ, et al. HIV incidence in a multinational cohort of men and transgender women who have sex with men in sub-Saharan Africa: Findings from HPTN 075. *PloS one* 2021; **16**(2): e0247195.
132. Sivay MV, Palumbo PJ, Zhang Y, et al. Human Immunodeficiency Virus (HIV) Drug Resistance, Phylogenetic Analysis, and Superinfection Among Men Who Have Sex with Men and Transgender Women in Sub-Saharan Africa: HIV Prevention Trials Network (HPTN) 075 Study. *Clinical Infectious Diseases* 2021; **73**(1): 60-7.
133. Zhang Y, Fogel JM, Guo X, et al. Antiretroviral drug use and HIV drug resistance among MSM and transgender women in sub-Saharan Africa. *AIDS (London, England)* 2018; **32**(10): 1301.
134. Fearon E, Bourne A, Tenza S, et al. Online socializing among men who have sex with men and transgender people in Nairobi and Johannesburg and implications for public health-related research and health promotion: an analysis of qualitative and respondent-driven sampling survey data. *J Int AIDS Soc* 2020; **23 Suppl 6**(Suppl 6): e25603.
135. Smith AD, Fearon E, Kabuti R, et al. Disparities in HIV/STI burden and care coverage among men and transgender persons who have sex with men in Nairobi, Kenya: a cross-sectional study. *BMJ Open* 2021; **11**(12): e055783.
136. Smith AD, Kimani J, Kabuti R, Weatherburn P, Fearon E, Bourne A. HIV burden and correlates of infection among transfeminine people and cisgender men who have sex with men in Nairobi, Kenya: an observational study. *Lancet HIV* 2021; **8**(5): e274-e83.
137. Bhattacharjee P, Isac S, Musyoki H, et al. HIV prevalence, testing and treatment among men who have sex with men through engagement in virtual sexual networks in Kenya: a cross-sectional bio-behavioural study. *J Int AIDS Soc* 2020; **23 Suppl 2**(Suppl 2): e25516.
138. Dijkstra M, Mohamed K, Kigoro A, et al. Peer Mobilization and Human Immunodeficiency Virus (HIV) Partner Notification Services Among Gay, Bisexual, and Other Men Who Have Sex With Men and Transgender Women in Coastal Kenya Identified a High Number of Undiagnosed HIV Infections. *Open Forum Infect Dis* 2021; **8**(6): ofab219.
139. Virkud AV, Arimi P, Ssengooba F, et al. Access to HIV prevention services in East African cross-border areas: a 2016-2017 cross-sectional bio-behavioural study. *J Int AIDS Soc* 2020; **23 Suppl 3**(Suppl 3): e25523.

140. Ntata PRT, Muula AS, Siziya S. Socio-demographic characteristics and sexual health related attitudes and practices of men having sex with men in central and southern Malawi. *Tanzania Journal of Health Research* 2008; **10**(3): 124-30.
141. Baral S, Trapence G, Motimedi F, et al. HIV prevalence, risks for HIV infection, and human rights among men who have sex with men (MSM) in Malawi, Namibia, and Botswana. *PLoS One* 2009; **4**(3): e4997.
142. Beyrer C, Trapence G, Motimedi F, et al. Bisexual concurrency, bisexual partnerships, and HIV among Southern African men who have sex with men. *Sexually transmitted infections* 2010; **86**(4): 323-7.
143. Fay H, Baral SD, Trapence G, et al. Stigma, health care access, and HIV knowledge among men who have sex with men in Malawi, Namibia, and Botswana. *AIDS Behav* 2011; **15**(6): 1088-97.
144. Stahlman S, Johnston LG, Yah C, et al. Respondent-driven sampling as a recruitment method for men who have sex with men in southern sub-Saharan Africa: a cross-sectional analysis by wave. *Sex Transm Infect* 2016; **92**(4): 292-8.
145. Wirtz AL, Jumbe V, Trapence G, et al. HIV among men who have sex with men in Malawi: elucidating HIV prevalence and correlates of infection to inform HIV prevention. *Journal of the International AIDS Society* 2013; **16**: 18742.
146. Wirtz AL, Trapence G, Jumbe V, et al. Feasibility of a combination HIV prevention program for men who have sex with men in Blantyre, Malawi. *Journal of acquired immune deficiency syndromes (1999)* 2015; **70**(2): 155.
147. Wirtz AL, Trapence G, Kamba D, et al. Geographical disparities in HIV prevalence and care among men who have sex with men in Malawi: results from a multisite cross-sectional survey. *The Lancet HIV* 2017; **4**(6): e260-e9.
148. Rucinski K, Masankha Banda L, Olawore O, et al. HIV Testing Approaches to Optimize Prevention and Treatment for Key and Priority Populations in Malawi. *Open Forum Infect Dis* 2022; **9**(4): ofac038.
149. Boothe MAS, Sathane I, Baltazar CS, et al. Low engagement in HIV services and progress through the treatment cascade among key populations living with HIV in Mozambique: alarming gaps in knowledge of status. *BMC Public Health* 2021; **21**(1): 146.
150. Boothe MAS, Semá Baltazar C, Sathane I, et al. Young key populations left behind: The necessity for a targeted response in Mozambique. *PLoS One* 2021; **16**(12): e0261943.
151. Horth RZ, Cummings B, Young PW, et al. Correlates of HIV Testing Among Men Who have Sex with Men in Three Urban Areas of Mozambique: Missed Opportunities for Prevention. *AIDS Behav* 2015; **19**(11): 1978-89.
152. Sathane I, Horth R, Young P, et al. Risk factors associated with HIV among men who have sex only with men and men who have sex with both men and women in three urban areas in Mozambique. *AIDS and Behavior* 2016; **20**(10): 2296-308.
153. Chapman J, Koleros A, Delmont Y, Pegurri E, Gahire R, Binagwaho A. High HIV risk behavior among men who have sex with men in Kigali, Rwanda: making the case for supportive prevention policy. *AIDS Care* 2011; **23**(4): 449-55.
154. Ntale RS, Rutayisire G, Mujyarugamba P, et al. HIV seroprevalence, self-reported STIs and associated risk factors among men who have sex with men: a cross-sectional study in Rwanda, 2015. *Sex Transm Infect* 2019; **95**(1): 71-4.
155. Twahirwa Rwema JO, Lyons CE, Herbst S, et al. HIV infection and engagement in HIV care cascade among men who have sex with men and transgender women in Kigali, Rwanda: a cross-sectional study. *J Int AIDS Soc* 2020; **23 Suppl 6**(Suppl 6): e25604.
156. Magesa DJ, Mtui LJ, Abdul M, et al. Barriers to men who have sex with men attending HIV related health services in Dar es Salaam, Tanzania. *Tanzan J Health Res* 2014; **16**(2): 118-26.
157. Dahoma M, Johnston LG, Holman A, et al. HIV and related risk behavior among men who have sex with men in Zanzibar, Tanzania: results of a behavioral surveillance survey. *AIDS and Behavior* 2011; **15**(1): 186-92.

158. Johnston LG, Holman A, Dahoma M, et al. HIV risk and the overlap of injecting drug use and high-risk sexual behaviours among men who have sex with men in Zanzibar (Unguja), Tanzania. *International Journal of Drug Policy* 2010; **21**(6): 485-92.
159. Khatib A, Haji S, Khamis M, et al. Reproducibility of Respondent-Driven Sampling (RDS) in Repeat Surveys of Men Who have Sex with Men, Unguja, Zanzibar. *AIDS Behav* 2017; **21**(7): 2180-7.
160. Nyoni J, Ross MW. Factors associated with HIV testing in men who have sex with men, in Dar es Salaam, Tanzania. *Sexually transmitted infections* 2012.
161. Nyoni JE, Ross MW. Condom use and HIV-related behaviors in urban Tanzanian men who have sex with men: a study of beliefs, HIV knowledge sources, partner interactions and risk behaviors. *AIDS Care* 2013; **25**(2): 223-9.
162. Mmbaga EJ, Dodo MJ, Leyna GH, Moen K, Leshabari MT. Sexual practices and perceived susceptibility to HIV infection among men who have sex with men in Dar Es Salaam, mainland Tanzania. *Journal of AIDS and Clinical Research* 2012; **2012**: 1-6.
163. Ahaneku H, Ross MW, Nyoni JE, et al. Depression and HIV risk among men who have sex with men in Tanzania. *AIDS care* 2016; **28**(sup1): 140-7.
164. Anderson AM, Ross MW, Nyoni JE, McCurdy SA. High prevalence of stigma-related abuse among a sample of men who have sex with men in Tanzania: implications for HIV prevention. *AIDS care* 2015; **27**(1): 63-70.
165. Romijnders KA, Nyoni JE, Ross MW, et al. Lubricant use and condom use during anal sex in men who have sex with men in Tanzania. *Int J STD AIDS* 2016; **27**(14): 1289-302.
166. Ross MW, Nyoni J, Ahaneku HO, Mbwambo J, McClelland RS, McCurdy SA. High HIV seroprevalence, rectal STIs and risky sexual behaviour in men who have sex with men in Dar es Salaam and Tanga, Tanzania. *BMJ Open* 2014; **4**(8): e006175.
167. Mmbaga EJ, Moen K, Leyna GH, Mpembeni R, Leshabari MT. HIV Prevalence and Associated Risk Factors Among Men Who Have Sex With Men in Dar es Salaam, Tanzania. *J Acquir Immune Defic Syndr* 2018; **77**(3): 243-9.
168. Ross MW, Kashiha J, Nyoni J, Larsson M, Agardh A. Electronic Media Access and Use for Sexuality and Sexual Health Education Among Men Who Have Sex With Men in Four Cities in Tanzania. *International Journal of Sexual Health* 2018; **30**(3): 264-70.
169. Kajubi P, Kamya MR, Raymond HF, et al. Gay and bisexual men in Kampala, Uganda. *AIDS Behav* 2008; **12**(3): 492-504.
170. Raymond HF, Kajubi P, Kamya MR, Rutherford GW, Mandel JS, McFarland W. Correlates of unprotected receptive anal intercourse among gay and bisexual men: Kampala, Uganda. *AIDS Behav* 2009; **13**(4): 677-81.
171. Hladik W, Barker J, Ssenkusu JM, et al. HIV infection among men who have sex with men in Kampala, Uganda--a respondent driven sampling survey. *PLoS One* 2012; **7**(5): e38143.
172. Robb ML, Eller LA, Kibuuka H, et al. Prospective study of acute HIV-1 infection in adults in East Africa and Thailand. *New England Journal of Medicine* 2016; **374**(22): 2120-30.
173. Wanyenze RK, Musinguzi G, Matovu JK, et al. "If You Tell People That You Had Sex with a Fellow Man, It Is Hard to Be Helped and Treated": Barriers and Opportunities for Increasing Access to HIV Services among Men Who Have Sex with Men in Uganda. *PLoS One* 2016; **11**(1): e0147714.
174. Hladik W, Sande E, Berry M, et al. Men Who Have Sex with Men in Kampala, Uganda: Results from a Bio-Behavioral Respondent Driven Sampling Survey. *AIDS Behav* 2017; **21**(5): 1478-90.
175. Okoboi S, Lazarus O, Castelnuovo B, et al. Peer distribution of HIV self-test kits to men who have sex with men to identify undiagnosed HIV infection in Uganda: A pilot study. *PLoS One* 2020; **15**(1): e0227741.
176. Okoboi S, Castelnuovo B, Van Geertruyden JP, et al. Cost-Effectiveness of Peer-Delivered HIV Self-Tests for MSM in Uganda. *Front Public Health* 2021; **9**: 651325.
177. Harris TG, Wu Y, Parmley LE, et al. HIV care cascade and associated factors among men who have sex with men, transgender women, and genderqueer individuals in Zimbabwe: findings from a biobehavioural survey using respondent-driven sampling. *The Lancet HIV* 2022; **9**(3): e182-e201.

178. Parmley LE, Chingombe I, Wu Y, et al. High Burden of Active Syphilis and Human Immunodeficiency Virus/Syphilis Coinfection Among Men Who Have Sex With Men, Transwomen, and Genderqueer Individuals in Zimbabwe. *Sex Transm Dis* 2022; **49**(2): 111-6.
179. Parmley LE, Harris TG, Chingombe I, et al. Engagement in the pre-exposure prophylaxis (PrEP) cascade among a respondent-driven sample of sexually active men who have sex with men and transgender women during early PrEP implementation in Zimbabwe. *J Int AIDS Soc* 2022; **25**(2): e25873.
180. Baral SD, Ketende S, Mnisi Z, et al. A cross-sectional assessment of the burden of HIV and associated individual-and structural-level characteristics among men who have sex with men in Swaziland. *Journal of the International AIDS Society* 2013; **16**: 18768.
181. Brown CA, Grosso AL, Adams D, et al. Characterizing the individual, social, and structural determinants of condom use among men who have sex with men in Swaziland. *AIDS research and human retroviruses* 2016; **32**(6): 539-46.
182. Grover E, Grosso A, Ketende S, et al. Social cohesion, social participation and HIV testing among men who have sex with men in Swaziland. *AIDS care* 2016; **28**(6): 795-804.
183. Risher K, Adams D, Sithole B, et al. Sexual stigma and discrimination as barriers to seeking appropriate healthcare among men who have sex with men in Swaziland. *Journal of the International AIDS Society* 2013; **16**: 18715.
184. Stahlman S, Grosso A, Ketende S, et al. Characteristics of men who have sex with men in southern Africa who seek sex online: a cross-sectional study. *Journal of medical Internet research* 2015; **17**(5): e4230.
185. Baral S, Adams D, Lebona J, et al. A cross-sectional assessment of population demographics, HIV risks and human rights contexts among men who have sex with men in Lesotho. *J Int AIDS Soc* 2011; **14**: 36.
186. Stahlman S, Grosso A, Ketende S, et al. Depression and social stigma among MSM in Lesotho: implications for HIV and sexually transmitted infection prevention. *AIDS and Behavior* 2015; **19**(8): 1460-9.
187. Wendi D, Stahlman S, Grosso A, et al. Depressive symptoms and substance use as mediators of stigma affecting men who have sex with men in Lesotho: a structural equation modeling approach. *Annals of epidemiology* 2016; **26**(8): 551-6.
188. Russell C, Tahlil K, Davis M, et al. Barriers to condom use among key populations in Namibia. *Int J STD AIDS* 2019; **30**(14): 1417-24.
189. Cloete A, Simbayi LC, Kalichman SC, Strebel A, Henda N. Stigma and discrimination experiences of HIV-positive men who have sex with men in Cape Town, South Africa. *AIDS Care* 2008; **20**(9): 1105-10.
190. Jobson G, Tucker A, de Swardt G, et al. Gender identity and HIV risk among men who have sex with men in Cape Town, South Africa. *AIDS Care* 2018; **30**(11): 1421-5.
191. Lane T, Shade SB, McIntyre J, Morin SF. Alcohol and sexual risk behavior among men who have sex with men in South African township communities. *AIDS and Behavior* 2008; **12**(1): 78-85.
192. Nel JA, Yi H, Sandfort TG, Rich E. HIV-untested men who have sex with men in South Africa: the perception of not being at risk and fear of being tested. *AIDS and Behavior* 2013; **17**(1): 51-9.
193. Sandfort TG, Nel J, Rich E, Reddy V, Yi H. HIV testing and self-reported HIV status in South African men who have sex with men: results from a community-based survey. *Sexually transmitted infections* 2008; **84**(6): 425-9.
194. Burrell E, Mark D, Grant R, Wood R, Bekker LG. Sexual risk behaviours and HIV-1 prevalence among urban men who have sex with men in Cape Town, South Africa. *Sex Health* 2010; **7**(2): 149-53.
195. Knox J, Sandfort T, Yi H, Reddy V, Maimane S. Social vulnerability and HIV testing among South African men who have sex with men. *International journal of STD & AIDS* 2011; **22**(12): 709-13.
196. Knox J, Reddy V, Kaighobadi F, Nel D, Sandfort T. Communicating HIV status in sexual interactions: assessing social cognitive constructs, situational factors, and individual characteristics among South African MSM. *AIDS and Behavior* 2013; **17**(1): 350-9.

197. Arnold MP, Struthers H, McIntyre J, Lane T. Contextual correlates of per partner unprotected anal intercourse rates among MSM in Soweto, South Africa. *AIDS and Behavior* 2013; **17**(1): 4-11.
198. Lane T, Raymond HF, Dladla S, et al. High HIV prevalence among men who have sex with men in Soweto, South Africa: results from the Soweto Men's Study. *AIDS and Behavior* 2011; **15**(3): 626-34.
199. Baral S, Burrell E, Scheibe A, Brown B, Beyrer C, Bekker LG. HIV risk and associations of HIV infection among men who have sex with men in peri-urban Cape Town, South Africa. *BMC Public Health* 2011; **11**: 766.
200. Buchbinder SP, Glidden DV, Liu AY, et al. Who should be offered HIV pre-exposure prophylaxis (PrEP)?: A secondary analysis of a Phase 3 PrEP efficacy trial in men who have sex with men and transgender women. *The Lancet infectious diseases* 2014; **14**(6): 468.
201. Buchbinder SP, Glidden DV, Liu AY, et al. HIV pre-exposure prophylaxis in men who have sex with men and transgender women: a secondary analysis of a phase 3 randomised controlled efficacy trial. *The Lancet infectious diseases* 2014; **14**(6): 468-75.
202. Tun W, Kellerman S, Maimane S, et al. HIV-related conspiracy beliefs and its relationships with HIV testing and unprotected sex among men who have sex with men in Tshwane (Pretoria), South Africa. *AIDS care* 2012; **24**(4): 459-67.
203. Eaton LA, Pitpitan EV, Kalichman SC, et al. Men who report recent male and female sex partners in Cape Town, South Africa: an understudied and underserved population. *Archives of Sexual Behavior* 2013; **42**(7): 1299-308.
204. Stephenson R, Rentsch C, Sullivan P. High levels of acceptability of couples-based HIV testing among MSM in South Africa. *AIDS care* 2012; **24**(4): 529-35.
205. Wagenaar BH, Sullivan PS, Stephenson R. HIV knowledge and associated factors among internet-using men who have sex with men (MSM) in South Africa and the United States. *PloS one* 2012; **7**(3): e32915.
206. Batist E, Brown B, Scheibe A, Baral SD, Bekker LG. Outcomes of a community-based HIV-prevention pilot programme for township men who have sex with men in Cape Town, South Africa. *Journal of the International AIDS Society* 2013; **16**: 18754.
207. Knox J, Reddy V, Lane T, Lovasi GS, Hasin D, Sandfort T. Safer sex intentions modify the relationship between substance use and sexual risk behavior among black South African men who have sex with men. *Int J STD AIDS* 2019; **30**(8): 786-94.
208. Maleke K, Makhakhe N, Peters RP, et al. HIV risk and prevention among men who have sex with men in rural South Africa. *Afr J AIDS Res* 2017; **16**(1): 31-8.
209. Rebe K, Lewis D, Myer L, et al. A Cross Sectional Analysis of Gonococcal and Chlamydial Infections among Men-Who-Have-Sex-with-Men in Cape Town, South Africa. *PLoS One* 2015; **10**(9): e0138315.
210. Siegler AJ, Sullivan PS, De Voux A, et al. Exploring repeat HIV testing among men who have sex with men in Cape Town and Port Elizabeth, South Africa. *AIDS care* 2015; **27**(2): 229-34.
211. Lane T, Osmand T, Marr A, et al. The Mpumalanga Men's Study (MPMS): results of a baseline biological and behavioral HIV surveillance survey in two MSM communities in South Africa. *PLoS One* 2014; **9**(11): e111063.
212. Lane T, Osmand T, Marr A, Struthers H, McIntyre JA, Shade SB. Brief Report: High HIV Incidence in a South African Community of Men Who Have Sex With Men: Results From the Mpumalanga Men's Study, 2012–2015. *JAIDS Journal of Acquired Immune Deficiency Syndromes* 2016; **73**(5): 609-11.
213. Maenetje P, Lindan C, Makkan H, et al. HIV incidence and predictors of inconsistent condom use among adult men enrolled into an HIV vaccine preparedness study, Rustenburg, South Africa. *PloS one* 2019; **14**(4): e0214786.
214. Kufa T, Lane T, Manyuchi A, et al. The accuracy of HIV rapid testing in integrated bio-behavioral surveys of men who have sex with men across 5 Provinces in South Africa. *Medicine (Baltimore)* 2017; **96**(28): e7391.

215. Rees K, Radebe O, Arendse C, et al. Utilization of Sexually Transmitted Infection Services at 2 Health Facilities Targeting Men Who Have Sex With Men in South Africa: A Retrospective Analysis of Operational Data. *Sex Transm Dis* 2017; **44**(12): 768-73.
216. van Liere G, Kock MM, Radebe O, et al. High Rate of Repeat Sexually Transmitted Diseases Among Men Who Have Sex With Men in South Africa: A Prospective Cohort Study. *Sex Transm Dis* 2019; **46**(11): e105-e7.
217. Chen Y-H, Gilmore HJ, Maleke K, et al. Increases in HIV status disclosure and sexual communication between South African men who have sex with men and their partners following use of HIV self-testing kits. *AIDS care* 2021; **33**(10): 1262-9.
218. Lippman SA, Lane T, Rabede O, et al. High acceptability and increased HIV testing frequency following introduction of HIV self-testing and network distribution among South African MSM. *Journal of acquired immune deficiency syndromes (1999)* 2018; **77**(3): 279.
219. Lippman SA, Gilmore HJ, Lane T, et al. Ability to use oral fluid and fingerstick HIV self-testing (HIVST) among South African MSM. *PLoS One* 2018; **13**(11): e0206849.
220. Radebe O, Lippman S, Lane T, et al. HIV self-screening distribution preferences and experiences among men who have sex with men in Mpumalanga Province: Informing policy for South Africa. *South African Medical Journal* 2019; **109**(4): 227-31.
221. Sullivan PS, Phaswana-Mafuya N, Baral SD, et al. HIV prevalence and incidence in a cohort of South African men and transgender women who have sex with men: the Sibanye Methods for Prevention Packages Programme (MP3) project. *Journal of the International AIDS Society* 2020; **23**: e25591.
222. Fearon E, Tenza S, Mokoena C, et al. HIV testing, care and viral suppression among men who have sex with men and transgender individuals in Johannesburg, South Africa. *PLoS One* 2020; **15**(6): e0234384.
223. Scheibe A, Young K, Versfeld A, et al. Hepatitis B, hepatitis C and HIV prevalence and related sexual and substance use risk practices among key populations who access HIV prevention, treatment and related services in South Africa: findings from a seven-city cross-sectional survey (2017). *BMC Infect Dis* 2020; **20**(1): 655.
224. Pillay D, Stankevitz K, Lanham M, et al. Factors influencing uptake, continuation, and discontinuation of oral PrEP among clients at sex worker and MSM facilities in South Africa. *PLoS One* 2020; **15**(4): e0228620.
225. Minnis AM, Atujuna M, Browne EN, et al. Preferences for long-acting Pre-Exposure prophylaxis (PreP) for HIV prevention among South African youth: results of a discrete choice experiment. *Journal of the International AIDS Society* 2020; **23**(6): e25528.
226. Montgomery ET, Browne EN, Atujuna M, et al. Long-Acting Injection and Implant Preferences and Trade-Offs for HIV Prevention Among South African Male Youth. *J Acquir Immune Defic Syndr* 2021; **87**(3): 928-36.
227. Metheny N, Stephenson R, Darbes LA, Chavanduka T, Essack Z, van Rooyen H. Correlates of substance misuse, transactional sex, and depressive symptomatology among partnered gay, bisexual and other men who have sex with men in South Africa and Namibia. *AIDS and Behavior* 2022; **26**(6): 2003-14.
228. Stephenson R, Darbes LA, Chavanduka T, Essack Z, van Rooyen H. HIV Testing, Knowledge and Willingness to Use PrEP Among Partnered Men Who Have Sex With Men in South Africa and Namibia. *AIDS Behav* 2021; **25**(7): 1993-2004.
229. Stephenson R, Darbes LA, Chavanduka T, Essack Z, van Rooyen H. Intimate Partner Violence among Male Couples in South Africa and Namibia. *Journal of Family Violence* 2022; **37**(3): 395-405.
